# MOLGENIS VIP: an open-source and modular pipeline for high-throughput and integrated DNA variant analysis

**DOI:** 10.1101/2024.04.11.24305656

**Authors:** W.T.K. Maassen, L.F. Johansson, B. Charbon, D. Hendriksen, S. van den Hoek, M.K. Slofstra, R. Mulder, M.T. Meems-Veldhuis, R. Sietsma, H.H. Lemmink, C.C. van Diemen, M.E. van Gijn, M.A. Swertz, K.J. van der Velde

**Author notes:** These authors contributed equally to this work and share last authorship (M.A. Swertz, K.J. van der Velde).

## Abstract

*In silico* variant interpretation pipelines have become an integral part of genetics research and genome diagnostics. However, challenges remain for automated variant interpretation and candidate shortlisting. Their reliability is affected by variability in input data caused due the use of differing sequencing platforms, erroneous nomenclature and changing experimental conditions. Similarly, differences in predictive algorithms can result in discordant results. Finally, scalability is essential to accommodate large amounts of input data, such as in whole genome sequencing (WGS). To accelerate causal variant detection and innovation in genome diagnostics and research, we developed the MOLGENIS Variant Interpretation Pipeline (VIP). VIP is a flexible open-source computational pipeline that generates interactive reports of variants in whole exome sequencing (WES) and WGS data for expert interpretation. VIP can process short- and long-read data from different platforms and offers tools for increased sensitivity: a configurable decision-tree, filters based on human phenotype ontology (HPO) and gene inheritance that can be used to pinpoint disease-causing variants or finetune a query for specific variants. Here, alongside presenting VIP, we provide a step-by-step protocol for how to use VIP to annotate, classify and filter genetic variants of patients with a rare disease that has a suspected genetic cause. Finally, we demonstrate how VIP performs using 25,664 previously classified variants from the data sharing initiative of the Vereniging van Klinisch Genetische Laboratoriumdiagnostiek (VKGL), a cohort of 18 diagnosed patients from routine diagnostics and a cohort of 41 patients with a rare disease (RD) who were not diagnosed in routine diagnostics but were diagnosed using novel omics approaches within the EU-wide project to solve rare diseases (EU-Solve-RD). VIP requires bioinformatic knowledge to configure, but once configured, any diagnostic professional can perform an analysis within 5 hours.

## Introduction

The field of clinical genetics focuses on the identification of genetic variants that cause disease. By understanding the pathogenesis of genetic diseases, we can provide patients with a prognosis and appropriate treatment^1–3^. Worldwide, around 350 million people are affected by one of 4440 RDs with a known genetic cause^4^. Pinpointing the causal RD variants among all the variants detected in targeted-enrichment sequencing, such as adaptive sampling and whole exome sequencing (WES), or even in whole genome sequencing (WGS) data, is not a trivial task and cannot be performed using a single strategy^2,5–7^. Several *in silico* pipelines have been created to combine published tools into a single solution^2,5^. Alissa Interpret (Agilent) is one example of a commercially developed tool in genome diagnostics, but there are also open-source alternatives, such as Kipoi and Scout^8–10^.

Although the rapid growth of next generation sequencing (NGS) technologies has also accelerated the development of *in silico* pipelines, many challenges remain. To ensure reliability and reproducibility, pipelines must deal with the variability in quality and types of input data caused by different sequencing platforms and experimental conditions. Pipelines also need to accommodate different sources of information with different nomenclatures and ontologies, such as reference genomes, DNA annotation files and knowledge bases^2,11–15^. Further, variability in prediction algorithms, which originates from inherent biases in the datasets used for the development and training of these tools, often causes discordant results^2,16^.

Additionally, the increasing volumes of sequencing data resulting from advances in NGS such as long-read and WGS require large-scale translation of results into the clinic^17^. To accomplish this, scalability must be ensured. Pipelines are also required to adapt to the continuous emergence of new bioinformatic methods and knowledge bases. Ideally, they should also include tools to interpret variants according to the American College of Medical Genetics (ACMG) guidelines^5,6,18^, which helps to accommodate more advanced variant interpretation and prioritization methods^17,19^.

Finally, because variant interpretation pipelines involve different algorithms and resources, their operation can require bioinformatic expertise. Most pipelines are restricted to individual institutions or bound to specific hardware configurations^17^. There are solutions that solve these problems using containerization and parallelization of the processes within the pipelines, but these are far from perfect^17^. Another example that often requires some level of bioinformatic expertise is the presentation and visualization of the results. This is typically of poor quality and difficult to understand, which negatively affects interpretation and translation toward the clinic^20^.

To accelerate causal variant detection and innovation in genome diagnostics and research, we present the MOLGENIS Variant Interpretation Pipeline (VIP). VIP is a flexible open-source pipeline to generate self-contained, interactive reports of variants in WES, WGS, targeted NGS (tNGS) and adaptive sampling data for expert interpretation. To our knowledge, VIP is the only pipeline to incorporate the aforementioned into a single solution. It features integrated best-in-practice algorithms and protocols from routine diagnostics to facilitate classification of coding and non-coding variants according to the ACMG guidelines. VIP applies experience from the VKGL, the EU-wide project to solve rare diseases (EU-Solve-RD), the European Joint Program on Rare Diseases (EJP-RD,) and European infrastructural collaboration to accelerate access and sharing of research data (CINECA) (table 1)^12,21^. We containerized and parallelized VIP for straightforward deployment and large-scale data analysis. VIP is easily modified by updating its tools, annotations, and classification trees. Results are presented in coherent interactive reports for expert interpretation. VIP has already been used in several studies, such as the curation and expansion of human phenotype ontology (HPO) for systemic autoinflammatory diseases and to study the use of a targeted gene panel in Dutch NGS-based newborn screening^22,23^.

**Table 1.**
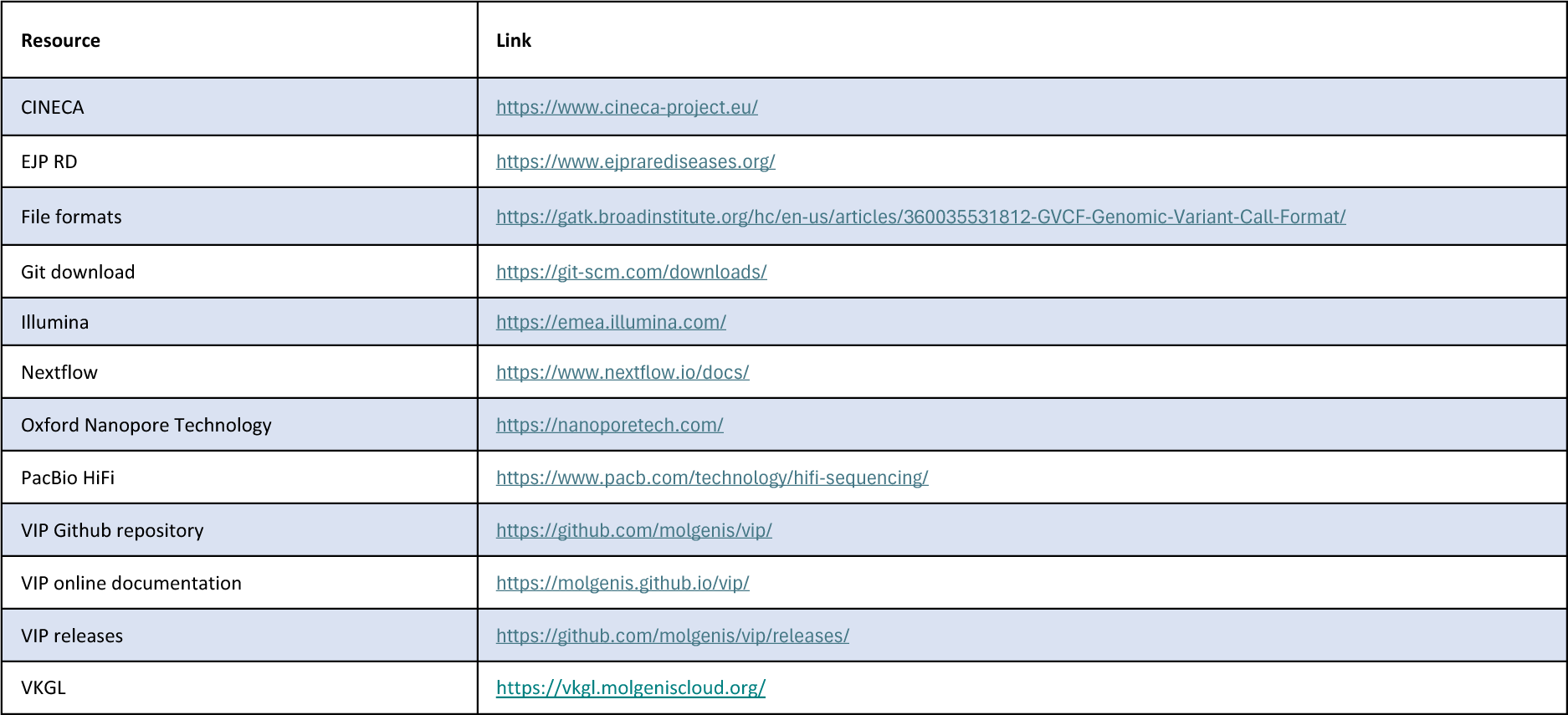
Online resources.

Here, we explain the different capabilities of VIP and how to use them. These include integrated best-in-practice annotation sources, such as our variant pathogenicity predictor CAPICE, and a step-by-step protocol for the analysis of NGS sequencing data, supporting researchers and diagnostic professionals in variant interpretation and classification. We demonstrate VIPs ability to classify and report any clinically relevant germline variants. Additionally, we demonstrate the classification of causal variants in representative cohorts of RD patients from the University Medical Center Groningen (UMCG) and the EU-Solve-RD project^21^.

### MOLGENIS VIP

VIP supports short- and long-read WGS, WES and otherwise targeted-enrichment WGS data from different sequencing platforms: PacBio HiFi long-read sequencing, Oxford Nanopore Tech (ONT) long-read sequencing and Illumina short-read sequencing (table 1). To annotate both coding and non-coding variants, VIP integrates different algorithms and knowledge bases and is divided into four modules: pre-processing, annotation, filtering and interactive reporting (figure 1). Depending on the input format, VIP starts one of four workflows: the FASTQ (raw human-readable sequencing output) workflow, the CRAM/BAM (Compressed Reference-oriented Alignment Map/Binary Alignment Map) workflow, the gVCF (Genomic Variant Call Format) or the VCF (Variant Call Format) workflow (figure 1). The sequence of the pre-processing steps is different for each workflow, but after arriving at a VCF file, the steps performed by the remaining modules are the same. From here, all variants are annotated using the different algorithms, knowledge bases, known genotype-phenotype relationships and inheritance patterns (figure 1, supplementary table 1). Finally, all variants are filtered using a user-specified decision tree and inheritance modes. This results in an interactive report containing a prioritized shortlist of classified variants of interest. Possible classifications are likely benign (LB), benign (B), variant of unknown significance (VUS), likely pathogenic (LP) and pathogenic (P). The raw output with the different annotations is also available as an unfiltered VCF file.

**Figure 1.**
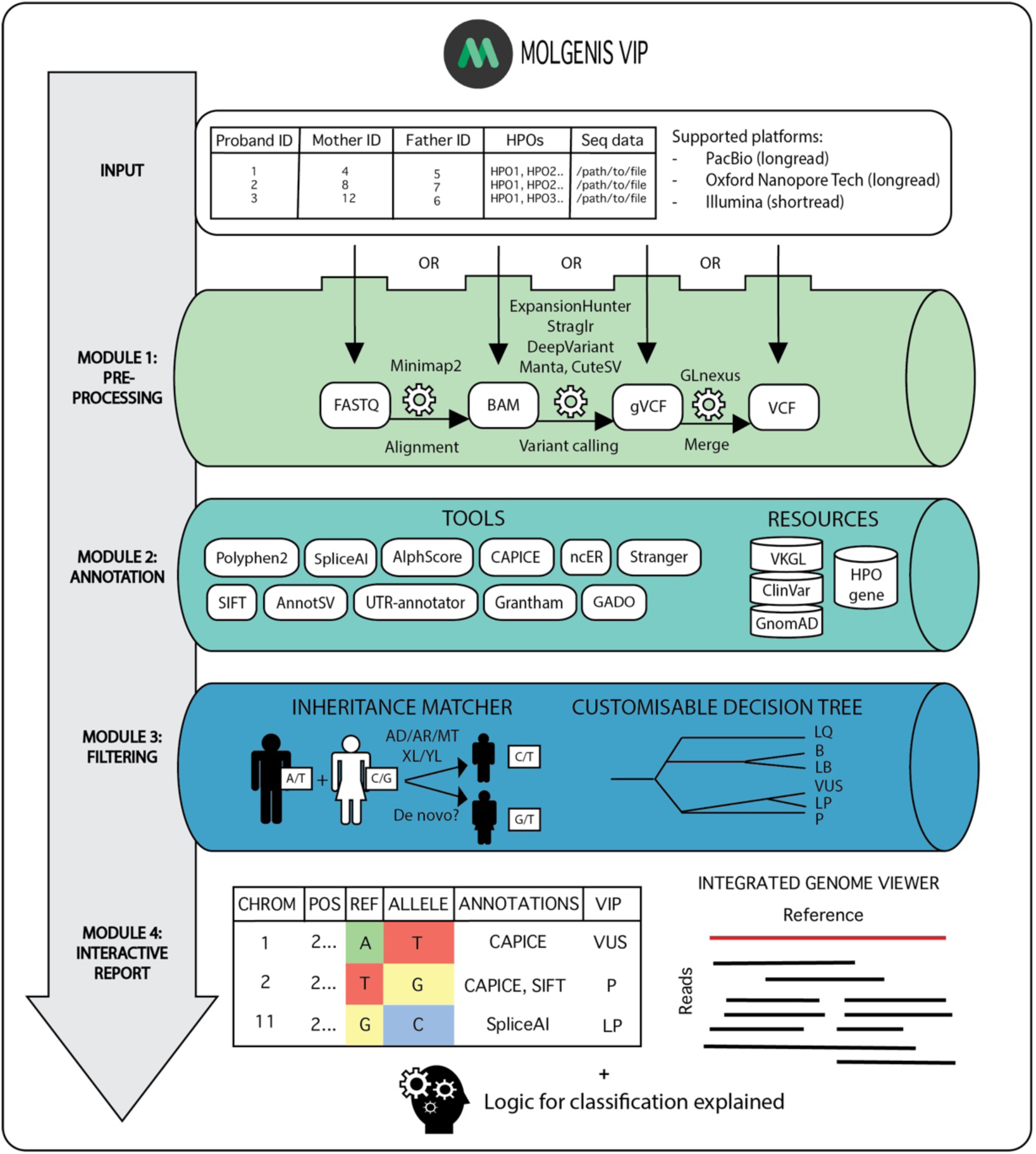
Summary of modules in VIP. As input, VIP requires a sample sheet in which the patient information is specified. In module 1, the input is validated and pre-processed, resulting in a VCF file. The workflow can be started at all points in the pre-processing step. Module 2 provides the variants in the VCF file with annotations from bioinformatic tools and resources (supplementary table 1). Module 3 filters the variants using a customizable decision tree and inheritance information from the previous modules. Finally, VIP generates an interactive report in which the logic for the classifications is explained. Diagnosticians and researchers can use this report for further interpretation of the variants and sharing of the results.

VIP has been developed in a combination of Java, JavaScript, Python, Perl and Shell scripting and runs on the command line. All processes performed by VIP are implemented using the workflow manager Nextflow, allowing parallel processing per chromosome to maximize performance. Apptainer is used for containerization, ensuring straightforward installation and reproducible deployment in different research or diagnostic environments^24,25^. Nextflow also creates intermediate files, allowing users to stop and resume the pipeline at any time during a run. This feature also allows users to analyze large amounts of data step-by-step and helps to accurately follow and reproduce all processes.

### Application of the method

With VIP we aim to provide a complete computational procedure to align reads, followed by calling, annotating, classifying and finally filtering genetic variants for research and diagnostics of patients with a RD with a suspected genetic cause. VIP offers tools to increase sensitivity to find previously unsolved or unknown variants or to finetune a query to find specific variants. The annotation module and decision tree can be altered or expanded to incorporate new annotation tools or resources. Therefore, we encourage users to develop and validate their own decision trees and to expand VIP for their own needs.

### Description of the workflow

#### Input

Diagnosticians and researchers can provide a sample sheet in the tab-separated values (TSV) format in which each row represents a sample taken from an individual. It contains the location of the sequencing data and metadata associated to the sample, such as the location of maternal and paternal sequencing data, phenotypic information, and the reference genome specified and sequencing method used. VIP supports several standard input formats for sequencing data produced by NGS: FASTQ, CRAM, BAM, VCF and gVCF files (table 1)^26–28^. In addition, phenotypic information can be provided using terms from the HPO system^29^.

#### Module 1: pre-processing

When sequencing data is provided in FASTQ format, VIP initiates the FASTQ workflow and maps the files to the specified reference genome using Minimap2^30^. Next, BAM files are aligned and indexed by Samtools. Structural tandem repeats are detected using ExpansionHunter for short-read data and Straglr for long-read data^31,32^. Single nucleotide variants (SNVs), and insertions and deletions (indels) are detected by DeepVariant. GLnexus is used to merge proband, maternal and paternal VCF files into a single VCF for further processing^33^. Finally, structural variants are detected using Manta and cuteSV^34,35^. Sequencing data generated with the following platforms are supported: Illumina short-read (single and paired-end), ONT long-read sequencing and PacBio HiFi long-read sequencing (table 1).

#### Module 2: annotation

Next, the detected variants are annotated with different levels of information. Examples are allele frequencies from GnomAD, known gene-phenotype relationships from the HPO database and classifications validated by the VKGL. VIP also integrates our powerful CAPICE machine learning (ML)– based variant pathogenicity predictor with other best-in-class annotation sources, such as Ensembl Variant Effect Predictor (VEP) and SpliceAI. Supplementary table 1 provides a complete list of the default tools and knowledge bases, including references, that VIP uses to annotate variants. These annotations are used to classify and display variants in the interactive report. We use the plugin framework of Ensembl VEP to extend the existing functionality with new tools and annotations^36^.

The two main annotation tools in VIP are CAPICE and SpliceAI. CAPICE is a ML- based method to predict the pathogenicity of SNVs and indels using diverse genomic features, such as genetic context, gene model annotations and evolutionary constraints. We chose CAPICE as it outperforms other similar pathogenicity predictors, such as CADD and REVEL^37^. Based on the study performed by Li et al. we updated and re-trained CAPICE. Our benchmark study showed that a cut-off value of 0.5 results in a recall rate of 0.95, which we used in our decision tree. SpliceAI is a ML-based method that exploits the sequence surrounding a variant to determine the likelihood that the position in the pre-mRNA transcript is a splice donor or acceptor site^38^. SpliceAI outperforms most other splice variant predictors. Based on a study in which we performed a sensitivity and specificity analysis on published splice variants, we chose a cut-off value of 0.42 as this resulted in the highest sensitivity (supplementary data 1).

### Module 3: filtering

#### Inheritance matcher

To study families for which genotypic and phenotypic information is available, diagnosticians can evaluate if variants segregate with disease or if they occurred *de novo*. This provides an additional level of evidence to determine if a variant is disease-causing. Therefore, VIP uses pedigree information consisting of trios (proband, mother and father) to check if the variant segregation matches disease disease-affected status, while taking into consideration the known inheritance modes of the gene of interest in the Clinical Genomic Database (CGD)^39^. Users can also provide VIP with known inheritance modes from the Online Mendelian Inheritance in Man (OMIM) database or other sources^39,40^. Both maternal and paternal genetic information can be specified in the sample sheet. VIP supports X-linked recessive, X-linked dominant, Y-linked recessive, Y-linked dominant, mitochondrial (MT), autosomal recessive, autosomal dominant and *de novo* inheritance modes. Additionally, VIP supports compound recessive inheritance patterns and inheritance with incomplete penetrance.

#### Customizable decision tree based on best-practices

VIP uses a decision tree to filter variants based on the results of the annotation module, which are collected in an interactive report. Users can compose a decision tree to fit their diagnostic workflow or research question. The decision tree is specified in a human-readable JavaScript object notation (JSON) formatted file. It can be customized by editing the different objects within this file. Each object represents criteria that ultimately lead to the predicted variant classification by VIP. VIP also produces intermediate files so that classification and filtering can be performed repeatedly using different decision trees. Figure 2 shows the default decision tree which is based on the VKGL and the Association for Clinical and Genetic Science (ACGS) guidelines and on collaboration with experts from Genome Diagnostics at the UMCG Genetics department^12,41,42^.

**Figure 2.**
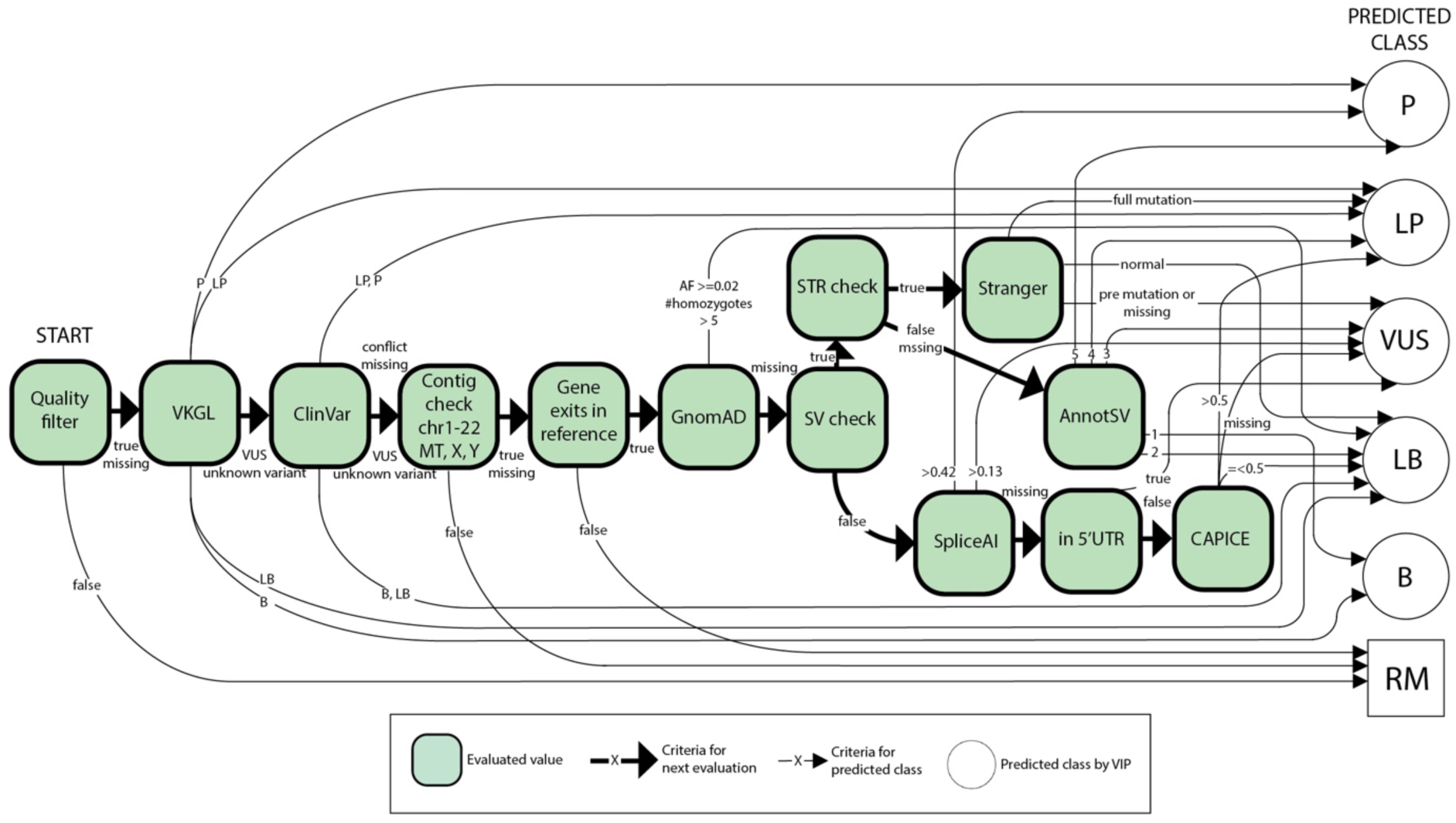
Default decision tree. The figure shows a schematic version of the default decision tree. The green blocks and bold arrows represent the general sequence of filtering steps and the values that are evaluated for each variant (see legend). Each value is calculated in the annotation module (supplementary table 1). Small arrows represent the decisions for the consequence classifications by VIP. VIP classifies the different consequences as B, LB, VUS, LP or P. After a variant is classified by VIP, it exits the filter tree. Variants with incorrect contigs or genes and low quality are removed (RM). Using the JSON format, each component in the decision tree can be customized to fit the workflow of the user.

### Module 4: interactive report

The final set of variants that passed through the decision tree is provided as a self-contained, interactive hypertext markup language (HTML) report that can be opened in any recent internet browser (figure 3). This report contains the filtered list of annotated variants. Diagnosticians can use these annotations to navigate the report and study the criteria that were used to classify the variants. Additionally, basic information, such as the genomic position, reference and alternative alleles, the consequence, and the gene in which the variant is located, are provided. A more detailed view of all available annotations for each transcript is also available. Experts can perform diagnostic-relevant filtering, such as hiding variants with a genotype quality ≤ 20, variants with allelic imbalance and all variants predicted to be LB and B. Similarly, using the HPO-match filter button, variants not located in a gene associated to the HPO terms provided are hidden. Variants that do not match known monogenic inheritance patterns or variants that are not *de novo* can be hidden using the inheritance-match buttons. The report is also provided as a raw VCF file as input for other tools. For the FASTQ and BAM workflows, a genome viewer is available to view the reads that overlap with a variant.

**Figure 3.**
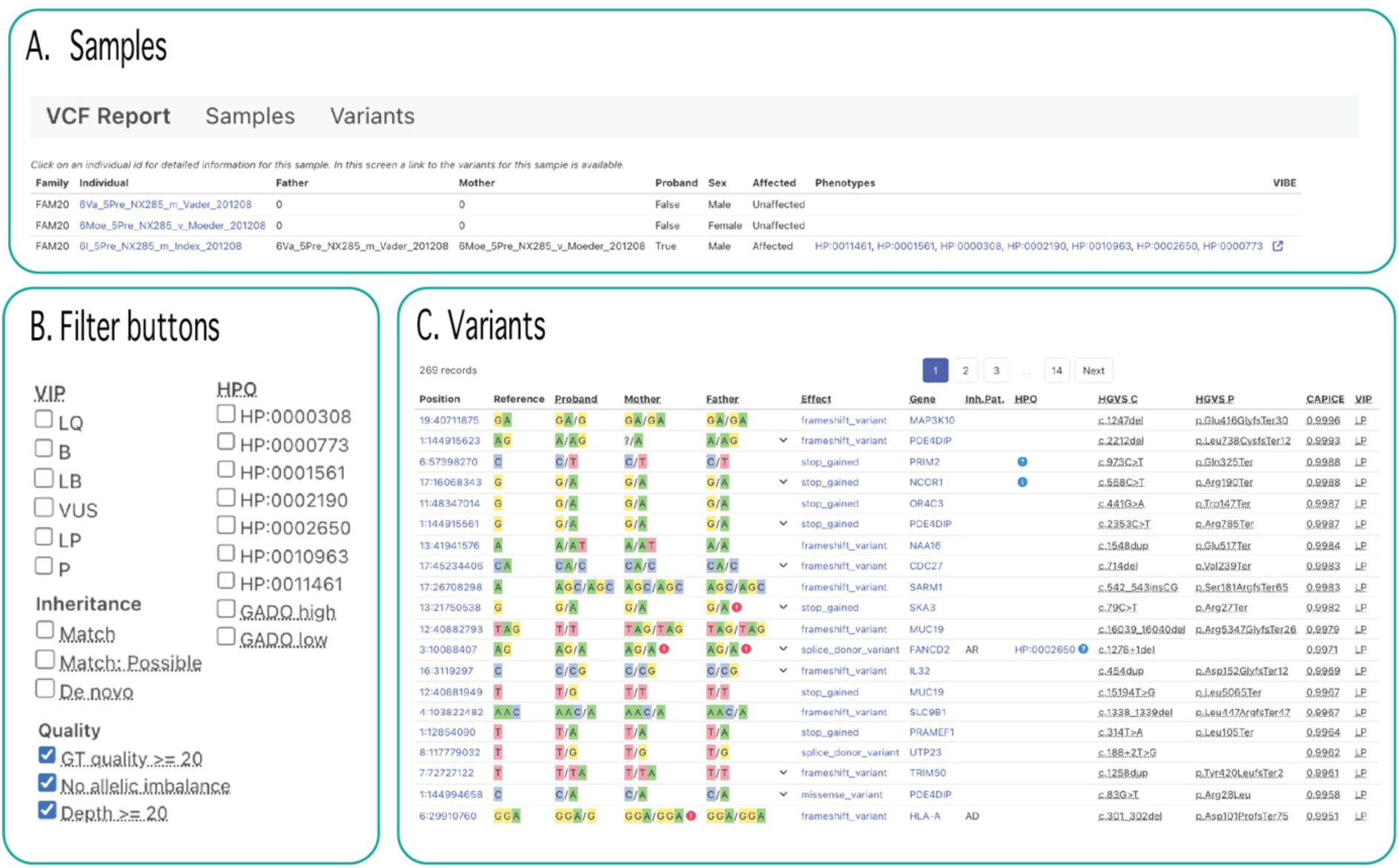
Interactive report. This figure shows an example of the base view of the interactive report. The interactive report opens in the sample screen (A). Here, all the individuals within a family are shown. To navigate to the list of variants with their predicted class, users can navigate to the variant view by clicking one of the individuals (C). The default variant view shows the variants and the consequence of the transcript with the highest CAPICE score. The variants in the list can be filtered using different filters, such as HPO filters, inheritance filters and predicted class filters (B). The report also contains more detailed views to show all annotations for each transcript and which criteria were used to classify a specific variant. When the BAM workflow is used, the built-in genome viewer can be used to study the context of variants within reads that were mapped to the reference genome.

#### Expertise needed

We advise that the installation, configuration and customization of the decision tree be done by a bioinformatician or system administrator. Subsequently, the protocol can be followed by any diagnostician or researcher.

## Materials

### Software

VIP has been developed for GNU-based Linux (e.g. Ubuntu, Windows Subsystem for Linux with x86_64 architecture with installed versions of Bash (version 3.2 or higher), Java (version 11 or higher), Apptainer (version 3.18 or higher) and Nextflow (version 23.10.0). Installation of Git is required to download and install VIP (table 1).

### Hardware

To install the version of VIP that is described in this paper (7.4.0), 220 GB of free diskspace is required. A multicore computer with at least 4 CPUs and 8 GB of RAM is advised, as this allows for parallel processing per chromosome to optimize performance. However, for the FASTQ and BAM workflows, the computing resources required are heavily dependent on the number of input files and the read depth. Similarly, the resources required for the VCF workflow depend heavily on the number of VCF files and the number of variants per sample. To install the most recent version of VIP, we refer to table 1.

## Procedure

Below we describe the procedure to download, install and run the VCF workflow and analyze the results. The same procedure applies to the FASTQ and BAM workflows. For more information on running the FASTQ and BAM workflows, see the online VIP documentation (table 1).

### Installation (1-2 hours)

1. Open a command-line Unix-like terminal, define a directory in which VIP will be installed and navigate to that directory. Download VIP using the following command: git clone https://github.com/molgenis/vip.git. Execute bash vip/install to install VIP. The install process creates a directory vip with all files required to run the application.

### Optional configuration (1 hour)

2. The default configuration files are stored in the directory vip/config. These files specify values for the different parameters in JSON format. For each workflow a custom configuration file can be created to overwrite values in the default configuration. All parameters that are not hardcoded in the config files can be overwritten. Our experience is that the most frequently adjusted parameters are the process parameters for allocating RAM, CPU and processing time limits to specific sub workflows, the reference genome parameters to specify the reference genome used, and the filter parameters to specify which variants are shown in the interactive report. Additionally, users can specify the decision tree that they want to use. For a detailed description of all parameters, with default and example values, see the online VIP documentation (table 1). The custom configuration file used to run the VCF workflow for the demonstration data set is available in supplementary data 2.
3. The file containing the default decision tree in a JSON formatted file is stored in the directory vip/resources. The decision tree can be customized by creating a copy of the default decision tree file and editing or changing the order of the different parameters. For a description of the parameters that can be edited, see the online VIP documentation (table 1).

### Create sample sheet (1 hour)

4. Users need to create a sample sheet containing the information of the individuals within each family. The required and default values are shown in table 2 and the online VIP documentation (table 1). When performing a trio analysis, we recommend providing the family identifier, individual identifier of the proband, the paternal and maternal identifiers, the affected status of the individuals, HPO terms of the proband, the sequencing method used (WES or WGS), the reference assembly used (GRCh37 or GRCh38) and the location of the stored VCF files. Examples of sample sheets used to run the VCF workflow for the demonstration data set are available in supplementary tables 4, 5 and 6.

**Table 2.** Sample sheet format. The first column describes for which workflow the fields need to be filled in. The second column represents the different columns within the samplesheet. The third column contains the description and the requirements of the infromation that can be provided.

| Category | Column | Description |
| --- | --- | --- |
| General | project_id | Unique identifier for each project.<br>Each projects can contain multiple families and individuals.<br>Default is 'vip'. |
|  | family_id | Unique identifier for each family.<br>A family can exit out of multiple layers of a father, mother and children.<br>Default is 'fam_<integer>'. |
|  | individual_id | Unique identifier for each family member.<br>This field is mandatory. |
|  | paternal_id | Unique identifier for each father within a family. |
|  | maternal_id | Unique identifier for each mother within a family. |
|  | sex | Biological sex of family member.<br>Possible values are 'male' and 'female'.<br>Default is 'unknown'. |
|  | affected | Specifies whether the family member is affected by the disease phenotype.<br>Allowed values are 'yes' or 'no'.<br>Default is 'unknown'. |
|  | proband | Specifies whether the family member is a proband.<br>Allowed values are 'yes' or 'no'.<br>If no probands are defined in the sample sheet, then all samples are considered to be probands. |
|  | hpo_ids | Comma-separated unique identifiers from the HPO database describing the phenotypes of the family members within the sample sheet. |
|  | sequencing_method | Specifies the sequencing method used. This determines which workflow-specific processes are used for the processing of the input data.<br>Allowed values are 'WES' or 'WGS'.<br>Default is 'WGS'. |
| Workflow specific |  |  |
| Category | Column | Description |
| FASTQ | fastq | Specifies the absolute path to the file containing raw sequencing data in FASTQ format.<br>Only for single read files.<br>Allowed file extensions are: fastq, fastq.gz, fq and fq.gz. |
|  | fastq_r1 | Specifies the absolute path to the file containing raw sequencing data in FASTQ format for one of the pairs in paired-end sequencing.<br>Allowed file extensions are: fastq, fastq.gz, fq and fq.gz. |
|  | fastq_r2 | Specifies the absolute path to the file containing raw sequencing data in FASTQ format for one of the pairs in paired-end sequencing.<br>Allowed file extensions are: fastq, fastq.gz, fq and fq.gz. |
|  | sequencing_platform | Specifies the sequencing platform used. This determines which workflow-specific processes are used for the processing of the input data.<br>Allowed values are: 'illumina', 'nanopore' or 'pacbio_hifi'.<br>Value must be the same for all project samples. |
| BAM | bam | Specifies the absolute path to the file containing sequencing data in BAM format.<br>Allowed file extensions are: bam, cram and sam. |
|  | sequencing_platform | Specifies the sequencing platform used. This determines which workflow-specific processes are used for the processing of the input data.<br>Allowed values are: 'illumina', 'nanopore' or 'pacbio_hifi'.<br>Value must be the same for all project samples. |
| gVCF | assembly | Identifier specifying the used build of the reference genome.<br>Allowed values are: 'GRCh37', 'GRCh38' and 'T2T'. |
|  | gvcf | Specifies the absolute path to the file containing the called variants in gVCF format.<br>In case a multisample gVCF is used, the individual_id has to match one of the samples in the VCF file. |
| VCF | assembly | Identifier specifying the used build of the reference genome.<br>Allowed values are: 'GRCh37', 'GRCh38' and 'T2T'. |
|  | vcf | Specifies the absolute path to the file containing the called variants in VCF format.<br>In case a multisample VCF is used, the individual_id has to match one of the samples in the VCF file. |

### Running the pipeline (4 hours)

5. To run the VCF workflow, navigate to the installation folder vip and execute vip –workflow vcf –input <PATH_TO_DIRECTORY>/<SAMPLE_SHEET_NAME>.tsv –output <PATH_TO_DIRECTORY>/<OUTPUT_FOLDER_NAME>. If custom configuration values are provided, add -config <PATH_TO_DIRECTORY>/<CONFIG_FILE_NAME>.cfg. The pipeline’s progress is displayed in the standard output of the terminal. After VIP has performed a quality check on the sample sheet, it will run the pre-processing, annotation, filtering and interactive reporting modules. The filtered VCF file, intermediate files, supporting files and interactive reports are stored in the respective output folders (table 3). An interactive report is created for each project specified in the sample sheet. See table 4 for potential problems and suggestions for troubleshooting potential errors.

**Table 3.** Output files. . The first column shows the name of the different output files that are created during a VIP run. The second column shows a description and function of the output files.

| File or folder<sup>1</sup> | Description |
| --- | --- |
| .nextflow<sup>2</sup> | Contains files with run statistics for each Nextflow run. |
| .nxf.home<sup>2</sup>, .nxf.tmp<sup>2</sup> | Contains files and executables used by Nextflow. |
| .nxf.log<sup>2</sup> | Logfile containing history of all performed processes.<br>Can be used to follow each run and debug failed runs. |
| .nxf.work<sup>2</sup> | Folder containing temporary files created by Nextflow for each process.<br>Needed by Nextflow to start subsequent processes.<br>This folder is also used by Nextflow to resume a failed run. |
| nxf_report.html<sup>2</sup> | Self-contained interactive summary of the statistics for each process that Nextflow performed, like runtime, memory and CPU usage.<br>Can be opened in any modern web browser. |
| nxf_timeline.html<sup>2</sup> | Self-contained schematic overview of the runtime and walltime of each Nextflow process.<br>Can be opened in any modern web browser. |
| vip.html | Self-contained interactive report based on variant calls and annotations in vip.vcf.gz. |
| vip.vcf.gz | Compressed VCF file containing all variant calls, annotations and predicted classifications.<br>This file can be used as input for other tools or subsequent analyses that require VCF files as input. |
| vip.vcf.gz.csi | Index file for vip.vcf.gz |
| intermediates | Compressed VCF and index files.<br>Allows user to monitor or rerun VIP starting from pre-defined modules. For different possible rerun options, refer to the VIP documentation (table 1).<br>Contents are dependent on the workflow started.<br>A run of the VCF workflow containing WES data for one trio results in the following most important files:<br>- <project_id>_<fam_id>_<individual_id>_liftover_accepted.vcf.gz<br>- <project_id>_<fam_id>_<individual_id>_liftover_accepted.vcf.gz.csi<br>- <project_id>_<fam_id>_<individual_id>_liftover_rejected.vcf.gz<br>- <project_id>_<fam_id>_<individual_id>_liftover_rejected.vcf.gz.csi<br>- <project_id>_<fam_id>_<individual_id>_annotations.vcf.gz<br>- <project_id>_<fam_id>_<individual_id>_annotations.vcf.gz.csi<br>- <project_id>_<fam_id>_<individual_id>_classifications.vcf.gz<br>- <project_id>_<fam_id>_<individual_id>_classifications.vcf.gz.csi<br>- <project_id>.sample_classifications.vcf.gz<br>- <project_id>.sample_classifications.vcf.gz.csi |
| 1) Path to files is specified by the user with the '--output' option when running VIP on the command line<br>2) For a detailed explanation, refer to the documentation of Nextflow (table 1) |  |

**Table 4.** Troubleshooting table.

| Step | Problem | Possible reason | Solution |
| --- | --- | --- | --- |
| 5 | VIP returns unkown contig in VCF header. | Used reference genome does not match input VCF. | Edit the config file for the corresponding workflow to adjust the reference genome. |
|  | Process runs out of memory. | Not enough memory allocated in the corresponding config file. | Edit the config file for the corresponding process to increase RAM and CPU usage, and time limits for the failed process. |
|  | VIP returns parsing error in the sample sheet validation process. | VIP has found a parsing error in the sample sheet. | Check the nxflow log output file for details about the error. Avoid special characters. Check if the columns line up with tab-separated values. Compare input with required and allowed values by using online VIP documentation and check for inconsistencies (table 1). |
|  | VIP fails during the annotation process (no out-of-memory error). | One of the processes within the annotation module has failed. | Check the nextflow log file for details about the error. Find the process working directory and locate it in the nextflow working directory. Check the command-files to pinpoint which process caused the error. Adjust your input accordingly and rerun your command by adding the resume option. |

### Navigating the interactive report

6. To interpret the results, download and open an interactive report <PROJECT_ID>.html from your output directory in a locally installed web browser. Navigate to the ‘samples tab’ and the proband of interest (figure 3a). Clicking ‘variants’ opens the base view, which displays the possible filters and all the effects of a variant on each possible transcript (figure 3b,c). Each line represents the effect of a variant with the highest CAPICE score. To show all effects for each variant, press the arrow next to an effect. By default, the report only displays the gene symbol, expected inheritance pattern, matching HPO terms, HGVS notations, CAPICE scores, VKGL and ClinVar classifications, GnomAD allele frequencies, predicted classifications by VIP and links to available literature. To display all available annotations, click ‘Variants’ in the header to open a detailed view of all annotations. To use a different sorting method, select an annotation by which to sort in the top right corner.
7. In the base view, use the filters on the left side of the report to filter the list (figure 3b). By default, the read quality, read depth and allelic imbalance filters are applied. To also view low quality variant calls, deselect these filters.
8. When VIP is run using the FASTQ or BAM workflow, click a chromosomal position in the base view to show the genomic context and the reads overlapping the variant for the different transcripts.
9. In the base view, click on an effect to navigate to a list of all annotations for the specific effect. All the annotations added by VIP are displayed in the left panel in the ‘consequence section’. The path through the decision tree is displayed in the right panel. This information can be used to verify which information VIP used to predict a class for a variant. Record- and sample-specific information, such as the chromosomal position and known inheritance mode, is shown in the bottom panel.
10. In the base view, click on a gene symbol to open the gene in a webpage of the HUGO Gene Nomenclature Committee (HGNC) with links to related genomic, clinical and proteomic information^43^.
11. If available, click on the citation in the base view to navigate to the related research article in PubMed.
12. In the base view, click on an HPO term in the variant screen to navigate to a webpage of the HPO database. This contains a description of the phenotype, its ontology and its associations to diseases and genes.
13. In the base view, click on the ClinVar classification to navigate to the associated webpage of ClinVar. This webpage contains detailed information about the variant and the submitted interpretations and evidence for its classifications^44^.

### Reanalysis

14. To re-analyze data from a previous run, VIP can be restarted from the filter module. This is, for example, useful for applying a different decision tree to classify variants without having to annotate all variants again. To rerun VIP, the basic procedure remains the same. In addition, the correct intermediate files should be provided in the sample sheet and the module from which to start should be added in the custom configuration file using the parameter vcf.start. For more instructions to restart VIP from a different module, see the online VIP documentation (table 1).

### Adding additional annotation sources

We also provide bioinformaticians and system administrators the ability to develop their own plugins to add new annotation sources. We use the VEP plugin framework to develop new plugins for VIP. The /vip/resources/vep/plugins directory contains examples with existing plugins. After a plugin is developed, the /vip/modules/vcf/templates/annotate.sh needs to be expanded with the newly developed plugin.

### Timing

Using VIP version 7.4.0, 16 GB of memory and 4 CPUs, the average runtime for WES data in VCF format for 20 samples (average size ∼216 MB) was 4 hours. However, the performance is highly dependent on the size of the dataset, the workflow used and the number of available CPUs and RAM.

#### Preparation (once)

- Step 1. Installing MOLGENIS VIP: 1–2 hours.
- Step 2-3. Creating custom configuration file and decision tree: 1 hour. When the default configuration and decision tree is used, this step can be skipped.

#### For each analysis

- Step 4. Creating the sample sheet: 1 hour.
- Step 5. Running the pipeline: 4 hours (waiting for output).

## Anticipated results

To demonstrate how VIP performs and how it can be used to interpret clinically relevant germline data, we used VIP version 7.4.0, 4 CPUs and 16 GB of RAM to analyze variants from the VKGL database and two anonymized patient cohorts. The total runtime was 4 hours.

### Input data

#### Previously classified variants

We collected 25,664 variants previously classified by experts as LP or P based on the consensus between the different Dutch genome diagnostic laboratories within the VKGL. For this demonstration, we used the LP and P variants in VKGL release 2023-11 and all newly added LP and P variants (added between release 2023-11 and 2024-2) to showcase how VIP classifies previously classified variants (table 1).

#### Routine diagnostics cohort

To demonstrate VIP in diagnostics, we created two anonymized patient cohorts. The first cohort was created to demonstrate how VIP is applied in routine diagnostics at the UMCG. This routine diagnostics cohort contained 18 patients with a molecular genetic diagnosis (supplementary table 2). These patients were selected out of 70 monthly interpretation requests by a clinical geneticist. Virtual gene panels were used to home in on potential disease-causing genes and prevent incidental findings. The panels consisted of genes specific for developmental delay, dilated cardiomyopathy and the clinical exome (supplementary data 3). For all patients HPO terms, WES data and maternal and paternal WES data were available.

#### Solve-RD research cohort

The second cohort contained 41 patients with RDs who had not been diagnosed following routine diagnostics but whose cases were solved within the EU-Solve-RD project using novel omics approaches^21^. These patients were selected based on the availability of a molecular diagnosis, HPO terms and maternal and paternal WES data. The resulting patient selection originated from three different European Reference Networks (ERNs): the ERN for neuromuscular disorders (ERN-EURO NMD), the ERN for rare neurological diseases (ERN-RND) and the ERN for rare malformation syndromes, intellectual and other neurodevelopmental disorders (ERN-ITHACA)^21^. The verified causal variants were extracted from the Genome Phenome analysis platform (GPAP) on the 12^th^ of February 2024^45^ (supplementary table 3).

### Comparing the number of candidate variants, recall rate and ranking

#### Create sample sheets

First, we created a sample sheet containing one file with all LP and P variants in VKGL release 2024-2, including variants that are part of the version of VIP used. Another sample sheet was created containing only newly added variants that are not part of the version of VIP used (supplementary table 4).

Following this step, we created one sample sheet with the patients from the routine diagnostics cohort and one containing the patients from the Solve-RD research cohort. Both sample sheets specified the HPO terms used, affected status, reference genomes, sequencing types and the proband, maternal and paternal WES data (supplementary table 5, 6).

#### Run VIP and analysis of results

VIP was used with the three sample sheets and the default decision tree to create an interactive report for each case. By default, variants with a genotype quality ≤ 20 and allelic imbalance and all LB and B variants were filtered out. We then collected the number of candidate variants when applying: (1) the HPO-match filter, (2) the inheritance-match filter and (3) both the HPO- and inheritance-match filters.

We studied the number of candidate variants per patient and the total number of confirmed molecular diagnoses found in order to compare the recall rates for the previously classified variants, the routine diagnostics cohort and the Solve-RD research cohort. To see how well VIP prioritized the molecular diagnoses, and demonstrate its added value for clinical geneticists, we also compared the assigned rank with the number of candidate variants.

#### Expectation

Because the variants of VKGL release 2023-11 were used by VIP in the VKGL and CAPICE annotation steps, we expect a 100% recall rate for these known variants. To showcase how VIP classifies known VKGL variants that were not part of the VKGL release used by VIP, we used the variants added between release 2023-11 and 2024-2. Because these are also known variants, we expect a recall rate of almost 100%.

The VCF files for the routine diagnostics cases are pre-filtered based on a virtual gene panel. Therefore, we expect that the average number of candidate variants in the interactive report for the routine diagnostics cohort will be smaller than for the Solve-RD research cohort. We also expect that the filtering based on VUS, LP and P variants, HPO term matches and inheritance matches results in the highest ranking of the causal variants. However, this could lead to a decreased recall rate. Not all genes in the HPO database are equally well annotated with HPO terms, and phenotypic features could have been missed when the patient’s phenotype was described in the clinic^22^. Similarly, not all genomic positions for the proband are also covered for both parents, resulting in a missing inheritance match.

The patients from the Solve-RD research cohort that we collected were initially unsolved by the individual expertise centers. For this reason, they were submitted to the Solve-RD project to be analyzed using novel omics methods, such as gene expression analysis and RNA-sequencing, instead of relying only on NGS. We expect that the recall rate for this cohort is lower compared to the routine diagnostics cohort.

### Results

#### Previously classified variants

Figure 4 shows the number of known variants recalled. As expected, VIP detected 100% (25,664/25,664) of these variants. In addition, VIP recalled 558 out of 597 (93.47%) of the variants that were newly added. Twenty-one variants missed because they were classified as LB based on the CAPICE scores. Two variants were missed because one was classified as LB in the ClinVar database and another variant had a gnomAD minor allele frequency (MAF) higher than 0.02. Finally, 16 variants were missed because they had a SpliceAI score between 0.13 and 0.42.

**Figure 4.**
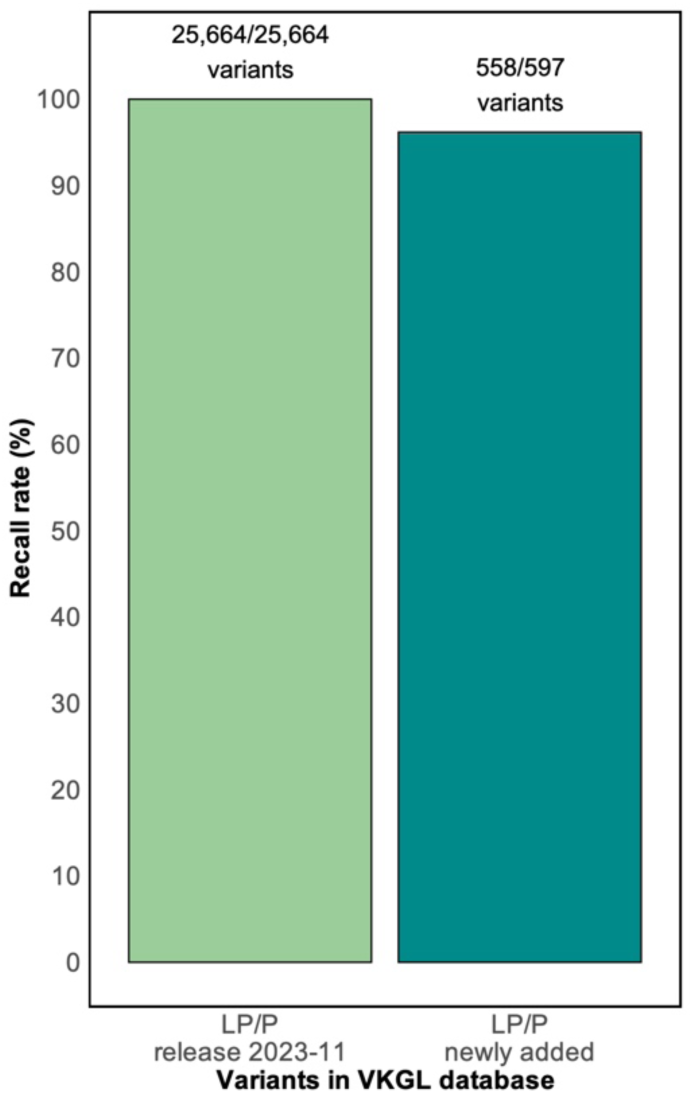
Number of recalled variants that are previously classified. Y-axis shows the percentage of variants that VIP was able to recall as LP or P. The first bar at left represents the variants of VKGL release 2024-2 that were used by VIP in the VKGL and CAPICE annotation steps. The second bar represents the newly added variants between VKGL release 2023-11 and 2024-2 that were not used by VIP. The absolute number of variants that were recalled is shown at the top of the bars.

#### Routine diagnostics cohort

Figure 5a shows the average number of candidate variants per patient and the total number of recalled molecular diagnoses for the routine diagnostics cohort. When LB and B variants are filtered out, VIP returned an average of 338 candidate variants per patient and recalled 18/18 molecular diagnoses. Applying the HPO-match and inheritance-match filters resulted in an average of 8 and 18 candidate variants per patient and 14/18 and 15/18 recalled molecular diagnoses, respectively. The most significant decrease in the number of candidates came after applying the HPO-match and inheritance-match filters simultaneously, resulting in four candidate variants per patient. However, the recall rate decreased to 11/18.

**Figure 5.**
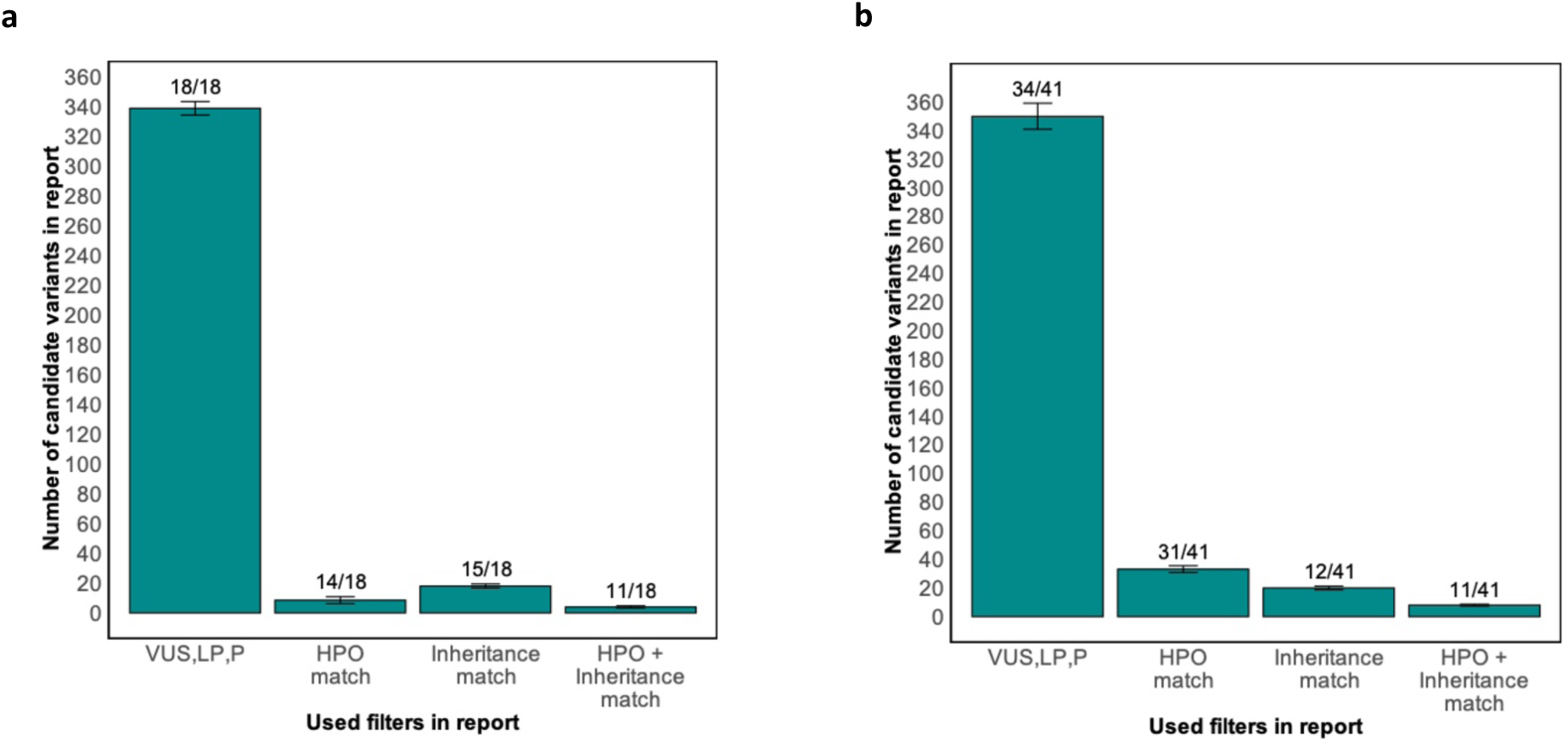
Number of candidate variants and recall rate. **a,** The average numbers of candidate variants per patient in the routine diagnostics cohort. **b,** The average number of candidate variants per patient in the Solve-RD research cohort. On the x-axis, the different filters used in the interactive report are specified. The recall rate per filter is displayed above the individual bars.

#### Solve-RD research cohort

Figure 5b shows the same results for the Solve-RD research cohort. After LB and B variants were filtered out, VIP returned an average of 349 candidate variants per patient and recalled 34/41 molecular diagnoses. Applying the HPO-match and inheritance-match filters resulted in an average of 33 and 20 candidate variants per patient and 31/41 and 12/41 recalled molecular diagnoses, respectively. The most significant decrease in the number of candidate variants came from applying both the HPO-match and inheritance-match filters simultaneously, resulting in eight candidate variants per patient. However, the recall rate decreased to 11/41.

#### Missed variants

In the Solve-RD research cohort, VIP missed 7 molecular diagnoses due to a CAPICE score that did not meet the cut-off value specified in the default decision tree (figure 2). The decrease in recall rate after applying the HPO-match filter can be explained by the fact that 4 patients in the routine diagnostics and 10 patients in the Solve-RD research were annotated with HPO terms that do not match the gene in which the molecular diagnosis is located. This was not unexpected. As described previously by W. Maassen et al., it is possible that not every patient’s phenotype is equally well described in HPO terms or that the gene is not annotated with HPO terms in the HPO database^22^. The decrease in recall rate after applying the inheritance-match filter for patients in the routine diagnostics and the Solve-RD research cohort can be explained by the fact that, for 3 and 29 patients, respectively, NGS did not cover both alleles for at least one parent.

#### Assigned rank versus number of candidate variants

In figure 6, the average rank of the molecular diagnoses was compared with the number of candidate variants per patient in the routine diagnostics and Solve-RD research cohort. This figure shows that the average rank increased and that the average number of candidate variants per patient decreased when the different filters were applied. This shows that VIP was better able to discriminate causal variants from other variants when the number of candidate variants decreases. The lowest number of candidate variants was returned when both the HPO- and the inheritance-match filter were applied.

**Figure 6.**
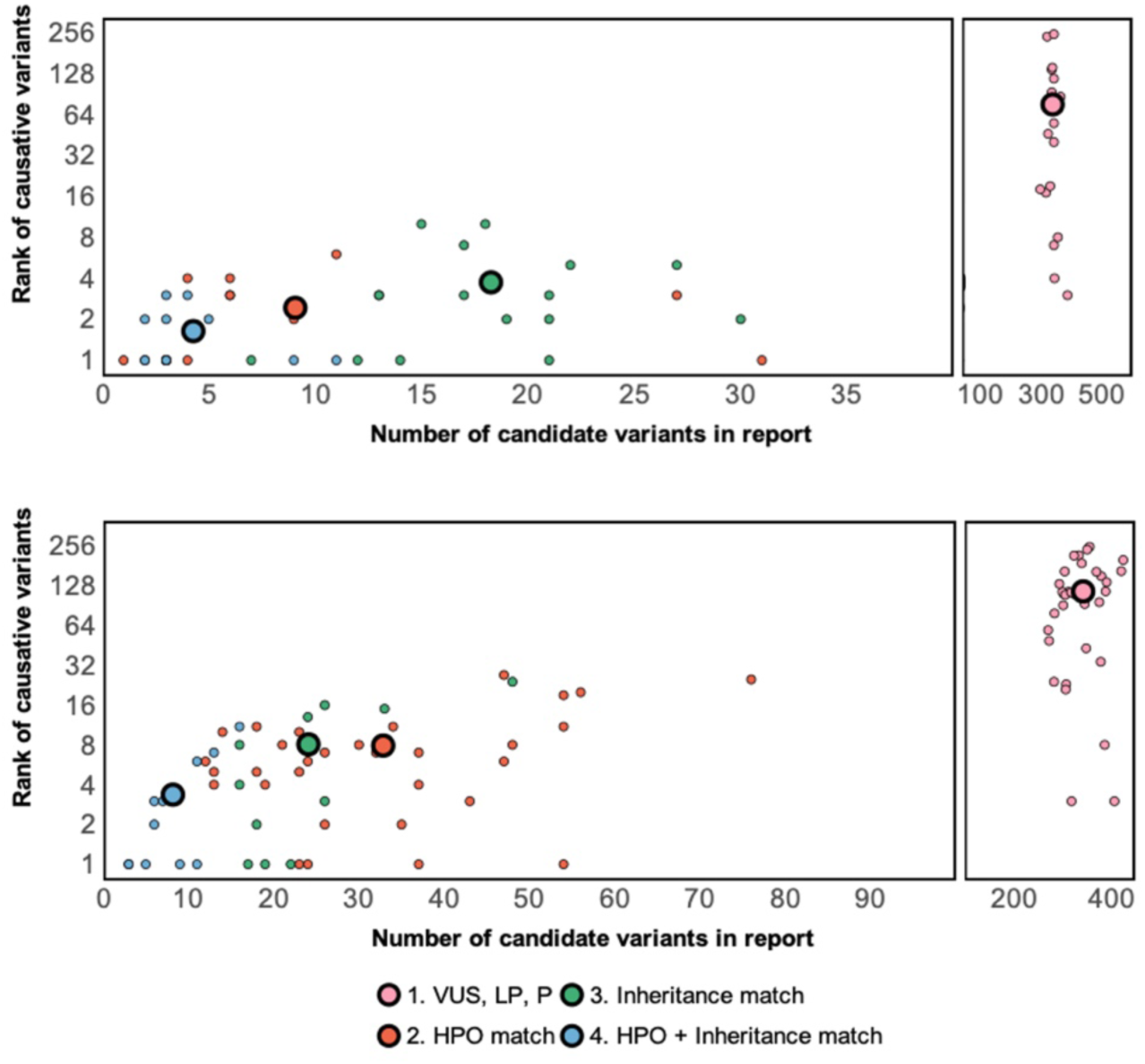
Number of candidate variants per patient and rank of molecular diagnoses. Panels show the average numbers of candidate variants per patient plotted versus the average rank of the molecular diagnosis for the patients in the (**a**) routine diagnostics and (**b**) Solve-RD research cohorts. Patients for whom the molecular diagnoses were not found after applying the different filters are not included.

For the routine diagnostics and Solve-RD research cohort, this resulted in an average rank of 2 and 4, respectively.

## Data availability

The previously classified variants from the VKGL can be downloaded from https://vkgl.molgeniscloud.org/. The routine diagnostics cohort contains patient data from patients within the UMCG and can therefore only be shared upon request. The Solve-RD research cohort is available as a dataset (EGAD50000000390) in the European Genome-Phenome Archive and can be accessed by sending a data access request to the data access committee of the EU-Solve-RD project.

## Ethics statement

The studies involving humans were approved by the ethical committee of the University Medical Center Groningen. The studies were conducted in accordance with the local legislation and institutional requirements. Written informed consent was obtained from the 18 UMCG patients for sharing their WES data. To access patient data from the EU-Solve-RD project, we received confirmation from the Solve-RD Data Access Committee.

## Code availability

MOLGENIS VIP is publicly available at https://github.com/molgenis/vip under the GNU Lesser General Public License v3.0. See https://github.com/molgenis/vip/blob/main/LICENSE for details. VIP is an aggregate work of many individual tools, each covered by their own license(s). Therefore, the individual license(s) of the relevant tools should also be considered.

## Author contributions statements

W.T.K.M., L.F.J. wrote manuscript. W.T.K.M., M.E.G., M.A.S., K.J.V. conceived and designed the experiments. W.T.K.M. performed analysis. W.T.K.M., L.F.J., C.C.D. gathered patient data. L.F.J., B.C., D.H., S.H., M.A.S., R.M., M.M-V., R.S., H.H.L., C.C.D., M.E.G., M.A.S., K.J.V. discussed results and reviewed paper. M.E.G., M.A.S., K.J.V. supervised project. D.H., B.C., S.H., M.A.S., R.S., W.T.K.M., L.F.J., K.J.V. developed and tested the software. All authors contributed to the results and approved the submitted version.

## Acknowledgements

This study has been performed with the support of the members of the UMCG Genomics Coordination Center, the MOLGENIS VIP development team and the Development and Innovation team at the Genetics Department of the UMCG. We want to thank all members and patients participating in the Solve-RD project for access to the Solve-RD database and the use of the GPAP analysis software. Finally, we would like to thank Kate Mc Intyre for editorial assistance.

This study received funding from the EU projects Solve-RD, EJP-RD and CINECA (H2020 779257, H2020 825575, H2020 825775, respectively) and NWO grant numbers 917.164.455 and 184.034.019.

