## supplementary data 1 for "MOLGENIS VIP: an open-source and modular pipeline for high-throughput and integrated DNA variant analysis"

### Summary of SpliceAI benchmark study

##### Selection of variants and datasets

SpliceAI is a machine-learning-based method that exploits the sequence surrounding a variant to determine the likelihood that the position in the pre-mRNA transcript is a splice donor or acceptor site. SpliceAI calculates a delta score from 0 to 1[1].

##### Selection of variants and datasets

After performing a literature search we found 37 papers in which minigene assays were performed to confirm splice-altering effects of 276 variants. Of these variants, 170 were confirmed to have a splice-altering effect and 106 did not have a splice-altering effect (table 1).

| Search | | Variants | | Date of search |
|---|---|---|---|---|
| | Papers | Splice | Non-splice | |
| Variants in epilepsy minigene (2011-2021) | 9 | 15 | 2 | 17-02-2021 |
| Variants in skeletal dysplasia minigene (2011-2021) | 5 | 7 | 2 | 20-02-2021 |
| Variants in hereditary cancer minigene (2011-2021 | 19 | 138 | 92 | 23-02-2021 |
| Variants in cholestasis minigene (2011-2021) | 4 | 10 | 9 | 04-03-2021 |

**Table 1** This table shows the different numbers of variants found accross papers researching epilepsy, skeletal dysplasia, hereditary cancer and cholestasis.

##### Benchmark

From the variants, 101 splice-altering variants and 64 non-splice-altering variants were used to perform sensitivity-specificity analysis (figure 1). After having performed an ROC analysis the AUROC was 0.9838.

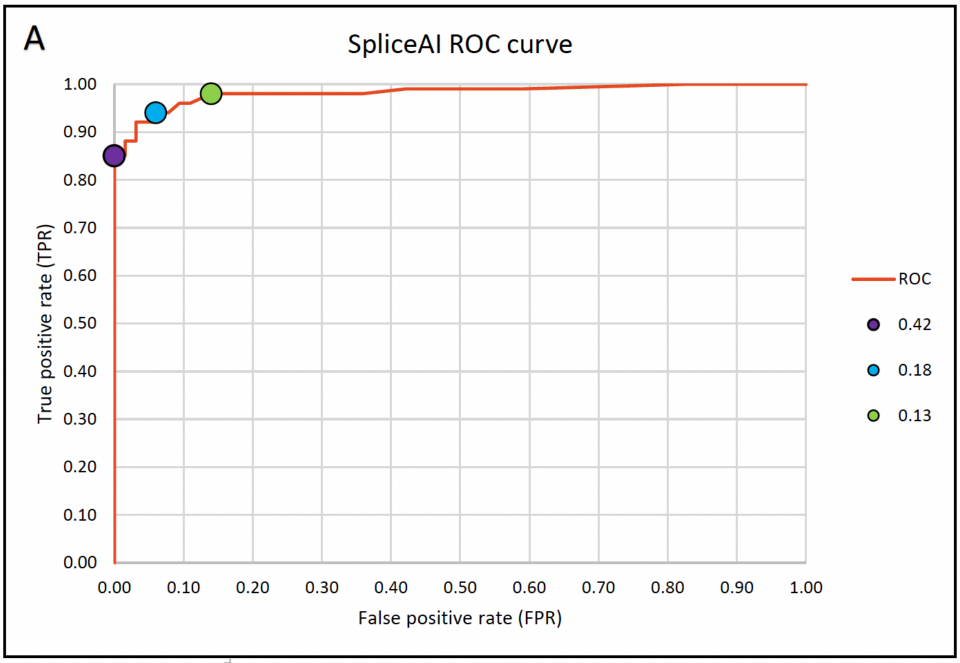


| Threshold | Sensitivity | Specificity | Accuracy |
|---|---|---|---|
| Δ0.13 | 0.86 | 0.98 | 0.93 |
| Δ0.18 | 0.94 | 0.94 | 0.94 |
| Δ0.42 | 1.00 | 0.85 | 0.91 |

**Figure 1** Panel A shows the ROC based on the true positive rate (y-axis) and false positive rate (x-axis). The table below shows the different sensitivity, specificy and accuracy values for the different determined thresholds.

Subsequently, we identified three delta score thresholds of interest: 0.13 for favoring specificiy, 0.42 for favoring sensitivity and 0.18 for favoring high accuracy. The Youden's J statistics for the 0.13, 0.18 and 0.42 thresholds were 0.84, 0.88 and 0.85, respectively.

Finally, the performance of the different delta score thresholds was compared for different subsets of variants: "canonocal", "intronic", "exonic between 1 and 5 basepairs away from the exon-intron boundary" and "exonic >5 basepairs away from the exon-intron boundary". Figure 2 shows the calculated sensitivity and specificity for the different groups. The trade-off between sensitivity and specificity for the 0.42 and 0.18 thresholds seems more beneficial for the "exonic splice variants >5 basepairs away from the exon-intron boundary". However, in terms of errors, the trade-off is three less false negative results and two more false positive results. Assuming that avoiding false positive results is favored in diagnostics, this data suggests that there is no significant difference in performance between the individual groups and the complete set.

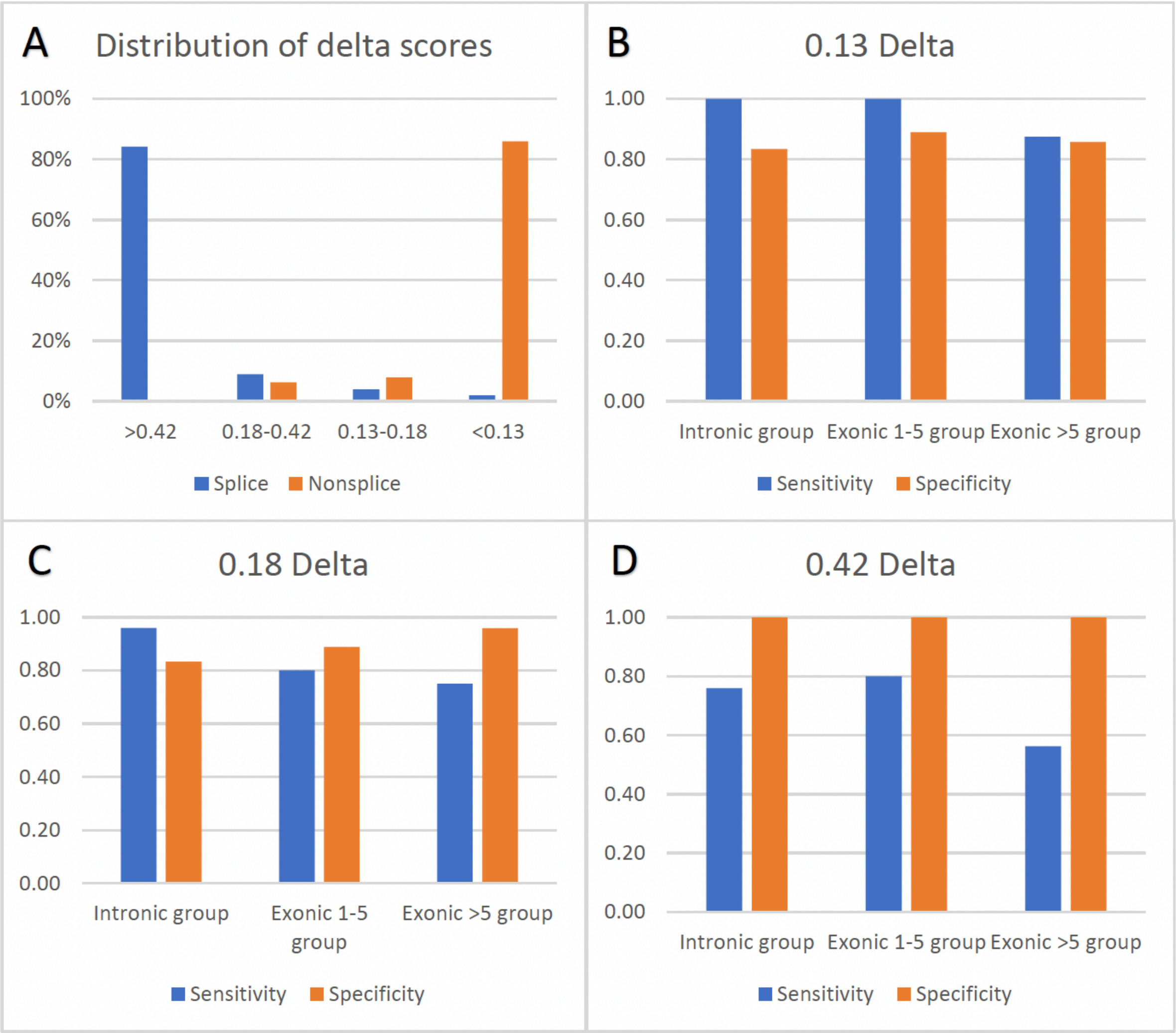


**Figure 2** Panel A shows the distribution in percentage of splice and non-splice variants that have a delta score that falls within one of the following categories: >0.42, between 0.18 and 0.42, between 0.13 and 0.18 and <0.13. Panel B to D show the sensitivity and specificity for the different splice variant groups using the different cut-off values.
