## supplementary data 3 for "MOLGENIS VIP: an open-source and modular pipeline for high-throughput and integrated DNA variant analysis"

| Gene panel developmental delay | Gene panel dilated cardiomyopathy | Gene panel clinical exome |
|---|---|---|
| A2ML1 | AARS2 | A2M |
| AAAS | ABCC6 | A2ML1 |
| AARS | ABCC9 | A4GALT |
| AASS | ACAD8 | AAAS |
| ABAT | ACAD9 | AAGAB |
| ABCC8 | ACADS | AARS |
| ABCC9 | ACADVL | AARS2 |
| ABCD1 | ACTA1 | AASS |
| ABCD4 | ACTC1 | ABAT |
| ABHD5 | ACTN2 | ABCA1 |
| ACAD9 | ADCY5 | ABCA12 |
| ACADS | AGK | ABCA3 |
| ACAT1 | AGL | ABCA4 |
| ACO2 | AGPAT2 | ABCA5 |
| ACOX1 | AHCY | ABCA7 |
| ACSF3 | AIP | ABCB1 |
| ACSL4 | ALG1 | ABCB11 |
| ACTB | ALG6 | ABCB4 |
| ACTG1 | ALMS1 | ABCB6 |
| ACTL6A | ALPK3 | ABCB7 |
| ACVR1 | ANKRD1 | ABCC11 |
| ACY1 | ANKRD11 | ABCC2 |
| ADAM22 | ANKS6 | ABCC6 |
| ADAR | ANO5 | ABCC8 |
| ADAT3 | APOPT1 | ABCC9 |
| ADGRG1 | ARSB | ABCD1 |
| ADK | ASNA1 | ABCD3 |
| ADNP | ATP5F1E | ABCD4 |
| ADSL | ATP6V1B2 | ABCG2 |
| AFF2 | ATPAF2 | ABCG5 |
| AFF4 | BAG3 | ABCG8 |
| AFG3L2 | BBS2 | ABHD12 |
| AGA | BCS1L | ABHD5 |
| AGO2 | BOLA3 | ABL1 |
| AGPAT2 | BRAF | ABO |
| AGTR2 | BRCA2 | ACAD8 |
| AHCY | BRCC3 | ACAD9 |
| AHDC1 | BRIP1 | ACADL |
| AHI1 | BSCL2 | ACADM |
| AIFM1 | CALR3 | ACADS |
| AIMP1 | CAP2 | ACADSB |
| AIMP2 | CASQ1 | ACADVL |
| AK1 | CAV1 | ACAN |
| AKT3 | CAV3 | ACAT1 |
| ALDH18A1 | CAVIN4 | ACBD5 |

| | | |
|---|---|---|
| ALDH3A2 | CDKN1C | ACE |
| ALDH4A1 | CHKB | ACHE |
| ALDH5A1 | CHRM2 | ACKR1 |
| ALDH7A1 | CISD2 | ACO2 |
| ALG1 | COA5 | ACOX1 |
| ALG11 | COA6 | ACOX2 |
| ALG12 | COG7 | ACP33 |
| ALG13 | COL7A1 | ACP4 |
| ALG2 | COQ2 | ACP5 |
| ALG3 | COQ4 | ACSF3 |
| ALG6 | COX10 | ACSL4 |
| ALG8 | COX14 | ACSL6 |
| ALG9 | COX20 | ACTA1 |
| ALMS1 | COX6B1 | ACTA2 |
| ALX1 | COX7B | ACTB |
| ALX4 | CPT1A | ACTC1 |
| AMER1 | CPT2 | ACTG1 |
| AMMECR1 | CRYAB | ACTG2 |
| AMN | CSRP3 | ACTL6A |
| AMPD2 | CTNNA3 | ACTN1 |
| AMT | D2HGDH | ACTN2 |
| ANK3 | DCAF8 | ACTN3 |
| ANKH | DCHS1 | ACTN4 |
| ANKLE2 | DES | ACVR1 |
| ANKRD11 | DLD | ACVR1B |
| ANO10 | DMD | ACVR2B |
| ANTXR1 | DMPK | ACVRL1 |
| AP1S1 | DNAJC19 | ACY1 |
| AP1S2 | DOLK | ADA |
| AP3B1 | DPM3 | ADA2 |
| AP3B2 | DSC2 | ADAM10 |
| AP3D1 | DSG2 | ADAM17 |
| AP4B1 | DSP | ADAM22 |
| AP4E1 | DTNA | ADAM9 |
| AP4M1 | DYSF | ADAMTS10 |
| AP4S1 | ELAC2 | ADAMTS13 |
| AP5Z1 | EMD | ADAMTS17 |
| APC2 | ENPP1 | ADAMTS18 |
| APOPT1 | EPG5 | ADAMTS2 |
| APTX | ERBB3 | ADAMTS3 |
| ARCN1 | ERCC4 | ADAMTS5 |
| ARFGEF2 | EYA4 | ADAMTSL2 |
| ARG1 | FAH | ADAMTSL4 |
| ARHGAP31 | FANCA | ADAR |
| ARHGEF6 | FANCB | ADAT3 |
| ARHGEF9 | FANCC | ADCY1 |
| ARID1A | FANCD2 | ADCY10 |
| ARID1B | FANCE | ADCY5 |

| | | |
|---|---|---|
| ARID2 | FANCF | ADCY6 |
| ARL13B | FANCG | ADD1 |
| ARL6 | FANCI | ADD3 |
| ARMC9 | FANCL | ADGRE2 |
| ARSA | FANCM | ADGRG1 |
| ARSE | FASTKD2 | ADGRG2 |
| ARV1 | FBN1 | ADGRG6 |
| ARX | FBXL4 | ADGRV1 |
| ASAH1 | FHL1 | ADH1B |
| ASCL1 | FHL2 | ADH1C |
| ASH1L | FIG4 | ADIPOQ |
| ASL | FKRP | ADK |
| ASNS | FKTN | ADNP |
| ASPA | FLNA | ADRA2B |
| ASPM | FLNC | ADRA2C |
| ASS1 | FOS | ADRB1 |
| ASXL1 | FOXD4 | ADRB2 |
| ASXL2 | FOXRED1 | ADRB3 |
| ASXL3 | FTO | ADSL |
| ATAD1 | FXN | ADSSL1 |
| ATAD3A | GAA | AF10 |
| ATCAY | GATA4 | AFF2 |
| ATIC | GATAD1 | AFF4 |
| ATL1 | GATB | AFG3L2 |
| ATN1 | GATC | AFP |
| ATP1A1 | GBE1 | AGA |
| ATP1A2 | GJA5 | AGBL1 |
| ATP1A3 | GLA | AGBL5 |
| ATP2A2 | GLB1 | AGK |
| ATP6AP2 | GMPPB | AGL |
| ATP6V0A2 | GNAS | AGO2 |
| ATP6V1A | GNPTAB | AGPAT2 |
| ATP6V1B2 | GNS | AGPS |
| ATP7A | GPC3 | AGRN |
| ATP7B | GPR101 | AGRP |
| ATP8A2 | GSN | AGT |
| ATPAF2 | GTPBP3 | AGTR1 |
| ATR | GYS1 | AGTR2 |
| ATRX | HADH | AGXT |
| AUH | HADHA | AHCY |
| AUTS2 | HADHB | AHDC1 |
| AVPR2 | HAMP | AHI1 |
| B3GALNT2 | HBB | AHNAK |
| B3GALT6 | HCCS | AHSP |
| B3GLCT | HCN4 | AICDA |
| B4GALNT1 | HFE | AIFM1 |
| B4GALT1 | HGSNAT | AIMP1 |
| B4GALT7 | HJV | AIMP2 |

| | | |
|---|---|---|
| B4GAT1 | HPS1 | AIPL1 |
| BBS1 | HRAS | AIRE |
| BBS10 | HSD17B10 | AK1 |
| BBS12 | HSPB6 | AK2 |
| BBS2 | IDH2 | AKAP10 |
| BBS4 | IDUA | AKAP9 |
| BBS5 | IGF2 | AKR1C2 |
| BBS7 | ILK | AKR1C4 |
| BBS9 | ISL1 | AKR1D1 |
| BCAP31 | ITGB1BP2 | AKT1 |
| BCKDHA | JPH2 | AKT2 |
| BCKDHB | JUP | AKT3 |
| BCKDK | KCNH1 | ALAD |
| BCL11A | KCNH2 | ALAS2 |
| BCL11B | KIF20A | ALB |
| BCOR | KLF1 | ALDH18A1 |
| BCORL1 | KRAS | ALDH1A2 |
| BCS1L | LAMA4 | ALDH1A3 |
| BLM | LAMP2 | ALDH1B1 |
| BOLA3 | LDB3 | ALDH2 |
| BPTF | LEMD2 | ALDH3A2 |
| BRAF | LIAS | ALDH4A1 |
| BRAT1 | LMNA | ALDH5A1 |
| BRD4 | MAP2K1 | ALDH6A1 |
| BRF1 | MAP2K2 | ALDH7A1 |
| BRPF1 | MGME1 | ALDOA |
| BRSK2 | MIB1 | ALDOB |
| BRWD3 | MLYCD | ALG1 |
| BSCL2 | MMUT | ALG10 |
| BTD | MRPL3 | ALG11 |
| BUB1B | MRPL44 | ALG12 |
| C12orf4 | MRPS22 | ALG13 |
| C12orf57 | MTCP1 | ALG14 |
| C12orf65 | MTO1 | ALG2 |
| C2CD3 | MYBPC3 | ALG3 |
| CA2 | MYH6 | ALG6 |
| CA5A | MYH7 | ALG8 |
| CA8 | MYL2 | ALG9 |
| CACNA1A | MYL3 | ALMS1 |
| CACNA1C | MYLK2 | ALOX12B |
| CACNA1D | MYOT | ALOX5 |
| CACNA1E | MYOZ1 | ALOX5AP |
| CACNA1G | MYOZ2 | ALOXE3 |
| CACNA2D1 | MYPN | ALPI |
| CACNG2 | NAGA | ALPK3 |
| CAD | NAGLU | ALPL |
| CAMK2A | NDUFA1 | ALS2 |
| CAMK2B | NDUFA11 | ALS2CL |

| | | |
|---|---|---|
| CAMTA1 | NDUFAF1 | ALX1 |
| CANT1 | NDUFAF2 | ALX3 |
| CAPN10 | NDUFAF3 | ALX4 |
| CARS2 | NDUFAF4 | AMACR |
| CASK | NDUFAF5 | AMBN |
| CBL | NDUFB11 | AMELX |
| CBLIF | NDUFB3 | AMER1 |
| CBS | NDUFB9 | AMH |
| CC2D1A | NDUFS1 | AMHR2 |
| CC2D2A | NDUFS2 | AMMECR1 |
| CCBE1 | NDUFS3 | AMN |
| CCDC115 | NDUFS4 | AMPD1 |
| CCDC174 | NDUFS6 | AMPD2 |
| CCDC22 | NDUFV1 | AMPD3 |
| CCDC78 | NDUFV2 | AMT |
| CCDC88A | NEBL | ANG |
| CCDC88C | NEU1 | ANGPT1 |
| CCND2 | NEXN | ANGPTL3 |
| CCNK | NF1 | ANGPTL4 |
| CDC42 | NKX2-5 | ANK1 |
| CDC6 | NONO | ANK2 |
| CDH11 | NPPA | ANK3 |
| CDH15 | NRAS | ANKH |
| CDK10 | NSD1 | ANKK1 |
| CDK13 | NUBPL | ANKLE2 |
| CDK5RAP2 | PALB2 | ANKRD1 |
| CDK8 | PCCA | ANKRD11 |
| CDKL5 | PCCB | ANKRD26 |
| CDKN1C | PDGFRA | ANKS3 |
| CDON | PDLIM3 | ANKS6 |
| CENPF | PET100 | ANKZF1 |
| CENPJ | PEX1 | ANLN |
| CEP104 | PEX10 | ANO10 |
| CEP120 | PEX11B | ANO3 |
| CEP135 | PEX12 | ANO5 |
| CEP152 | PEX13 | ANO6 |
| CEP290 | PEX14 | ANOS1 |
| CEP41 | PEX16 | ANTXR1 |
| CEP57 | PEX19 | ANTXR2 |
| CEP83 | PEX2 | ANXA5 |
| CEP89 | PEX26 | AP1S1 |
| CHAMP1 | PEX3 | AP1S2 |
| CHD1 | PEX5 | AP1S3 |
| CHD2 | PEX6 | AP2S1 |
| CHD3 | PEX7 | AP3B1 |
| CHD4 | PGM1 | AP3B2 |
| CHD7 | PHYH | AP3D1 |
| CHD8 | PIGT | AP4B1 |

| | | |
|---|---|---|
| CHKB | PKP2 | AP4E1 |
| CHMP1A | PLEC | AP4M1 |
| CHRNA4 | PLEKHM2 | AP4S1 |
| CIC | PLN | AP5Z1 |
| CIT | PMM2 | APC2 |
| CKAP2L | PNPLA2 | APCDD1 |
| CLCN4 | POLG | APOA1 |
| CLCNKB | POMT1 | APOA2 |
| CLIC2 | POU1F1 | APOA5 |
| CLIP1 | PPARG | APOB |
| CLN3 | PPCS | APOC2 |
| CLN5 | PRDM16 | APOC3 |
| CLN6 | PRKAG2 | APOL1 |
| CLN8 | PRPS1 | APOPT1 |
| CLP1 | PSEN1 | APPL1 |
| CLPB | PSEN2 | APRT |
| CLTC | PTPN11 | APTX |
| CNKSR2 | QRSL1 | AQP1 |
| CNNM2 | RAB3GAP2 | AQP2 |
| CNOT1 | RAD51C | AQP3 |
| CNOT3 | RAF1 | AQP5 |
| CNPY3 | RBCK1 | AQP7 |
| CNTNAP2 | RBM20 | ARCN1 |
| COASY | RET | ARFGEF2 |
| COG1 | RIT1 | ARG1 |
| COG4 | RMND1 | ARHGAP26 |
| COG5 | RYR2 | ARHGAP31 |
| COG6 | SCARB2 | ARHGDIA |
| COG7 | SCN5A | ARHGEF1 |
| COG8 | SCO1 | ARHGEF10 |
| COL4A1 | SCO2 | ARHGEF2 |
| COL4A2 | SDHA | ARHGEF6 |
| COL4A3BP | SDHAF1 | ARHGEF9 |
| COLEC11 | SDHD | ARID1A |
| COQ2 | SELENON | ARID1B |
| COQ4 | SGCA | ARID2 |
| COQ8A | SGCB | ARL13B |
| COQ9 | SGCD | ARL2BP |
| COX10 | SGSH | ARL3 |
| COX15 | SHOC2 | ARL6 |
| COX6B1 | SKI | ARL6IP1 |
| CP | SLC19A2 | ARMC4 |
| CPLANE1 | SLC22A5 | ARMC5 |
| CPLX1 | SLC25A20 | ARMC9 |
| CPS1 | SLC25A3 | ARNT2 |
| CRADD | SLC25A4 | ARPC1B |
| CRBN | SLC2A10 | ARSA |
| CREBBP | SLX4 | ARSB |

| | | |
|---|---|---|
| CRLF1 | SNAP29 | ARSE |
| CSNK2A1 | SOD2 | ART4 |
| CSNK2B | SOS1 | ARV1 |
| CSPP1 | SPEG | ARX |
| CSTB | SYNE1 | ASAH1 |
| CTBP1 | SYNE2 | ASB10 |
| CTCF | TACO1 | ASCC1 |
| CTDP1 | TAZ | ASCL1 |
| CTNNA2 | TBX20 | ASH1L |
| CTNNB1 | TCAP | ASIP |
| CTNND1 | TEAD1 | ASL |
| CTNND2 | TERT | ASNA1 |
| CTSA | TGFB1 | ASNS |
| CTSD | TGFB3 | ASPA |
| CTTNBP2 | TIMM50 | ASPH |
| CUBN | TMEM126A | ASPM |
| CUL4B | TMEM43 | ASPN |
| CUX1 | TMEM70 | ASPSCR1 |
| CUX2 | TMPO | ASS1 |
| CWC27 | TNNC1 | ASXL1 |
| CWF19L1 | TNNI3 | ASXL2 |
| CXorf56 | TNNI3K | ASXL3 |
| CYB5R3 | TNNT2 | ATAD1 |
| CYP27A1 | TPI1 | ATAD3A |
| CYP2U1 | TPM1 | ATAD3B |
| D2HGDH | TPM3 | ATCAY |
| DAG1 | TRNT1 | ATF6 |
| DARS | TSFM | ATG16L1 |
| DARS2 | TTN | ATIC |
| DBT | TTPA | ATL1 |
| DCAF17 | TTR | ATL3 |
| DCC | TUFM | ATM |
| DCHS1 | TWNK | ATN1 |
| DCPS | TXNRD2 | ATOH7 |
| DCX | UBE2T | ATP13A2 |
| DDC | UBR1 | ATP1A1 |
| DDHD2 | VCL | ATP1A2 |
| DDX11 | VPS13A | ATP1A3 |
| DDX3X | WFS1 | ATP2A1 |
| DDX59 | XK | ATP2A2 |
| DEAF1 | XPNPEP3 | ATP2B2 |
| DENND5A | YARS2 | ATP2B3 |
| DEPDC5 | | ATP2C1 |
| DHCR24 | | ATP5F1A |
| DHCR7 | | ATP5F1B |
| DHDDS | | ATP5F1C |
| DHFR | | ATP5F1D |
| DHTKD1 | | ATP5F1E |

| | |
|---|---|
| DHX30 | ATP5IF1 |
| DIAPH1 | ATP5MC1 |
| DIP2B | ATP5MC2 |
| DIS3L2 | ATP5MC3 |
| DKC1 | ATP5ME |
| DLAT | ATP5MF |
| DLD | ATP5MG |
| DLG3 | ATP5MGL |
| DLG4 | ATP5PB |
| DMD | ATP5PD |
| DMPK | ATP5PF |
| DNAJC12 | ATP5PO |
| DNAJC19 | ATP6AP1 |
| DNM1 | ATP6AP2 |
| DNMT3A | ATP6V0A2 |
| DNMT3B | ATP6V0A4 |
| DOCK6 | ATP6V1A |
| DOCK7 | ATP6V1B1 |
| DOCK8 | ATP6V1B2 |
| DOLK | ATP7A |
| DONSON | ATP7B |
| DPAGT1 | ATP8A2 |
| DPF2 | ATP8B1 |
| DPH1 | ATPAF1 |
| DPM1 | ATPAF2 |
| DPP6 | ATR |
| DPYD | ATRX |
| DPYS | ATXN1 |
| DST | ATXN10 |
| DYM | ATXN2 |
| DYNC1H1 | ATXN3 |
| DYRK1A | ATXN7 |
| EBF3 | ATXN8OS |
| EBP | AUH |
| ECHS1 | AURKA |
| EDC3 | AURKC |
| EED | AUTS2 |
| EEF1A2 | AVP |
| EFNB2 | AVPR2 |
| EFTUD2 | AXIN1 |
| EHMT1 | AXL |
| EIF2AK3 | B2M |
| EIF2S3 | B3GALNT1 |
| EIF3F | B3GALNT2 |
| EIF4A3 | B3GALT6 |
| EIF4G1 | B3GAT3 |
| ELAC2 | B3GLCT |
| ELOVL4 | B4GALNT1 |

ELP2
EMC1
EML1
EMX2
ENTPD1
EP300
EPB41L1
EPG5
ERCC1
ERCC2
ERCC3
ERCC5
ERCC6
ERCC8
ERLIN2
ESCO2
ETFA
ETFB
ETFDH
ETHE1
EXOC8
EXOSC2
EXOSC3
EXOSC9
EXTL3
EZH2
FA2H
FAM126A
FAM20C
FANCD2
FAR1
FARSB
FAT4
FBN1
FBXL3
FBXL4
FBXO11
FBXO31
FGD1
FGF12
FGF14
FGFR1
FGFR2
FGFR3
FH
FIBP
FIGN
FKRP
B4GALT1
B4GALT7
B4GAT1
B9D1
B9D2
BAAT
BACH2
BAG3
BANF1
BARD1
BAX
BBIP1
BBS1
BBS10
BBS12
BBS2
BBS4
BBS5
BBS7
BBS9
BCAM
BCAP31
BCHE
BCKDHA
BCKDHB
BCKDK
BCL10
BCL11A
BCL11B
BCL2
BCL7A
BCL9
BCO1
BCOR
BCORL1
BCR
BCS1L
BDNF
BEAN1
BEST1
BFSP1
BFSP2
BGN
BHLHA9
BHLHE41
BICC1
BICD2
BIN1

| | |
|---|---|
| FKTN | BLK |
| FLNA | BLM |
| FLVCR1 | BLNK |
| FLVCR2 | BLOC1S3 |
| FMN2 | BLOC1S6 |
| FMR1 | BLVRA |
| FOLR1 | BMP1 |
| FOXG1 | BMP15 |
| FOXP1 | BMP2 |
| FOXP2 | BMP4 |
| FOXRED1 | BMPER |
| FRAS1 | BMPR1B |
| FREM2 | BMPR2 |
| FRMD4A | BMS1 |
| FRMPD4 | BNC2 |
| FRRS1L | BOLA1 |
| FTCD | BOLA2 |
| FTO | BOLA3 |
| FTSJ1 | BPGM |
| FUCA1 | BPTF |
| FUT8 | BRAF |
| GABBR2 | BRAT1 |
| GABRA1 | BRCA2 |
| GABRA3 | BRCC3 |
| GABRB1 | BRD4 |
| GABRB2 | BRF1 |
| GABRB3 | BRPF1 |
| GABRG2 | BRSK2 |
| GAD1 | BRWD3 |
| GALC | BSCL2 |
| GALE | BSG |
| GALT | BSND |
| GAMT | BTD |
| GATAD2B | BTK |
| GATM | BTLA |
| GBA | BTNL2 |
| GCDH | BUB1 |
| GCH1 | BUB1B |
| GCSH | BVES |
| GDI1 | C12orf4 |
| GFAP | C12orf57 |
| GFM1 | C12orf65 |
| GFM2 | C15orf41 |
| GJA1 | C19orf12 |
| GJB1 | C1GALT1C1 |
| GJC2 | C1QA |
| GK | C1QB |
| GLB1 | C1QBP |

GLDC
GLI2
GLI3
GLIS3
GLUD1
GLYCTK
GM2A
GMPPA
GMPPB
GNAO1
GNAS
GNB1
GNB5
GNPAT
GNPTAB
GNPTG
GNS
GPAA1
GPC3
GPHN
GPSM2
GPT2
GRIA3
GRIA4
GRID2
GRIK2
GRIN1
GRIN2A
GRIN2B
GRIN2D
GRIN3B
GRIP1
GRM1
GRN
GSE1
GSS
GTF2H5
GTPBP2
GTPBP3
GUSB
HACE1
HADH
HADHA
HAX1
HCCS
HCFC1
HCN1
HDAC4
C1QC
C1QTNF5
C1R
C1S
C2
C2CD3
C3
C4A
C4B
C5
C6
C7
C8A
C8B
C8G
C8orf37
C9
C9orf72
CA12
CA2
CA4
CA5A
CA8
CABP2
CABP4
CACNA1A
CACNA1B
CACNA1C
CACNA1D
CACNA1E
CACNA1F
CACNA1G
CACNA1H
CACNA1S
CACNA2D1
CACNA2D2
CACNA2D4
CACNB2
CACNB4
CACNG2
CAD
CALCR
CALM1
CALM2
CALM3
CALR
CALR3
CAMK2A

| | |
|---|---|
| HDAC6 | CAMK2B |
| HDAC8 | CAMTA1 |
| HECTD1 | CANT1 |
| HECW2 | CAP2 |
| HEPACAM | CAPN1 |
| HERC1 | CAPN10 |
| HERC2 | CAPN12 |
| HESX1 | CAPN3 |
| HEXA | CAPN5 |
| HEXB | CARD11 |
| HGSNAT | CARD14 |
| HIBCH | CARD9 |
| HIST1H1E | CARMIL2 |
| HIST1H4C | CARS2 |
| HIVEP2 | CARTPT |
| HLCS | CASK |
| HMGCL | CASP10 |
| HMGCS2 | CASP12 |
| HNMT | CASP14 |
| HNRNPH2 | CASP8 |
| HNRNPK | CASQ1 |
| HNRNPU | CASQ2 |
| HOXA1 | CASR |
| HPD | CAST |
| HPRT1 | CAT |
| HRAS | CATSPER1 |
| HSD17B10 | CAV1 |
| HSD17B4 | CAV3 |
| HSPA9 | CAVIN1 |
| HSPD1 | CAVIN4 |
| HTRA2 | CBL |
| HUWE1 | CBLIF |
| HYLS1 | CBS |
| IARS | CBX2 |
| IARS2 | CC2D1A |
| IDS | CC2D2A |
| IDUA | CCBE1 |
| IER3IP1 | CCDC103 |
| IFIH1 | CCDC114 |
| IFT172 | CCDC115 |
| IFT81 | CCDC137 |
| IGBP1 | CCDC14 |
| IGF1 | CCDC151 |
| IGF1R | CCDC174 |
| IKBKG | CCDC22 |
| IL1RAPL1 | CCDC28B |
| IMPA1 | CCDC39 |
| INPP5E | CCDC40 |

| | |
|---|---|
| INPP5K | CCDC50 |
| IQSEC2 | CCDC65 |
| IRF2BPL | CCDC78 |
| ISCA2 | CCDC8 |
| ISPD | CCDC88A |
| ITGA7 | CCDC88C |
| ITPA | CCL11 |
| ITPR1 | CCL2 |
| IVD | CCL3L1 |
| JAG1 | CCL5 |
| JAM3 | CCM2 |
| JMJD1C | CCN6 |
| KALRN | CCND1 |
| KANK1 | CCND2 |
| KANSL1 | CCNK |
| KAT6A | CCNO |
| KAT6B | CCNQ |
| KATNB1 | CCR2 |
| KCNA2 | CCR5 |
| KCNA4 | CCT5 |
| KCNB1 | CD151 |
| KCNC1 | CD164 |
| KCNC3 | CD19 |
| KCNH1 | CD207 |
| KCNJ10 | CD209 |
| KCNJ11 | CD244 |
| KCNJ6 | CD247 |
| KCNK9 | CD27 |
| KCNQ2 | CD2AP |
| KCNQ3 | CD320 |
| KCNQ5 | CD36 |
| KCNT1 | CD3D |
| KCTD7 | CD3E |
| KDM1A | CD3G |
| KDM5B | CD4 |
| KDM5C | CD40 |
| KDM6A | CD40LG |
| KIAA0586 | CD44 |
| KIAA1109 | CD46 |
| KIDINS220 | CD55 |
| KIF11 | CD59 |
| KIF14 | CD70 |
| KIF1A | CD79A |
| KIF1BP | CD79B |
| KIF2A | CD81 |
| KIF4A | CD8A |
| KIF5C | CD96 |
| KIF7 | CDAN1 |

| | |
|---|---|
| KIRREL3 | CDC14A |
| KLF7 | CDC42 |
| KLHL15 | CDC45 |
| KMT2A | CDC6 |
| KMT2B | CDC73 |
| KMT2C | CDCA7 |
| KMT2D | CDH11 |
| KMT5B | CDH15 |
| KNL1 | CDH23 |
| KPTN | CDH3 |
| KRAS | CDHR1 |
| KRBOX4 | CDK10 |
| L1CAM | CDK13 |
| L2HGDH | CDK5 |
| LAMA1 | CDK5RAP2 |
| LAMA2 | CDK6 |
| LAMB1 | CDK8 |
| LAMC3 | CDKAL1 |
| LAMP2 | CDKL5 |
| LARGE1 | CDKN1A |
| LARP7 | CDKN1C |
| LAS1L | CDKN2B |
| LGI4 | CDKN2C |
| LIAS | CDON |
| LIG4 | CDSN |
| LINGO1 | CDT1 |
| LINS1 | CEACAM16 |
| LMAN2L | CEBPE |
| LMBRD1 | CEL |
| LONP1 | CELSR2 |
| LRP2 | CENPE |
| LRPPRC | CENPF |
| LZTFL1 | CENPJ |
| LZTR1 | CEP104 |
| MAB21L1 | CEP120 |
| MAB21L2 | CEP135 |
| MACF1 | CEP152 |
| MAF | CEP164 |
| MAG | CEP19 |
| MAGEL2 | CEP290 |
| MAGT1 | CEP41 |
| MAN1B1 | CEP57 |
| MAN2B1 | CEP63 |
| MANBA | CEP78 |
| MAOA | CEP83 |
| MAP1B | CEP89 |
| MAP2K1 | CERKL |
| MAP2K2 | CERS1 |

| | |
|---|---|
| MAPK8IP3 | CERS3 |
| MAPRE2 | CES1 |
| MASP1 | CETP |
| MAT1A | CFAP298 |
| MAU2 | CFAP53 |
| MBD5 | CFAP57 |
| MBOAT7 | CFB |
| MBTPS2 | CFC1 |
| MCCC1 | CFD |
| MCCC2 | CFH |
| MCOLN1 | CFHR1 |
| MCPH1 | CFHR2 |
| MDH2 | CFHR3 |
| MECP2 | CFHR4 |
| MECR | CFHR5 |
| MED12 | CFI |
| MED13 | CFL2 |
| MED13L | CFP |
| MED17 | CFTR |
| MED23 | CHAMP1 |
| MED25 | CHAT |
| MEF2C | CHCHD10 |
| MEGF8 | CHCHD2 |
| MEIS2 | CHD1 |
| METTL23 | CHD2 |
| MFF | CHD3 |
| MFSD2A | CHD4 |
| MFSD8 | CHD7 |
| MGAT2 | CHD8 |
| MGP | CHI3L1 |
| MICU1 | CHIC2 |
| MID1 | CHIT |
| MID2 | CHIT1 |
| MKKS | CHKB |
| MKS1 | CHM |
| MLC1 | CHMP1A |
| MLYCD | CHMP2B |
| MMAA | CHMP4B |
| MMAB | CHN1 |
| MMACHC | CHRDL1 |
| MMADHC | CHRM2 |
| MMUT | CHRM3 |
| MOCS1 | CHRNA1 |
| MOCS2 | CHRNA2 |
| MOGS | CHRNA3 |
| MPDU1 | CHRNA4 |
| MPDZ | CHRNA5 |
| MPLKIP | CHRNB1 |

| | |
|---|---|
| MRPL3 | CHRNB2 |
| MRPS22 | CHRND |
| MSL3 | CHRNE |
| MSMO1 | CHRNG |
| MTFMT | CHST14 |
| MTHFR | CHST3 |
| MTOR | CHST6 |
| MTR | CHST8 |
| MTRR | CHSY1 |
| MVK | CHUK |
| MYCN | CIB2 |
| MYH9 | CIC |
| MYO5A | CIDEC |
| MYT1L | CIITA |
| NAA10 | CILP |
| NAA15 | CISD2 |
| NACC1 | CISH |
| NAGA | CIT |
| NAGLU | CITED2 |
| NAGS | CIZ1 |
| NALCN | CKAP2L |
| NANS | CLCF1 |
| NARS2 | CLCN1 |
| NBEA | CLCN2 |
| NBN | CLCN4 |
| NDE1 | CLCN5 |
| NDP | CLCN7 |
| NDST1 | CLCNKA |
| NDUFA1 | CLCNKB |
| NDUFA11 | CLDN1 |
| NDUFA12 | CLDN10 |
| NDUFA2 | CLDN14 |
| NDUFAF3 | CLDN16 |
| NDUFAF5 | CLDN19 |
| NDUFS1 | CLEC4D |
| NDUFS2 | CLEC7A |
| NDUFS3 | CLIC2 |
| NDUFS4 | CLIC5 |
| NDUFS6 | CLIP1 |
| NDUFS7 | CLMP |
| NDUFS8 | CLN3 |
| NDUFV1 | CLN5 |
| NDUFV2 | CLN6 |
| NECAP1 | CLN8 |
| NECTIN1 | CLP1 |
| NEDD4L | CLPB |
| NEU1 | CLPP |
| NEXMIF | CLRN1 |

NF1
NFATC1
NFE2L2
NFIA
NFIX
NFU1
NGLY1
NHS
NIPBL
NKX2-1
NLGN3
NLGN4X
NLRP3
NONO
NPC1
NPC2
NPHP1
NR2F1
NR4A2
NR5A1
NRAS
NRXN1
NSD1
NSD2
NSDHL
NSUN2
NT5C2
NT5C3A
NTRK1
NTRK2
NUBPL
NUP62
NUS1
OAT
OCLN
OCRL
ODC1
OFD1
OGT
OPHN1
ORC1
OSGEP
OTC
OTUD6B
OTX2
OXCT1
P4HTM
PACS1
CLTC
CNBP
CNGA1
CNGA3
CNGB1
CNGB3
CNKSR2
CNNM2
CNNM4
CNOT1
CNOT3
CNPY3
CNTN1
CNTN2
CNTNAP1
CNTNAP2
COA1
COA3
COA5
COA6
COA7
COASY
COCH
COG1
COG4
COG5
COG6
COG7
COG8
COL10A1
COL11A1
COL11A2
COL12A1
COL13A1
COL14A1
COL17A1
COL18A1
COL1A1
COL1A2
COL25A1
COL27A1
COL2A1
COL3A1
COL4A1
COL4A2
COL4A3
COL4A3BP
COL4A4

PACS2
PAFAH1B1
PAH
PAK3
PANK2
PANX1
PARN
PAX1
PAX6
PAX8
PBX1
PC
PCBD1
PCCA
PCCB
PCDH19
PCGF2
PCLO
PCNT
PDE4D
PDHA1
PDHX
PDP1
PDSS1
PDSS2
PEPD
PET100
PEX1
PEX10
PEX11B
PEX12
PEX13
PEX16
PEX19
PEX2
PEX26
PEX3
PEX5
PEX6
PEX7
PGAP1
PGAP2
PGAP3
PGK1
PGM3
PHF21A
PHF6
PHF8
COL4A5
COL4A6
COL5A1
COL5A2
COL6A1
COL6A2
COL6A3
COL7A1
COL8A2
COL9A1
COL9A2
COL9A3
COLEC11
COLQ
COMP
COMT
COPA
COQ2
COQ4
COQ5
COQ6
COQ7
COQ8A
COQ8B
COQ9
CORIN
CORO1A
COX10
COX14
COX15
COX20
COX4I1
COX4I2
COX5A
COX5B
COX6A1
COX6A2
COX6B1
COX6B2
COX6C
COX7A1
COX7A2
COX7B
COX7B2
COX7C
COX8A
COX8C
CP

PHGDH
PHIP
PI4KA
PIGA
PIGC
PIGG
PIGL
PIGN
PIGO
PIGT
PIGV
PIGW
PIGY
PIK3CA
PIK3R2
PLA2G6
PLAA
PLCB1
PLK4
PLP1
PLPBP
PLXND1
PMM2
PMPCA
PMPCB
PNKP
PNP
PNPLA6
POC1A
POGZ
POLG
POLR3A
POLR3B
POMGNT1
POMGNT2
POMK
POMT1
POMT2
PORCN
POU1F1
POU3F3
PPM1D
PPOX
PPP1CB
PPP1R15B
PPP2CA
PPP2R1A
PPP2R5B
CPA6
CPAMD8
CPLANE1
CPLX1
CPN1
CPOX
CPS1
CPT1A
CPT1C
CPT2
CPZ
CR1
CR2
CRADD
CRB1
CRB2
CRBN
CREB1
CREB3L1
CREBBP
CRELD1
CRIPT
CRLF1
CRTAP
CRTC1
CRX
CRYAA
CRYAB
CRYBA1
CRYBA2
CRYBA4
CRYBB1
CRYBB2
CRYBB3
CRYGB
CRYGC
CRYGD
CRYGS
CRYM
CSF1R
CSF2RA
CSF2RB
CSF3R
CSMF
CSNK1D
CSNK2A1
CSNK2B
CSPP1

| | |
|---|---|
| PPP2R5C | CSRP3 |
| PPP2R5D | CST3 |
| PPP3CA | CSTA |
| PPT1 | CSTB |
| PQBP1 | CTBP1 |
| PRF1 | CTC1 |
| PRKAR1A | CTCF |
| PRMT7 | CTDP1 |
| PRODH | CTF1 |
| PRPS1 | CTH |
| PRR12 | CTLA4 |
| PRRT2 | CTNNA2 |
| PRSS12 | CTNNA3 |
| PRUNE1 | CTNNB1 |
| PSAP | CTNND1 |
| PSAT1 | CTNND2 |
| PSEN1 | CTNS |
| PSMD12 | CTPS1 |
| PSPH | CTRC |
| PTCH1 | CTSA |
| PTCHD1 | CTSC |
| PTDSS1 | CTSD |
| PTEN | CTSF |
| PTF1A | CTSK |
| PTPN11 | CTTNBP2 |
| PTRH2 | CUBN |
| PTRHD1 | CUL3 |
| PTS | CUL4B |
| PUF60 | CUL7 |
| PUM1 | CUX1 |
| PURA | CUX2 |
| PUS1 | CWC27 |
| PUS3 | CWF19L1 |
| PUS7 | CX3CR1 |
| PYCR1 | CXCL12 |
| PYCR2 | CXCR1 |
| QARS | CXCR4 |
| QDPR | CXorf56 |
| QRICH1 | CYB5A |
| RAB11B | CYB5R3 |
| RAB18 | CYBA |
| RAB23 | CYBB |
| RAB27A | CYBC1 |
| RAB39B | CYC1 |
| RAB3GAP1 | CYCS |
| RAB3GAP2 | CYLD |
| RAB40AL | CYP11A1 |
| RAC1 | CYP11B1 |

RAD21
RAF1
RAI1
RALA
RARB
RARS2
RBBP8
RBFOX1
RBM10
RBM28
RBPJ
RCBTB1
RECQL4
RELN
RERE
REV3L
RFT1
RHEB
RHOBTB2
RIT1
RLIM
RMND1
RNASEH2A
RNASEH2B
RNASEH2C
RNASET2
RNF113A
RNF125
ROGDI
ROR2
RORA
RPGRIP1L
RPL10
RPS19
RPS6KA3
RRM2B
RSPRY1
RSRC1
RTEL1
RTN4IP1
RTTN
RUBCN
RUSC2
RXYLT1
SALL1
SAMD9
SAMHD1
SARS
CYP11B2
CYP17A1
CYP19A1
CYP1A2
CYP1B1
CYP21A2
CYP24A1
CYP26B1
CYP26C1
CYP27A1
CYP27B1
CYP2A6
CYP2B6
CYP2C
CYP2C19
CYP2C8
CYP2C9
CYP2D6
CYP2R1
CYP2U1
CYP3A4
CYP3A5
CYP4F2
CYP4F22
CYP4V2
CYP7A1
CYP7B1
CYTB
D2HGDH
DAG1
DAND5
DAPK3
DARS
DARS2
DAX1
DAZL
DBET
DBH
DBT
DCAF17
DCAF8
DCC
DCDC2
DCHS1
DCLRE1B
DCLRE1C
DCN
DCPS

SATB2
SBDS
SC5D
SCAPER
SCN1A
SCN1B
SCN2A
SCN3A
SCN8A
SCO1
SCO2
SCYL1
SDCCAG8
SDHA
SEMA3E
SEPSECS
SERAC1
SET
SETBP1
SETD1A
SETD1B
SETD2
SETD5
SF1
SGPL1
SGSH
SHANK2
SHANK3
SHH
SHOC2
SHROOM4
SIK1
SIL1
SIN3A
SIX3
SKI
SLC12A5
SLC12A6
SLC13A5
SLC16A2
SLC17A5
SLC19A3
SLC1A1
SLC1A2
SLC1A4
SLC25A1
SLC25A12
SLC25A13

DCTN1
DCX
DCXR
DDB2
DDC
DDHD1
DDHD2
DDOST
DDR2
DDX11
DDX3X
DDX41
DDX58
DDX59
DEAF1
DECR1
DENND5A
DEPDC5
DES
DGAT1
DGCR2
DGKE
DGUOK
DHCR24
DHCR7
DHDDS
DHFR
DHH
DHODH
DHTKD1
DHX30
DIABLO
DIAPH1
DIAPH2
DIAPH3
DIP2B
DIS3L2
DISC1
DJ1
DKC1
DLAT
DLC1
DLD
DLG3
DLG4
DLL1
DLL3
DLL4

| | |
|---|---|
| SLC25A15 | DLST |
| SLC25A22 | DLX3 |
| SLC25A24 | DLX4 |
| SLC2A1 | DLX5 |
| SLC33A1 | DMAC1 |
| SLC35A1 | DMAC2 |
| SLC35A2 | DMAC2L |
| SLC35A3 | DMD |
| SLC35C1 | DMGDH |
| SLC39A12 | DMP1 |
| SLC39A14 | DMPK |
| SLC39A8 | DMXL2 |
| SLC46A1 | DNA2 |
| SLC4A4 | DNAAF1 |
| SLC52A2 | DNAAF2 |
| SLC6A1 | DNAAF3 |
| SLC6A17 | DNAAF4 |
| SLC6A19 | DNAAF5 |
| SLC6A3 | DNAH1 |
| SLC6A8 | DNAH11 |
| SLC6A9 | DNAH5 |
| SLC7A7 | DNAH6 |
| SLC9A6 | DNAI1 |
| SLC9A7 | DNAI2 |
| SMAD4 | DNAJB13 |
| SMAD6 | DNAJB2 |
| SMARCA1 | DNAJB6 |
| SMARCA2 | DNAJC12 |
| SMARCA4 | DNAJC13 |
| SMARCB1 | DNAJC19 |
| SMARCC2 | DNAJC21 |
| SMARCE1 | DNAJC3 |
| SMC1A | DNAJC5 |
| SMC3 | DNAJC6 |
| SMG9 | DNAL1 |
| SMOC1 | DNAL4 |
| SMPD1 | DNASE1 |
| SMS | DNASE1L3 |
| SNAP25 | DNM1 |
| SNAP29 | DNM1L |
| SNIP1 | DNM2 |
| SNRPB | DNMT1 |
| SNRPN | DNMT3A |
| SNX14 | DNMT3B |
| SOBP | DOCK2 |
| SON | DOCK6 |
| SOS1 | DOCK7 |
| SOS2 | DOCK8 |

| | |
|---|---|
| SOX10 | DOK7 |
| SOX11 | DOLK |
| SOX2 | DONSON |
| SOX3 | DPAGT1 |
| SOX4 | DPF2 |
| SOX5 | DPH1 |
| SPART | DPM1 |
| SPAST | DPM2 |
| SPATA5 | DPM3 |
| SPECC1L | DPP6 |
| SPG11 | DPY19L2 |
| SPOCK1 | DPYD |
| SPR | DPYS |
| SPRED1 | DRAM2 |
| SPTAN1 | DRC1 |
| SPTBN2 | DRD2 |
| SRCAP | DRD3 |
| SRD5A3 | DRD4 |
| SRPX2 | DRD5 |
| SSR4 | DSC1 |
| ST3GAL3 | DSC2 |
| ST3GAL5 | DSC3 |
| STAG1 | DSE |
| STAMBP | DSG1 |
| STIL | DSG2 |
| STRA6 | DSG3 |
| STRADA | DSG4 |
| STT3A | DSP |
| STT3B | DSPP |
| STX1B | DST |
| STXBP1 | DSTYK |
| SUCLA2 | DTHD1 |
| SUCLG1 | DTNA |
| SUMF1 | DTNBP1 |
| SUOX | DUOX2 |
| SURF1 | DUOXA2 |
| SUZ12 | DUSP6 |
| SVBP | DVL1 |
| SYN1 | DVL3 |
| SYNCRIP | DYM |
| SYNE1 | DYNC1H1 |
| SYNGAP1 | DYNC2H1 |
| SYNJ1 | DYNC2LI1 |
| SYP | DYRK1A |
| SYT1 | DYRK1B |
| SYT14 | DYSF |
| SZT2 | EARS2 |
| TAF1 | EBF3 |

| | |
|---|---|
| TAF13 | EBP |
| TAF2 | ECE1 |
| TAF6 | ECEL1 |
| TANC2 | ECHS1 |
| TANGO2 | ECM1 |
| TAT | ECSIT |
| TBC1D20 | EDA |
| TBC1D23 | EDA1 |
| TBC1D24 | EDAR |
| TBC1D7 | EDARADD |
| TBCD | EDC3 |
| TBCE | EDN1 |
| TBCK | EDN3 |
| TBL1XR1 | EDNRA |
| TBP | EDNRB |
| TBR1 | EED |
| TBX1 | EEF1A2 |
| TCF20 | EEF2 |
| TCF4 | EFEMP1 |
| TCF7L2 | EFEMP2 |
| TCN2 | EFHC1 |
| TCTN2 | EFNB1 |
| TCTN3 | EFNB2 |
| TDP2 | EFTUD2 |
| TECPR2 | EGF |
| TECR | EGLN1 |
| TELO2 | EGR2 |
| TFAP2A | EHBP1 |
| TGDS | EHHADH |
| TGFBR1 | EHMT1 |
| TGFBR2 | EIF2AK3 |
| TGIF1 | EIF2AK4 |
| TH | EIF2B1 |
| THOC2 | EIF2B2 |
| THOC6 | EIF2B3 |
| THRB | EIF2B4 |
| TIMM50 | EIF2B5 |
| TIMM8A | EIF2S3 |
| TINF2 | EIF3F |
| TKT | EIF4A3 |
| TLK2 | EIF4E |
| TMCO1 | EIF4G1 |
| TMEM165 | ELAC2 |
| TMEM216 | ELANE |
| TMEM231 | ELF4 |
| TMEM237 | ELMO2 |
| TMEM240 | ELMOD3 |
| TMEM67 | ELN |

| | |
|---|---|
| TMEM70 | ELOVL4 |
| TMLHE | ELOVL5 |
| TMTC3 | ELP1 |
| TNIK | ELP2 |
| TOE1 | ELP4 |
| TP53RK | EMC1 |
| TPI1 | EMD |
| TPO | EMG1 |
| TPP1 | EML1 |
| TPRKB | EMP2 |
| TRAF7 | EMX2 |
| TRAIP | ENAM |
| TRAPPC11 | ENG |
| TRAPPC6B | ENO3 |
| TRAPPC9 | ENPP1 |
| TREX1 | ENTPD1 |
| TRIM32 | EOGT |
| TRIO | EP300 |
| TRIP12 | EPAS1 |
| TRIP4 | EPB41 |
| TRIT1 | EPB41L1 |
| TRMT1 | EPB42 |
| TRMT10A | EPCAM |
| TRNT1 | EPG5 |
| TRRAP | EPHA2 |
| TSC1 | EPHB2 |
| TSC2 | EPHB4 |
| TSEN15 | EPHX1 |
| TSEN2 | EPHX2 |
| TSEN54 | EPM2A |
| TSFM | EPO |
| TSHB | EPOR |
| TSPAN7 | EPS8 |
| TTC19 | EPX |
| TTC37 | ERAL1 |
| TTC8 | ERBB2 |
| TTI2 | ERBB3 |
| TUBA1A | ERBB4 |
| TUBA8 | ERCC1 |
| TUBB | ERCC2 |
| TUBB2A | ERCC3 |
| TUBB2B | ERCC4 |
| TUBB3 | ERCC5 |
| TUBB4A | ERCC6 |
| TUBG1 | ERCC6L2 |
| TUBGCP4 | ERCC8 |
| TUBGCP6 | ERF |
| TUSC3 | ERLIN1 |

TWIST1
TWNK
UBA5
UBE2A
UBE3A
UBE3B
UBR1
UBTF
UFC1
UFM1
UNC13A
UNC80
UPB1
UPF3B
UQCRQ
UROC1
USP27X
USP7
USP9X
VAMP1
VAMP2
VLDLR
VPS11
VPS13B
VPS37A
VPS53
VRK1
VWA3B
WAC
WARS2
WASF1
WASHC4
WDPCP
WDR13
WDR19
WDR26
WDR4
WDR45
WDR45B
WDR62
WDR73
WDR81
WFS1
WWOX
XPA
XPNPEP3
XRCC4
XYLT1
ERLIN2
ERMAP
ERMARD
ESAM
ESCO2
ESPN
ESR1
ESRRB
ETFA
ETFB
ETFDH
ETHE1
ETV6
EVC
EVC2
EWSR1
EXOC8
EXOSC2
EXOSC3
EXOSC8
EXOSC9
EXPH5
EXT1
EXT2
EXTL3
EYA1
EYA4
EYS
EZH2
F10
F11
F12
F13A1
F13B
F2
F5
F7
F8
F9
FA2H
FAAH
FAAH2
FAAP24
FAAP95
FADD
FAH
FAM111A
FAM111B

XYLT2
YAP1
YME1L1
YWHAE
YWHAG
YY1
ZBTB11
ZBTB16
ZBTB18
ZBTB20
ZBTB24
ZC3H14
ZC4H2
ZDHHC15
ZDHHC9
ZEB2
ZFYVE26
ZIC1
ZIC2
ZMIZ1
ZMYND11
ZNF148
ZNF292
ZNF407
ZNF41
ZNF462
ZNF592
ZNF674
ZNF711
ZNF81
ZSWIM6

FAM126A
FAM161A
FAM20A
FAM20C
FAM83G
FAM83H
FANCA
FANCB
FANCC
FANCD2
FANCE
FANCF
FANCG
FANCI
FANCL
FANCM
FAR1
FARS2
FARSB
FAS
FASLG
FASTKD2
FAT2
FAT4
FBLN1
FBLN5
FBN1
FBN2
FBP1
FBXL3
FBXL4
FBXO11
FBXO31
FBXO38
FBXO7
FCGR1A
FCGR2A
FCGR2B
FCGR3A
FCGR3B
FCGRT
FCN3
FCYT
FDPS
FDX2
FDXR
FECH
FERMT1

FERMT3
FEZF1
FFAR4
FGA
FGB
FGD1
FGD4
FGF10
FGF12
FGF14
FGF16
FGF17
FGF20
FGF23
FGF3
FGF5
FGF8
FGF9
FGFR1
FGFR2
FGFR3
FGFR4
FGG
FHL1
FHL2
FIBP
FIG4
FIGLA
FIGN
FKBP10
FKBP14
FKBP5
FKBPL
FKRP
FKTN
FLAD1
FLG
FLG2
FLI1
FLJ22792
FLNA
FLNB
FLNC
FLRT3
FLT3
FLT4
FLVCR1
FLVCR2

FMN2
FMO3
FMR1
FN1
FOLR1
FOS
FOXC1
FOXC2
FOXD3
FOXD4
FOXE1
FOXE3
FOXF1
FOXF2
FOXG1
FOXH1
FOXI1
FOXL1
FOXL2
FOXN1
FOXO1
FOXP1
FOXP2
FOXP3
FOXRED1
FPR1
FRAS1
FREM1
FREM2
FRMD4A
FRMD7
FRMPD4
FRRS1L
FRZB
FSCN2
FSHB
FSHR
FTCD
FTH1
FTL
FTO
FTSJ1
FUCA1
FUS
FUT1
FUT2
FUT3
FUT6

FUT8
FUZ
FXN
FXYD2
FYCO1
FZD4
FZD6
G6PC
G6PC3
G6PD
GAA
GABBR2
GABRA1
GABRA2
GABRA3
GABRB1
GABRB2
GABRB3
GABRD
GABRG2
GAD1
GAL
GALC
GALE
GALK1
GALNS
GALNT11
GALNT12
GALNT3
GALT
GAMT
GAN
GANAB
GARS
GAS1
GAS8
GATA1
GATA3
GATA4
GATA5
GATA6
GATAD1
GATAD2B
GATB
GATC
GATM
GBA
GBA2

GBE1
GCDH
GCGR
GCH1
GCK
GCKR
GCLC
GCLM
GCM2
GCNT2
GCSH
GDAP1
GDAP2
GDF1
GDF2
GDF3
GDF5
GDF6
GDI1
GDNF
GFAP
GFER
GFI1
GFI1B
GFM1
GFM2
GFPT1
GGCX
GH1
GHR
GHRHR
GHRL
GHSR
GIGYF2
GINS1
GIPC3
GJA1
GJA3
GJA5
GJA8
GJB1
GJB2
GJB3
GJB4
GJB6
GJC2
GK
GLA

GLB1
GLCCI1
GLDC
GLDN
GLE1
GLI2
GLI3
GLIS2
GLIS3
GLML
GLMN
GLRA1
GLRB
GLRX5
GLUD1
GLUD2
GLUL
GLYCTK
GM2A
GMNN
GMPPA
GMPPB
GNA11
GNAI2
GNAI3
GNAL
GNAO1
GNAQ
GNAS
GNAS-AS1
GNAT1
GNAT2
GNB1
GNB3
GNB4
GNB5
GNE
GNMT
GNPAT
GNPTAB
GNPTG
GNRH1
GNRHR
GNS
GORAB
GOSR2
GOT1
GP1BA

GP1BB
GP6
GP9
GPAA1
GPC3
GPC4
GPC6
GPD1
GPD1L
GPD2
GPHN
GPI
GPIHBP1
GPR101
GPR143
GPR161
GPR179
GPR68
GPR88
GPSM2
GPT2
GPX1
GPX4
GREM1
GREM2
GRHL2
GRHL3
GRHPR
GRIA3
GRIA4
GRID2
GRIK2
GRIK4
GRIN1
GRIN2A
GRIN2B
GRIN2D
GRIN3B
GRIP1
GRK1
GRM1
GRM6
GRN
GRXCR1
GRXCR2
GSBS
GSC
GSDME

GSE1
GSN
GSS
GTF2E2
GTF2H5
GTPBP2
GTPBP3
GUCA1A
GUCA1B
GUCY1A1
GUCY2C
GUCY2D
GUF1
GUSB
GYG1
GYPA
GYPB
GYPC
GYS1
GYS2
H19
H6PD
HAAO
HACE1
HADH
HADHA
HADHB
HAL
HAMP
HAND1
HAND2
HARS
HARS2
HAS2
HAVCR1
HAX1
HBA1
HBA2
HBB
HBD
HBG1
HBG2
HCCS
HCFC1
HCN1
HCN4
HCRT
HDAC4

HDAC6
HDAC8
HDC
HECTD1
HECW2
HELLS
HEPACAM
HERC1
HERC2
HES7
HESX1
HEXA
HEXB
HEY2
HFE
HFM1
HGD
HGF
HGSNAT
HIBCH
HIKESHI
HINT1
HIST1H1E
HIST1H4C
HIVEP2
HJV
HK1
HLA-A
HLA-B
HLA-C
HLA-DPB1
HLA-DQA1
HLA-DQB1
HLA-DRB1
HLCS
HMBS
HMCN1
HMGA1
HMGB3
HMGCL
HMGCR
HMGCS2
HMMR
HMOX1
HMX1
HNF1A
HNF1B
HNF4A

HNMT
HNRNPA1
HNRNPA2B1
HNRNPDL
HNRNPH2
HNRNPK
HNRNPU
HOGA1
HOMER2
HOMEZ
HOXA1
HOXA11
HOXA13
HOXA2
HOXB1
HOXB13
HOXC13
HOXD10
HOXD13
HP
HPCA
HPD
HPGD
HPRT1
HPS1
HPS3
HPS4
HPS5
HPS6
HPSE2
HR
HRAS
HRG
HS6ST1
HSD11B1
HSD11B2
HSD17B10
HSD17B3
HSD17B4
HSD3B2
HSD3B7
HSF4
HSPA1L
HSPA9
HSPB1
HSPB3
HSPB6
HSPB8

HSPD1
HSPG2
HTR1A
HTR2A
HTRA1
HTRA2
HTT
HUWE1
HYAL1
HYDIN
HYLS1
HYOU1
IARS
IARS2
IBA57
ICAM1
ICAM4
ICK
ICOS
IDH1
IDH2
IDH3B
IDS
IDUA
IER3IP1
IFIH1
IFITM3
IFITM5
IFNAR2
IFNG
IFNGR1
IFNGR2
IFNL3
IFRD1
IFT122
IFT140
IFT172
IFT27
IFT43
IFT52
IFT74
IFT80
IFT81
IFT88
IGBP1
IGF1
IGF1R
IGF2

IGF2BP2
IGF2R
IGFALS
IGFBP7
IGHMBP2
IGLL1
IGSF1
IGSF3
IHH
IKBKB
IKBKG
IKZF1
IL10
IL10RA
IL10RB
IL11RA
IL12B
IL12RB1
IL13
IL17F
IL17RA
IL17RC
IL17RD
IL18
IL1B
IL1RAPL1
IL1RN
IL2
IL21
IL21R
IL23R
IL2RA
IL2RG
IL31RA
IL36RN
IL4R
IL6
IL6R
IL6ST
IL7R
ILDR1
ILK
IMPA1
IMPAD1
IMPDH1
IMPDH2
IMPG1
IMPG2

INF2
ING1
INO80
INPP5D
INPP5E
INPP5K
INPPL1
INS
INSL3
INSR
INVS
IPMK
IQCB1
IQSEC2
IRAK1
IRAK3
IRAK4
IRF1
IRF2BP2
IRF2BPL
IRF3
IRF4
IRF5
IRF6
IRF7
IRF8
IRGM
IRS1
IRS2
IRX1
IRX5
ISCA2
ISCU
ISG15
ISL1
ISPD
ITCH
ITGA2B
ITGA3
ITGA6
ITGA7
ITGA8
ITGAM
ITGB1BP2
ITGB2
ITGB3
ITGB4
ITGB6

ITIH4
ITK
ITM2B
ITPA
ITPKC
ITPR1
ITPR2
IVD
IYD
JAG1
JAGN1
JAK1
JAK2
JAK3
JAM3
JARID2
JMJD1C
JPH1
JPH2
JPH3
JUP
KALRN
KANK1
KANK2
KANSL1
KARS
KAT6A
KAT6B
KATNB1
KBTBD13
KCNA1
KCNA2
KCNA4
KCNA5
KCNB1
KCNC1
KCNC3
KCND3
KCNE1
KCNE2
KCNE3
KCNE5
KCNH1
KCNH2
KCNJ1
KCNJ10
KCNJ11
KCNJ13

KCNJ18
KCNJ2
KCNJ3
KCNJ5
KCNJ6
KCNJ8
KCNK18
KCNK3
KCNK9
KCNMA1
KCNMB1
KCNN4
KCNQ1
KCNQ1OT1
KCNQ2
KCNQ3
KCNQ4
KCNQ5
KCNT1
KCNV2
KCTD1
KCTD17
KCTD7
KDF1
KDM1A
KDM5B
KDM5C
KDM6A
KDR
KDSR
KEL
KERA
KHDC3L
KHK
KIAA0319
KIAA0442
KIAA0556
KIAA0586
KIAA0753
KIAA1109
KIDINS220
KIF11
KIF14
KIF1A
KIF1B
KIF1BP
KIF1C
KIF20A

KIF21A
KIF22
KIF26B
KIF2A
KIF4A
KIF5A
KIF5C
KIF7
KIND1
KIND3
KIR3DL1
KIRREL3
KISS1
KISS1R
KITLG
KIZ
KL
KLC2
KLF1
KLF11
KLF6
KLF7
KLHDC8B
KLHL10
KLHL15
KLHL24
KLHL3
KLHL40
KLHL41
KLHL7
KLK1
KLK4
KLKB1
KLLN
KMT2A
KMT2B
KMT2C
KMT2D
KMT5B
KNG1
KNL1
KPNA7
KPTN
KRAS
KRBOX4
KREMEN1
KRIT1
KRT1

KRT10
KRT12
KRT13
KRT14
KRT16
KRT17
KRT18
KRT2
KRT25
KRT3
KRT4
KRT5
KRT6A
KRT6B
KRT6C
KRT71
KRT74
KRT75
KRT8
KRT81
KRT83
KRT85
KRT86
KRT9
KTU
KY
KYNU
L1CAM
L2HGDH
LACC1
LACTB
LAMA1
LAMA2
LAMA3
LAMA4
LAMB1
LAMB2
LAMB3
LAMC1
LAMC2
LAMC3
LAMP2
LAMTOR2
LARGE1
LARP7
LARS
LARS2
LAS1L

LAT
LBN
LBR
LCA5
LCAT
LCK
LCT
LDB3
LDHA
LDHB
LDLR
LDLRAP1
LEF1
LEFTY1
LEFTY2
LEMD2
LEMD3
LEP
LEPR
LFNG
LGALS2
LGI1
LGI4
LGR4
LHB
LHCGR
LHFPL5
LHX3
LHX4
LIAS
LIFR
LIG1
LIG4
LIM2
LIMS2
LINGO1
LINS1
LIPA
LIPC
LIPE
LIPG
LIPH
LIPI
LIPN
LIPT1
LIPT2
LITAF
LMAN1

LMAN2L
LMBR1
LMBRD1
LMF1
LMNA
LMNB1
LMNB2
LMOD3
LMX1B
LOC387715
LONP1
LOR
LOX
LOXHD1
LOXL1
LPA
LPAR6
LPIN1
LPIN2
LPL
LPP
LRAT
LRBA
LRIG2
LRIT3
LRMDA
LRP1
LRP2
LRP4
LRP5
LRP6
LRP8
LRPAP1
LRPPRC
LRRC6
LRRC8A
LRRK2
LRSAM1
LRTOMT
LSS
LTA
LTBP2
LTBP3
LTBP4
LTC4S
LUM
LYRM4
LYRM7

LYST
LYZ
LZTFL1
LZTR1
LZTS1
MAB21L1
MAB21L2
MACF1
MAD1L1
MAD2L2
MAF
MAFB
MAG
MAGED2
MAGEL2
MAGI2
MAGT1
MAK
MAL
MALT1
MAML1
MAML2
MAMLD1
MAN1B1
MAN2B1
MANBA
MAOA
MAP1B
MAP2K1
MAP2K2
MAP3K1
MAP3K14
MAP3K7
MAP3K8
MAPK1
MAPK10
MAPK8IP1
MAPK8IP3
MAPKAP3
MAPKAPK3
MAPKBP1
MAPRE2
MARS
MARS2
MARVELD2
MASP1
MASP2
MASTL

MAT1A
MAT2A
MATN3
MATN4
MATR3
MAU2
MBD5
MBL2
MBOAT7
MBTPS2
MC1R
MC2R
MC3R
MC4R
MCC
MCCC1
MCCC2
MCEE
MCFD2
MCIDAS
MCM2
MCM4
MCM6
MCM8
MCM9
MCOLN1
MCP
MCPH1
MCT8
MCTP2
MCUR1
MDH2
MDM2
MECOM
MECP2
MECR
MED12
MED13
MED13L
MED17
MED20
MED23
MED25
MEF2A
MEF2C
MEFV
MEGF10
MEGF8

MEIS2
MEN1
MEOX1
MERTK
MESP1
MESP2
METTL23
MFAP5
MFF
MFN2
MFRP
MFSD2A
MFSD8
MGAT2
MGME1
MGP
MIAT
MIB1
MICOS13
MICU1
MICU2
MID1
MID2
MIEF2
MIF
MINPP1
MIP
MIPEP
MIR17HG
MIR184
MIR204
MIR2861
MIR96
MKKS
MKRN3
MKS1
MLC1
MLPH
MLYCD
MMAA
MMAB
MMACHC
MMADHC
MME
MMP1
MMP13
MMP14
MMP19

MMP2
MMP20
MMP21
MMP3
MMP9
MMUT
MN1
MNX1
MOCOS
MOCS1
MOCS2
MOG
MOGS
MORC2
MPC1
MPDU1
MPDZ
MPI
MPL
MPLKIP
MPV17
MPZ
MR1
MRAP
MRAP2
MRE11
MRM2
MRPL12
MRPL3
MRPL40
MRPL44
MRPL57
MRPS14
MRPS16
MRPS2
MRPS22
MRPS23
MRPS34
MRPS7
MRRF
MRTFA
MS4A1
MS4A2
MSL3
MSMB
MSMO1
MSN
MSR1

MSRB3
MST1R
MSTN
MSTO1
MSX1
MSX2
MT-ATP6
MT-ATP8
MT-CO1
MT-CO2
MT-CO3
MT-CYB
MT-ND1
MT-ND2
MT-ND3
MT-ND4
MT-ND4L
MT-ND5
MT-ND6
MT-RNR1
MT-RNR2
MT-TC
MT-TE
MT-TF
MT-TH
MT-TI
MT-TK
MT-TL1
MT-TP
MT-TQ
MT-TS1
MT-TS2
MT-TT
MTAP
MTCP1
MTFMT
MTHFD1
MTHFR
MTM1
MTMR14
MTMR2
MTNR1B
MTO1
MTOR
MTPAP
MTR
MTRR
MTTP

MUC1
MUC5B
MUC7
MUSK
MVD
MVK
MXI1
MYB
MYBPC1
MYBPC3
MYC
MYCN
MYD88
MYF6
MYH10
MYH11
MYH14
MYH2
MYH3
MYH6
MYH7
MYH8
MYH9
MYL2
MYL3
MYL4
MYLIP
MYLK
MYLK2
MYO15A
MYO18B
MYO1A
MYO1C
MYO1E
MYO1F
MYO3A
MYO5A
MYO5B
MYO6
MYO7A
MYO9B
MYOC
MYOM1
MYOT
MYOZ1
MYOZ2
MYPN
MYSM1

MYT1L
NAA10
NAA15
NACC1
NADK2
NAGA
NAGLU
NAGS
NALCN
NANOS1
NANS
NARS2
NAT1
NAT2
NAT8L
NAXE
NBAS
NBEA
NBEAL2
NBN
NCF1
NCF2
NCF4
NCR3
NCSTN
NDE1
NDN
NDP
NDRG1
NDST1
NDUFA1
NDUFA10
NDUFA11
NDUFA12
NDUFA13
NDUFA2
NDUFA3
NDUFA4
NDUFA5
NDUFA6
NDUFA7
NDUFA8
NDUFA9
NDUFAB1
NDUFAF1
NDUFAF2
NDUFAF3
NDUFAF4

NDUFAF5
NDUFAF6
NDUFAF7
NDUFB1
NDUFB10
NDUFB11
NDUFB2
NDUFB3
NDUFB4
NDUFB5
NDUFB6
NDUFB7
NDUFB8
NDUFB9
NDUFC1
NDUFC2
NDUFS1
NDUFS2
NDUFS3
NDUFS4
NDUFS5
NDUFS6
NDUFS7
NDUFS8
NDUFV1
NDUFV2
NDUFV3
NEB
NEBL
NECAP1
NECTIN1
NECTIN4
NEDD4L
NEFH
NEFL
NEK1
NEK2
NEK8
NEK9
NEU1
NEUROD1
NEUROG3
NEXMIF
NEXN
NF1
NFAT5
NFATC1
NFE2L2

NFIA
NFIX
NFKB1
NFKB2
NFKBIA
NFKBIL1
NFS1
NFU1
NGF
NGLY1
NHEJ1
NHLRC1
NHP2
NHS
NID1
NIN
NIPA1
NIPAL4
NIPBL
NKX2-1
NKX2-5
NKX2-6
NKX3-2
NLGN3
NLGN4X
NLRC4
NLRP1
NLRP12
NLRP3
NLRP7
NME1
NME8
NMNAT1
NNT
NOBOX
NOD2
NODAL
NOG
NOL3
NONO
NOP10
NOP56
NOS2
NOS3
NOTCH1
NOTCH2
NPAT
NPC1

NPC1L1
NPC2
NPHP1
NPHP3
NPHP4
NPHS1
NPHS2
NPM1
NPPA
NPR2
NPR2L
NPRL2
NPRL3
NPSR1
NQO1
NR0B1
NR0B2
NR1D2
NR1H4
NR2E3
NR2F1
NR2F2
NR3C1
NR3C2
NR4A2
NR5A1
NRAS
NRG1
NRIP1
NRL
NRP1
NRXN1
NSD1
NSD2
NSD3
NSDHL
NSMCE2
NSMCE3
NSMF
NSUN2
NSUN3
NT5C2
NT5C3A
NT5E
NTF4
NTRK1
NTRK2
NUB1

NUBPL
NUDT15
NUMA1
NUP107
NUP155
NUP188
NUP205
NUP214
NUP62
NUP93
NUS1
NXF5
NYX
OAS1
OAT
OBSL1
OCA2
OCLN
OCRL
ODAM
ODAPH
ODC1
OFD1
OGDH
OGG1
OGT
OLR1
OPA1
OPA3
OPCML
OPHN1
OPLAH
OPN1LW
OPN1MW
OPN1SW
OPTN
OR2J3
ORAI1
ORC1
ORC4
ORC6
OSBPL2
OSGEP
OSMR
OSTM1
OTC
OTOA
OTOF

OTOG
OTOGL
OTUD4
OTUD6B
OTULIN
OTX2
OVOL2
OXA1L
OXCT1
P2RX1
P2RX2
P2RY12
P3H1
P3H2
P4HA2
P4HB
P4HTM
PABPN1
PACS1
PACS2
PADI3
PADI4
PADI6
PAFAH1B1
PAH
PAK3
PALLD
PAM16
PANK2
PANX1
PAPSS2
PARK7
PARN
PARS2
PAX1
PAX2
PAX3
PAX4
PAX6
PAX7
PAX8
PAX9
PAXX
PBRM1
PBX1
PC
PCARE
PCBD1

PCCA
PCCB
PCDH15
PCDH19
PCGF2
PCLO
PCNA
PCNT
PCSK1
PCSK9
PCYT1A
PDCD1
PDCD10
PDCN
PDE10A
PDE11A
PDE3A
PDE4D
PDE6A
PDE6B
PDE6C
PDE6D
PDE6G
PDE6H
PDE8B
PDGFB
PDGFRB
PDGFRL
PDHA1
PDHB
PDHX
PDIA2
PDK1
PDK2
PDK3
PDK4
PDLIM3
PDP1
PDSS1
PDSS2
PDX1
PDYN
PDZD7
PEPD
PER2
PER3
PERP
PET100

PET117
PEX1
PEX10
PEX11B
PEX12
PEX13
PEX14
PEX16
PEX19
PEX2
PEX26
PEX3
PEX5
PEX6
PEX7
PFKM
PFN1
PGAM2
PGAP1
PGAP2
PGAP3
PGK1
PGM1
PGM3
PHB
PHC1
PHEX
PHF11
PHF21A
PHF6
PHF8
PHGDH
PHIP
PHKA1
PHKA2
PHKB
PHKG2
PHOX2A
PHOX2B
PHYH
PHYKPL
PI4KA
PIBF1
PICALM
PIEZO1
PIEZO2
PIFO
PIGA

PIGC
PIGG
PIGL
PIGM
PIGN
PIGO
PIGT
PIGV
PIGW
PIGY
PIH1D3
PIK3CA
PIK3CD
PIK3R1
PIK3R2
PIK3R5
PIKFYVE
PINK1
PIP5K1C
PITPNM3
PITRM1
PITX1
PITX2
PITX3
PJVK
PKD1
PKD1L1
PKD2
PKHD1
PKLR
PKP1
PKP2
PLA2G2A
PLA2G4A
PLA2G5
PLA2G6
PLA2G7
PLAA
PLAG1
PLAGL1
PLAU
PLCB1
PLCB4
PLCD1
PLCE1
PLCG2
PLCZ1
PLD1

PLD3
PLEC
PLEKHG2
PLEKHG5
PLEKHM1
PLEKHM2
PLG
PLIN1
PLK4
PLN
PLOD1
PLOD2
PLOD3
PLP1
PLPBP
PLS3
PLTP
PLVAP
PLXND1
PML
PMM2
PMP22
PMPCA
PMPCB
PMVK
PNKD
PNKP
PNLIP
PNP
PNPLA1
PNPLA2
PNPLA6
PNPLA8
PNPO
PNPT1
POC1A
POC1B
PODXL
POF1B
POFUT1
POGLUT1
POGZ
POLA1
POLE
POLE2
POLG
POLG2
POLH

POLR1A
POLR1C
POLR1D
POLR3A
POLR3B
POMC
POMGNT1
POMGNT2
POMK
POMP
POMT1
POMT2
PON1
PON2
POR
PORCN
POU1F1
POU3F3
POU3F4
POU4F3
POU6F2
PPA2
PPARA
PPARG
PPARGC1B
PPCS
PPIB
PPL
PPM1D
PPM1K
PPOX
PPP1CB
PPP1R15B
PPP1R3A
PPP2CA
PPP2R1A
PPP2R1B
PPP2R2B
PPP2R5B
PPP2R5C
PPP2R5D
PPP3CA
PPT1
PQBP1
PRCC
PRCD
PRDM1
PRDM12

PRDM16
PRDM5
PRDM6
PRDM8
PREPL
PRF1
PRG4
PRICKLE1
PRICKLE2
PRIMPOL
PRKAA1
PRKACA
PRKACG
PRKAG2
PRKAG3
PRKCA
PRKCD
PRKCG
PRKCH
PRKCSH
PRKD1
PRKDC
PRKG1
PRKN
PRKRA
PRLR
PRMT7
PRNP
PROC
PRODH
PROK2
PROKR2
PROM1
PROP1
PROS1
PROX1
PROZ
PRPF3
PRPF31
PRPF4
PRPF6
PRPF8
PRPH
PRPH2
PRPS1
PRR12
PRRT2
PRRX1

PRSS1
PRSS12
PRSS56
PRUNE1
PRX
PSAP
PSAT1
PSENEN
PSMA3
PSMA6
PSMB4
PSMB8
PSMB9
PSMC3IP
PSMD12
PSMG2
PSPH
PSTPIP1
PTCH1
PTCH2
PTCHD1
PTDSS1
PTEN
PTF1A
PTGDR
PTGER2
PTGIS
PTH
PTH1R
PTHB1
PTHLH
PTPN1
PTPN11
PTPN12
PTPN14
PTPN22
PTPRC
PTPRF
PTPRJ
PTPRO
PTPRQ
PTRH2
PTRHD1
PTS
PUF60
PUM1
PURA
PUS1

PUS3
PUS7
PXDN
PYCR1
PYCR2
PYGL
PYGM
PYROXD1
QARS
QDPR
QRICH1
QRSL1
RAB10
RAB11B
RAB18
RAB23
RAB27A
RAB28
RAB33B
RAB39B
RAB3GAP1
RAB3GAP2
RAB40AL
RAB7A
RAC1
RAC2
RAD21
RAD50
RAD51
RAD51B
RAD54B
RAD54L
RAF1
RAG1
RAG2
RAI1
RALA
RANBP2
RANGRF
RAP1GDS1
RAPSN
RARB
RARS
RARS2
RASA1
RASGRP1
RASGRP2
RAX

RAX2
RB1
RB1CC1
RBBP8
RBCK1
RBFOX1
RBFOX2
RBM10
RBM20
RBM28
RBM8A
RBMX
RBP3
RBP4
RBPJ
RCBTB1
RD3
RDH11
RDH12
RDH5
RDX
RECQL2
RECQL3
RECQL4
REEP1
REEP2
REEP6
RELB
RELN
REN
RERE
REST
RET
RETN
RETREG1
REV3L
RFT1
RFX5
RFX6
RFXANK
RFXAP
RGR
RGS9
RGS9BP
RHAG
RHBDF2
RHCE
RHD

RHEB
RHO
RHOBTB2
RHOH
RIL
RIMS1
RIN2
RIPK1
RIPK4
RIPOR2
RIPPLY2
RIT1
RLBP1
RLIM
RMND1
RMRP
RNASEH1
RNASEH2A
RNASEH2B
RNASEH2C
RNASEL
RNASET2
RNF113A
RNF125
RNF135
RNF139
RNF168
RNF170
RNF212
RNF213
RNF216
RNF31
RNF6
RNU4ATAC
ROBO1
ROBO2
ROBO3
ROBO4
ROCK2
ROGDI
ROM1
ROR2
RORA
RORC
RP1
RP1L1
RP2
RP9

RPE65
RPGR
RPGRIP1
RPGRIP1L
RPIA
RPL10
RPL11
RPL15
RPL21
RPL26
RPL35A
RPL5
RPS10
RPS14
RPS15
RPS17
RPS19
RPS24
RPS26
RPS28
RPS29
RPS6KA3
RPS7
RPSA
RRAS2
RREB1
RRM2B
RS1
RSPH1
RSPH3
RSPH4A
RSPH9
RSPO1
RSPO4
RSPRY1
RSRC1
RTEL1
RTN2
RTN4IP1
RTN4R
RTTN
RUBCN
RUNX2
RUSC2
RXFP2
RXYLT1
RYR1
RYR2

S1PR2
SACS
SAG
SALL1
SALL2
SALL4
SAMD9
SAMD9L
SAMHD1
SANS
SAR1B
SARDH
SARS
SARS2
SART3
SASH1
SASS6
SATB2
SBDS
SBF1
SBF2
SC5D
SCAPER
SCARB1
SCARB2
SCARF2
SCGB3A2
SCN10A
SCN11A
SCN1A
SCN1B
SCN2A
SCN2B
SCN3A
SCN3B
SCN4A
SCN4B
SCN5A
SCN8A
SCN9A
SCNN1A
SCNN1B
SCNN1G
SCO1
SCO2
SCP2
SCYL1
SDC3

SDCCAG8
SDHAF1
SDHB
SDR9C7
SEC23A
SEC23B
SEC24C
SEC24D
SEC61A1
SEC63
SECISBP2
SELENON
SELP
SEMA3A
SEMA3D
SEMA3E
SEMA4A
SEMA7A
SEPSECS
Sep/12
Sep/09
SERAC1
SERPINA1
SERPINA3
SERPINA6
SERPINA7
SERPINB6
SERPINB7
SERPINB8
SERPINC1
SERPIND1
SERPINE1
SERPINF1
SERPINF2
SERPING1
SERPINH1
SERPINI1
SESN1
SET
SETBP1
SETD1A
SETD1B
SETD2
SETD5
SETX
SF1
SF3B1
SF3B4

SFRP4
SFTPA1
SFTPA2
SFTPB
SFTPC
SFXN4
SGCA
SGCB
SGCD
SGCE
SGCG
SGO1
SGPL1
SGSH
SH2B1
SH2B3
SH2D1A
SH3BP2
SH3PXD2B
SH3TC2
SHANK2
SHANK3
SHH
SHOC2
SHOX
SHOXY
SHPK
SHROOM3
SHROOM4
SI
SIAE
SIGMAR1
SIK1
SIL1
SIM1
SIN3A
SIPA1L3
SIX1
SIX2
SIX3
SIX5
SIX6
SKI
SKIV2L
SLC10A2
SLC10A7
SLC11A1
SLC11A2

SLC12A1
SLC12A3
SLC12A5
SLC12A6
SLC13A5
SLC14A1
SLC16A1
SLC16A12
SLC16A2
SLC17A3
SLC17A5
SLC17A8
SLC17A9
SLC18A3
SLC19A2
SLC19A3
SLC1A1
SLC1A2
SLC1A3
SLC1A4
SLC20A1
SLC20A2
SLC22A12
SLC22A18
SLC22A4
SLC22A5
SLC24A1
SLC24A4
SLC24A5
SLC25A1
SLC25A10
SLC25A12
SLC25A13
SLC25A15
SLC25A19
SLC25A20
SLC25A21
SLC25A22
SLC25A24
SLC25A26
SLC25A3
SLC25A32
SLC25A38
SLC25A4
SLC25A42
SLC25A46
SLC26A1
SLC26A2

SLC26A3
SLC26A4
SLC26A5
SLC26A8
SLC27A4
SLC27A5
SLC29A3
SLC2A1
SLC2A10
SLC2A2
SLC2A9
SLC30A10
SLC30A2
SLC30A8
SLC33A1
SLC34A1
SLC34A2
SLC34A3
SLC35A1
SLC35A2
SLC35A3
SLC35C1
SLC35D1
SLC36A2
SLC37A4
SLC38A8
SLC39A12
SLC39A13
SLC39A14
SLC39A4
SLC39A5
SLC39A8
SLC3A1
SLC40A1
SLC41A1
SLC45A2
SLC46A1
SLC4A1
SLC4A10
SLC4A11
SLC4A4
SLC52A1
SLC52A2
SLC52A3
SLC5A1
SLC5A2
SLC5A5
SLC5A7

SLC6A1
SLC6A14
SLC6A17
SLC6A19
SLC6A2
SLC6A20
SLC6A3
SLC6A4
SLC6A5
SLC6A8
SLC6A9
SLC7A14
SLC7A7
SLC7A9
SLC9A1
SLC9A3
SLC9A3R1
SLC9A6
SLC9A7
SLC9A9
SLCO1B1
SLCO1B3
SLCO2A1
SLFN14
SLITRK1
SLITRK6
SLMAP
SLURP1
SLURP2
SLX4
SMAC
SMAD1
SMAD2
SMAD3
SMAD4
SMAD6
SMAD7
SMAD9
SMARCA1
SMARCA2
SMARCA4
SMARCAD1
SMARCAL1
SMARCB1
SMARCC2
SMARCD2
SMARCE1
SMC1A

SMC3
SMCHD1
SMG9
SMIM1
SMN1
SMN2
SMO
SMOC1
SMOC2
SMPD1
SMPX
SMS
SNAI2
SNAP25
SNAP29
SNCA
SNCAIP
SNCB
SNIP1
SNORA2C
SNORD118
SNRNP200
SNRPB
SNRPE
SNRPN
SNTA1
SNX10
SNX14
SOBP
SOCS4
SOD1
SOD2
SOD3
SOHLH1
SON
SORL1
SORT1
SOS1
SOS2
SOST
SOX10
SOX11
SOX17
SOX18
SOX2
SOX3
SOX4
SOX5

SOX9
SP110
SP7
SPAG1
SPARC
SPART
SPAST
SPATA16
SPATA5
SPATA7
SPECC1L
SPEG
SPG11
SPG21
SPG7
SPINK1
SPINK5
SPINT2
SPOCK1
SPPL2A
SPR
SPRED1
SPRTN
SPRY2
SPRY4
SPTA1
SPTAN1
SPTB
SPTBN2
SPTLC1
SPTLC2
SQSTM1
SRC
SRCAP
SRD5A2
SRD5A3
SRGAP1
SRP72
SRPX2
SRY
SSR4
SSTR5
SSX1
SSX2
ST14
ST3GAL3
ST3GAL5
STAC3

STAG1
STAG3
STAMBP
STAP1
STAR
STAT1
STAT2
STAT3
STAT4
STAT5B
STAT6
STEAP3
STIL
STIM1
STK4
STN1
STOX1
STRA6
STRADA
STRC
STS
STT3A
STT3B
STUB1
STX11
STX16
STX1B
STX3
STXBP1
STXBP2
SUCLA2
SUCLG1
SUCLG2
SUFU
SUGCT
SULF1
SULT2B1
SUMF1
SUMO1
SUMO4
SUN5
SUOX
SURF1
SUZ12
SVBP
SYCE1
SYCP3
SYN1

SYN2
SYNCRIP
SYNE1
SYNE2
SYNE4
SYNGAP1
SYNJ1
SYP
SYT1
SYT14
SYT2
SZT2
TAB2
TAC3
TACO1
TACR3
TACSTD2
TAF1
TAF13
TAF2
TAF4B
TAF6
TAL1
TAL2
TALDO1
TANC2
TANGO2
TAP1
TAP2
TAPBP
TAPT1
TARDBP
TARS2
TAS2R16
TAS2R38
TAT
TAZ
TBC1D20
TBC1D23
TBC1D24
TBC1D4
TBC1D7
TBCD
TBCE
TBCK
TBK1
TBL1XR1
TBL1Y

TBP
TBR1
TBX1
TBX15
TBX18
TBX19
TBX2
TBX20
TBX21
TBX22
TBX3
TBX4
TBX5
TBX6
TBXA2R
TBXAS1
TBXT
TCAP
TCF12
TCF20
TCF3
TCF4
TCF7L2
TCHH
TCIRG1
TCN2
TCOF1
TCTN1
TCTN2
TCTN3
TDGF1
TDP1
TDP2
TDRD7
TEAD1
TECPR2
TECR
TECRL
TECT1
TECTA
TEK
TELO2
TENM1
TENM3
TENM4
TERC
TERF2IP
TET2

TEX11
TF
TFAM
TFAP2A
TFAP2B
TFE3
TFG
TFR2
TFRC
TG
TGDS
TGFB1
TGFB2
TGFB3
TGFBI
TGFBR1
TGFBR2
TGFBR3
TGIF1
TGM1
TGM3
TGM5
TGM6
TH
THAP1
THBD
THBS1
THBS2
THG1L
THOC2
THOC6
THPO
THRA
THRB
TIA1
TICAM1
TIMM44
TIMM50
TIMM8A
TIMMDC1
TIMP3
TINF2
TIRAP
TJP2
TK2
TKT
TLE6
TLK2

TLL1
TLR1
TLR2
TLR3
TLR4
TLR5
TM4SF20
TMC1
TMC6
TMC8
TMCO1
TMEM107
TMEM126A
TMEM126B
TMEM138
TMEM165
TMEM17
TMEM173
TMEM186
TMEM199
TMEM216
TMEM231
TMEM237
TMEM240
TMEM38B
TMEM43
TMEM65
TMEM67
TMEM70
TMEM98
TMIE
TMLHE
TMPO
TMPRSS15
TMPRSS3
TMPRSS6
TMTC3
TNC
TNF
TNFAIP3
TNFRSF10B
TNFRSF11A
TNFRSF11B
TNFRSF13B
TNFRSF13C
TNFRSF1A
TNFRSF4
TNFSF11

TNFSF12
TNFSF4
TNIK
TNNC1
TNNI2
TNNI3
TNNI3K
TNNT1
TNNT2
TNNT3
TNPO3
TNXB
TOE1
TOP1
TOP2A
TOPORS
TOR1A
TOR1AIP1
TOR1AIP2
TP53RK
TP63
TPCN2
TPH2
TPI1
TPK1
TPM1
TPM2
TPM3
TPMT
TPO
TPP1
TPP2
TPRKB
TPRN
TRAF3
TRAF3IP1
TRAF3IP2
TRAF6
TRAF7
TRAIP
TRAP1
TRAPPC11
TRAPPC2
TRAPPC6B
TRAPPC9
TRDN
TREM2
TREX1

TRH
TRHR
TRIM2
TRIM32
TRIM37
TRIM44
TRIO
TRIOBP
TRIP11
TRIP12
TRIP4
TRIT1
TRMT1
TRMT10A
TRMT10C
TRMT5
TRMU
TRNH
TRNI
TRNK
TRNP1
TRNQ
TRNS2
TRNT
TRNT1
TRPA1
TRPC3
TRPC4
TRPC6
TRPM1
TRPM2
TRPM4
TRPM6
TRPM7
TRPS1
TRPV3
TRPV4
TRRAP
TSC1
TSC2
TSEN15
TSEN2
TSEN34
TSEN54
TSFM
TSG101
TSHB
TSHR

TSHZ1
TSPAN12
TSPAN7
TSPEAR
TSPYL1
TSR2
TTBK2
TTC19
TTC21B
TTC25
TTC37
TTC7A
TTC8
TTI2
TTLL5
TTN
TTPA
TTR
TUB
TUBA1A
TUBA4A
TUBA8
TUBB
TUBB1
TUBB2A
TUBB2B
TUBB3
TUBB4A
TUBB8
TUBG1
TUBGCP4
TUBGCP6
TUFM
TUFT1
TULP1
TUSC3
TWIST1
TWIST2
TWNK
TXN2
TXNL4A
TXNRD2
TYK2
TYMP
TYR
TYROBP
TYRP1
UBA1

UBA5
UBE2A
UBE2T
UBE3A
UBE3B
UBIAD1
UBQLN2
UBR1
UBTF
UCHL1
UCP1
UCP2
UCP3
UFC1
UFD1
UFM1
UFSP2
UGT1A1
UGT1A4
UGT1A5
UGT2B17
UMOD
UMPS
UNC119
UNC13A
UNC13D
UNC45B
UNC80
UNC93B1
UNG
UPB1
UPF3B
UPK3A
UQCC1
UQCC2
UQCC3
UQCR10
UQCR11
UQCRB
UQCRC1
UQCRC2
UQCRFS1
UQCRH
UQCRQ
UROC1
UROD
UROS
USB1

USF1
USH1C
USH1G
USH2A
USP18
USP27X
USP7
USP9X
USP9Y
UTP4
UVRAG
UVSSA
VAC14
VAMP1
VAMP2
VANGL1
VANGL2
VAPB
VARS2
VAV1
VAX1
VCAN
VCL
VCP
VDR
VEGFA
VEGFC
VHL
VIL1
VIM
VIPAS39
VKORC1
VLDLR
VMA21
VNN1
VPS11
VPS13A
VPS13B
VPS13C
VPS13D
VPS33A
VPS33B
VPS35
VPS37A
VPS39
VPS45
VPS53
VRK1

VSX1
VSX2
VWA3B
VWF
WAC
WARS2
WAS
WASF1
WASHC4
WASHC5
WDPCP
WDR1
WDR11
WDR13
WDR19
WDR26
WDR34
WDR35
WDR36
WDR4
WDR45
WDR45B
WDR60
WDR62
WDR72
WDR73
WDR81
WFS1
WHRN
WIPF1
WNK1
WNK4
WNT1
WNT10A
WNT10B
WNT2B
WNT3
WNT4
WNT5A
WNT7A
WRAP53
WRN
WT1
WWC1
WWOX
XBP1
XDH
XEDAR

XG
XIAP
XIST
XK
XPA
XPC
XPNPEP2
XPNPEP3
XPR1
XRCC2
XRCC3
XRCC4
XRCC5
XRCC6
XYLT1
XYLT2
YAP1
YARS
YARS2
YME1L1
YWHAE
YWHAG
YY1
YY1AP1
ZAK
ZAP70
ZBTB11
ZBTB16
ZBTB18
ZBTB20
ZBTB24
ZBTB42
ZC3H14
ZC4H2
ZDHHC15
ZDHHC9
ZEB1
ZEB2
ZFAT
ZFHX3
ZFP57
ZFPM2
ZFYVE26
ZFYVE27
ZIC1
ZIC2
ZIC3
ZIC4

ZIM2
ZMIZ1
ZMPSTE24
ZMYM3
ZMYND10
ZMYND11
ZMYND15
ZNF141
ZNF148
ZNF252P
ZNF292
ZNF335
ZNF365
ZNF407
ZNF408
ZNF41
ZNF423
ZNF462
ZNF469
ZNF480
ZNF513
ZNF565
ZNF592
ZNF644
ZNF674
ZNF687
ZNF711
ZNF750
ZNF81
ZP1
ZSWIM6
