## supplementary table 1 for "MOLGENIS VIP: an open-source and modular pipeline for high-throughput and integrated DNA variant analysis"

| Annotation source | Version | Description | Region | Reference |
|---|---|---|---|---|
| PolyPhen2 | v2.2.3 | Machine learning (ML)-based method to determine pathogenic effect on protein function and structure. Compares features between mutant and wild-type (WT) alleles that define amino acid replacement. Calculates score from 0 to 1. | Coding | 1 |
| SIFT | v6.2.1 | Exploits sequence homology and amino acid changes at well-conserved positions within the protein family to determine the effect on protein function. Calculates score from 0 to 1. | Coding | 2 |
| AnnotSV | v3.3.6 | Combines functionally, regulatory, and clinically relevant information and a panel of different datasets to interpret SV pathogenicity. Pathogenicity is devided in 5 grades (1-5). | Coding, non-coding | 3 |
| SpliceAI | v1.3.1 | A national platform for sharing validated variant classifications performed by multiple Dutch genome diagnostic laboratories. Variants are annotated with the consensus VKGL classification (benign (B), likely benign (LB), variant of unknown significance (VUS), likely pathogenic (LP), pathogenic (P)). | Coding, non-coding | 4 |
| CAPICE | v5.1.2 | ML-based method to predict pathogenicity of SNVs and indels using diverse genomic features (e.g. genetic context, gene model annotations, evolutionary constraints). Calculates a score from 0 to 1. | Coding | 5 |
| UTRannotator | v1.0 | By evaluating if variants in five prime untranslated regions (5'UTRs) are not a multiple of three, it determines if variants interfere with existing or introduce new start- or stopcodons, creating or disrupting upstream open reading frames (uORFs). Outcomes can be 'true' or 'false'. | Non-coding | 6 |
| VKGL database | release 2023-11-01 | A national platform for sharing validated variant classifications performed by multiple Dutch genome diagnostic laboratories. Variants are annotated with the consensus VKGL classification (benign (B), likely benign (LB), variant of unknown significance (VUS), likely pathogenic (LP), pathogenic (P)). | Coding, non-coding | 7 |
| ClinVar database | release 2024-01-19 | Public archive of clinical evidence of relationships among sequence variation and human phenotypes and variant classifications. Variants are annotated with ClinVar classification (B, LB, VUS, LP or P). | Coding, non-coding | 8 |
| GnomAD database | v4.0 | Genome aggregation database developed by the Exome Aggegation Consortium summarizing population variant data, such as constraint scores, variant co-occurrence and population allele frequencies. | Coding, non-coding | 9 |
| Stranger | v0.8.1 | ExpansionHunter annotates the number of times a repeat is present in a bam file. Stranger annotates output files from ExpansionHunter with the pathogenicity of the repeat sizes. Possible values are 'normal', 'pre mutation' and 'full mutation'. | Coding, non-coding | 10 |
| Grantham | - | Score to predict evolutionary distance between two amino acids. A a lower score represents a lower evolutionary distance and vice versa. | Coding | 11 |
| GADO | v1.0.3 | Gene-expression-based model to predict phenotypes caused by genes. It can be used to predict previously unknown disease gene associations and genes that previously have been associated with disease. | Coding | 12 |
| AlphScore | release 2023-08-25 | Artificial-intellegence-based system to predict three-dimensional structures of proteins from amino acid sequences. The calculated score represents the predicted impact of missense variants on pathogenicity. | Coding | 13 |
| ncER | v2.0 | Non-coding essential regulation score (ncER) that indicates if a region is likely to have a regulatory function. Calculated scores range from 0 to 1. | Non-coding | 14 |

Supplementary table 1 **Annotation sources used in annotation module of VIP.** The first two columns represent the name and version of the annotation source that is part of VIP version 7.4.0. The third column provides a description of the annotation source. The fourth column indicates whether the annotation source applies to coding and/or non-coding regions of the genome. The last column contains a reference to the specific annotation source.
