## supplementary table 2 for "MOLGENIS VIP: an open-source and modular pipeline for high-throughput and integrated DNA variant analysis"

| virtual gene panel | proband sample_id | variant | transcript | gene | chrom | position | ref | alt |
|---|---|---|---|---|---|---|---|---|
| Postnatal clinical exome | 12I_5Post_NX284_v_Index | LP_MED12_c.1101+1G>A | NM_005120.2 | MED12 | X | 70341667 | G | A |
| Developmental delay | 9I_OA_NX307_m_Index | P_ADNP c.2155delT | NM_015339.4 | ADNP | 20 | 49509096 | A | . |
| Postnatal clinical exome | 1I_5Post_NX284_v_Index | LP_Notch 1 c.6897_6903delCGGGTCC | NM_017617.5 | NOTCH1 | 9 | 139391288 | GGAC…G | . |
| Prenatal clinical exome | 19I_5Pre_NX285_m_Index | P_KCNQ1 c.887T>C | NM_000218.2 | KCNQ1 | 11 | 2594182 | T | C |
| Prenatal clinical exome | 21I_5Pre_NX283_m_Index | P_FGFR3 c.1111A>T | NM_000142.4 | FGFR3 | 4 | 1806092 | A | T |
| Developmental delay | 14I_OA_NX307_v_Index | P_GLI3 c.4431dupT | NM_000168.5 | GLI3 | 7 | 42004239 | A | . |
| Prenatal clinical exome | 18I_5Pre_NX283_m_Index | LP_BICD2 c.2048T>C | NM_001003800.1 | BICD2 | 9 | 95480879 | A | G |
| Postnatal clinical exome | 18I_5Post_NX284_m_Index | P_NROB1_c.728_734delTGCGGCC | NM_000475.4 | NROB1 | X | 30326747 | GGCC…A | . |
| Postnatal clinical exome | 15I_5Post_NX284_m_Index | P_RRAS2 c.208G>A | NM_012250.6 | RRAS2 | 11 | 14316397 | C | T |
| Postnatal clinical exome | 21I_5Post_NX284_v_Index | P_MAGEL2 c.1996dupC | NM_019066.4 | MAGEL2 | 15 | 23890893 | . | G |
| Postnatal clinical exome | 24I_5Post_NX284_v_Index | P_PACS1 c.607C>T | NM_018026.3 | PACS1 | 11 | 65978677 | C | T |
| Prenatal clinical exome | 2I_5Pre_NX283_v_Index | P_RIT1 c.170C>G | NM_006912.6 | RIT1 | 1 | 155874589 | G | C |
| Dilated cardiomyopathy children | 1I_kDCM_NX288_m_Index | P_MYL2 c.403-1G>C | NM_000432.3 | MYL2 | 12 | 111348980 | C | G |
| Prenatal clinical exome | 17I_5Pre_NX285_m_Index | P_IRF6 c.202C>T | NM_006147.3 | IRF6 | 1 | 209969870 | G | A |
| Postnatal clinical exome | 5I_5Post_NX284_m_Index | P_NPC1 c.3451G>A | NM_000271.4 | NPC1 | 18 | 21115459 | C | T |
| Dilated cardiomyopathy children | 4I_kDCM_NX288_m_Index | P_MYBPC3 c.927-2A>G | NM_000256.3 | MYBPC3 | 11 | 47367923 | T | C |
| Postnatal clinical exome | 5I_5Post_NX284_m_Index | P_NPC1 c.1918G>A | NM_000271.4 | NPC1 | 18 | 21124953 | C | T |
| Postnatal clinical exome | 22I_5Post_NX284_m_Index | P_MECP2 c.806delG | NM_004992.3 | MECP2 | X | 153296473 | C | . |
| Dilated cardiomyopathy children | 4I_kDCM_NX288_m_Index | P_MYBPC3 c.2827C>T | NM_000256.3 | MYBPC3 | 11 | 47356671 | G | A |

Supplementary table 2 **Routine diagnostic cohort** Probands within routine diagnostic cohort.
