## supplementary table 3 for "MOLGENIS VIP: an open-source and modular pipeline for high-throughput and integrated DNA variant analysis"

| proband sample | chrom | start_pos | end_pos | ref | alt | gene | status | significance |
|---|---|---|---|---|---|---|---|---|
| E481613 | 2 | 179598569 | 179598569 | GTCC | G | TTN | Causative | Pathogenic |
| E603477 | MT | 3243 | 3243 | A | G | MT-TL1 | Causative | Pathogenic |
| E431798 | 15 | 64698591 | 64698591 | C | T | TRIP4 | Causative | Pathogenic |
| E395076 | 17 | 4804151 | 4804151 | A | G | CHRNE | Causative | Pathogenic |
| E989776 | 2 | 241722504 | 241722504 | G | A | KIF1A | Causative | Pathogenic |
| E261621 | 18 | 12337503 | 12337503 | C | T | AFG3L2 | Causative | Pathogenic |
| E100304 | 12 | 79842738 | 79842738 | T | C | SYT1 | Causative | Pathogenic |
| E925075 | 6 | 75861641 | 75861641 | G | C | COL12A1 | Causative | Pathogenic |
| E287287 | 22 | 32210991 | 32210991 | C | T | DEPDC5 | Causative | Pathogenic |
| E470458 | 9 | 109687808 | 109687808 | C | T | ZNF462 | Causative | Pathogenic |
| E313710 | 7 | 4827302 | 4827302 | T | C | AP5Z1 | Causative | Pathogenic |
| E186172 | 7 | 92140317 | 92140317 | CT | C | PEX1 | Causative | Pathogenic |
| E571022 | MT | 9035 | 9035 | T | C | MT-ATP6 | Causative | Pathogenic |
| E830835 | 15 | 42701566 | 42701566 | GA | G | CAPN3 | Causative | Pathogenic |
| E568630 | 1 | 119683231 | 119683231 | A | C | WARS2 | Causative | Pathogenic |
| E724379 | 17 | 48685266 | 48685266 | A | T | CACNA1G | Causative | Pathogenic |
| E597202 | 1 | 45793573 | 45793573 | C | A | HPDL | Causative | Pathogenic |
| E867414 | 20 | 62044909 | 62044909 | G | A | KCNQ2 | Causative | Pathogenic |
| E513765 | 10 | 88813161 | 88813161 | C | A | GLUD1 | Causative | Pathogenic |
| E473734 | 5 | 92929372 | 92929372 | C | T | NR2F1 | Causative | Pathogenic |
| E599679 | 7 | 151880107 | 151880107 | AG | A | KMT2C | Causative | Pathogenic |
| E499319 | 18 | 48593490 | 48593490 | TAGAC | T | SMAD4 | Causative | Pathogenic |
| E822195 | 17 | 60650729 | 60650729 | G | A | TLK2 | Causative | Pathogenic |
| E547381 | 12 | 7048281 | 7048281 | A | C | ATN1 | Causative | Pathogenic |
| E038467 | 7 | 105131982 | 105131982 | C | T | PUS7 | Causative | Pathogenic |
| E440074 | 1 | 27097595 | 27097595 | G | A | ARID1A | Causative | Pathogenic |
| E666478 | 9 | 98268884 | 98268884 | G | C | PTCH1 | Causative | Pathogenic |
| E570269 | 10 | 89272896 | 89272896 | C | A | MINPP1 | Causative | Pathogenic |
| E447588 | 1 | 115280664 | 115280664 | G | A | CSDE1 | Causative | Pathogenic |
| E128884 | 1 | 111146180 | 111146180 | T | G | KCNA2 | Causative | Pathogenic |
| E484318 | 19 | 38931375 | 38931375 | G | A | RYR1 | Causative | Pathogenic |
| E282663 | 1 | 990280 | 990280 | CGTG | T | AGRN | Causative | Pathogenic |
| E476365 | 15 | 45694818 | 45694818 | G | A | SPATA5L1 | Causative | Pathogenic |
| E062108 | 19 | 3656410 | 3656410 | T | C | PIP5K1C | Causative | Pathogenic |
| E221264 | 15 | 25620739 | 25620739 | T | A | UBE3A | Causative | Pathogenic |
| E594094 | 12 | 49579524 | 49579524 | T | C | TUBA1A | Causative | Pathogenic |
| E685615 | 1 | 26764697 | 26764697 | C | A | DHDDS | Causative | Pathogenic |
| E339658 | 8 | 140631149 | 140631149 | C | T | KCNK9 | Causative | Pathogenic |
| E733888 | 4 | 39515774 | 39515774 | G | A | UGDH | Causative | Pathogenic |
| E714923 | 2 | 220285680 | 220285680 | G | A | DES | Causative | Pathogenic |
| E023128 | X | 70346318 | 70346318 | T | A | MED12 | Causative | Pathogenic |

Supplementary table 3 **Solve-RD research cohort** Probands within Solve-RD research cohort.
