## supplementary table 4 for "MOLGENIS VIP: an open-source and modular pipeline for high-throughput and integrated DNA variant analysis"

| project_id | family_id | individual_id | proband | assembly | vcf | sequencing_method |
|---|---|---|---|---|---|---|
| vkgl_202402 | vkgl | vkgl_202402 | TRUE | GRCh37 | /path_to/vkgl_202402.vcf | WES |
| vkgl_202402 | vkgl | vkgl_202402 | TRUE | GRCh37 | /path_to/vkgl_202402_202311_diff.vcf | WES |

Supplementary table 4 **Example of sample sheet containing VKGL variants**
