## supplementary table 5 for "MOLGENIS VIP: an open-source and modular pipeline for high-throughput and integrated DNA variant analysis"

| project_id | family_id | individual_id | paternal_id | maternal_id | sex | affected | proband | hpo_ids | assembly | gvcf | sequencing_method |
|---|---|---|---|---|---|---|---|---|---|---|---|
| FAM0 | FAM0 | 1I_OA_NX307_v_Index_201204 | 1Va_OA_NX307_m_Vader_201204 | 1Moe_OA_NX307_v_Moeder_201204 | female | TRUE | TRUE | HP:0012759, HP:0000729 | GRCh37 | _NX307_v_Index_201204.g.vcf.gz | WES |
| FAM0 | FAM0 | 1Va_OA_NX307_m_Vader_201204 | | | male | FALSE | FALSE | | GRCh37 | OA_NX307_m_Vader_201204.g.vcf.gz | WES |
| FAM0 | FAM0 | 1Moe_OA_NX307_v_Moeder_201204 | | | female | FALSE | FALSE | | GRCh37 | _OA_NX307_v_Moeder_201204.g.vcf.gz | WES |
| FAM1 | FAM1 | 2I_OA_NX307_v_Index_201204 | 2Va_OA_NX307_m_Vader_201204 | 2Moe_OA_NX307_v_Moeder_201204 | female | TRUE | TRUE | HP:0012759, HP:0001250 | GRCh37 | _NX307_v_Index_201204.g.vcf.gz | WES |
| FAM1 | FAM1 | 2Moe_OA_NX307_v_Moeder_201204 | | | female | FALSE | FALSE | | GRCh37 | _OA_NX307_v_Moeder_201204.g.vcf.gz | WES |
| FAM1 | FAM1 | 2Va_OA_NX307_m_Vader_201204 | | | male | FALSE | FALSE | | GRCh37 | OA_NX307_m_Vader_201204.g.vcf.gz | WES |
| FAM10 | FAM10 | 11I_OA_NX307_m_Index_201204 | 11Va_OA_NX307_m_Vader_201204 | 11Moe_OA_NX307_v_Moeder_201204 | male | TRUE | TRUE | [illegible] HP:0001249, HP:0001513, HP:0007834, HP:0000518, HP:0004322, | GRCh37 | /path_to/11I_OA_NX307_m_Index_201204.g.vcf.gz | WES |
| FAM10 | FAM10 | 11Moe_OA_NX307_v_Moeder_201204 | | | female | FALSE | FALSE | | GRCh37 | e_OA_NX307_v_Moeder_201204.g.vcf.gz | WES |

| | | | | | | | | | | | |
|---|---|---|---|---|---|---|---|---|---|---|---|
| FAM10 | FAM10 | 11Va_OA_NX307_m_Vader_201204 | | | male | FALSE | FALSE | | GRCh37 | [illegible]OA_NX307_m_Vader_201204.g.vcf.gz | WES |
| FAM11 | FAM11 | 12I_OA_NX307_m_Index_201204 | 12Va_OA_NX307_m_Vader_201204 | 12Moe_OA_NX307_v_Moeder_201204 | male | TRUE | TRUE | HP:0000750, HP:0000729 | GRCh37 | [illegible]A_NX307_m_Index_201204.g.vcf.gz | WES |
| FAM11 | FAM11 | 12Moe_OA_NX307_v_Moeder_201204 | | | female | FALSE | FALSE | | GRCh37 | [illegible]e_OA_NX307_v_Moeder_201204.g.vcf.gz | WES |
| FAM11 | FAM11 | 12Va_OA_NX307_m_Vader_201204 | | | male | FALSE | FALSE | | GRCh37 | [illegible]OA_NX307_m_Vader_201204.g.vcf.gz | WES |
| FAM12 | FAM12 | 13I_OA_NX307_m_Index_201204 | 13Va_OA_NX307_m_Vader_201204 | 13Moe_OA_NX307_m_Moeder_201204 | male | TRUE | TRUE | [illegible] HP:0012759, HP:0000750, HP:0001290, HP:0011968, HP:0001600 | GRCh37 | /path_to/13I_OA_NX307_m_Index_201204.g.vcf.gz | WES |
| FAM12 | FAM12 | 13Moe_OA_NX307_m_Moeder_201204 | | | female | FALSE | FALSE | | GRCh37 | [illegible]e_OA_NX307_m_Moeder_201204.g.vcf.gz | WES |
| FAM12 | FAM12 | 13Va_OA_NX307_m_Vader_201204 | | | male | FALSE | FALSE | | GRCh37 | [illegible]OA_NX307_m_Vader_201204.g.vcf.gz | WES |
| FAM13 | FAM13 | 14I_OA_NX307_v_Index_201204 | 14Va_OA_NX307_m_Vader_201204 | 14Moe_OA_NX307_v_Moeder_201204 | female | TRUE | TRUE | HP:0010442, HP:0000256, HP:0000316 | GRCh37 | [illegible]A_NX307_v_Index_201204.g.vcf.gz | WES |

| | | | | | | | | | | | |
|---|---|---|---|---|---|---|---|---|---|---|---|
| FAM13 | FAM13 | 14Moe_OA_NX307_v_Moeder_201204 | | | female | FALSE | FALSE | | GRCh37 | e_OA_NX307_v_Moeder_201204.g.vcf.gz | WES |
| FAM13 | FAM13 | 14Va_OA_NX307_m_Vader_201204 | | | male | FALSE | FALSE | | GRCh37 | OA_NX307_m_Vader_201204.g.vcf.gz | WES |
| FAM14 | FAM14 | 15I_OA_NX307_m_Index_201204 | 15Va_OA_NX307_m_Vader_201204 | 15Moe_OA_NX307_v_Moeder_201204 | male | TRUE | TRUE | HP:0000750, HP:0001508, HP:0000253, HP:0011968 | GRCh37 | /path_to/15I_OA_NX307_m_Index_201204.g.vcf.gz | WES |
| FAM14 | FAM14 | 15Moe_OA_NX307_v_Moeder_201204 | | | female | FALSE | FALSE | | GRCh37 | e_OA_NX307_v_Moeder_201204.g.vcf.gz | WES |
| FAM14 | FAM14 | 15Va_OA_NX307_m_Vader_201204 | | | male | FALSE | FALSE | | GRCh37 | OA_NX307_m_Vader_201204.g.vcf.gz | WES |
| FAM15 | FAM15 | 1I_5Pre_NX283_v_Index_201208 | 1Va_5Pre_NX283_m_Vader_201208 | 1Moe_5Pre_NX283_v_Moeder_201208 | female | TRUE | TRUE | HP:0011461, HP:0031853, HP:0003826 | GRCh37 | e_NX283_v_Index_201208.g.vcf.gz | WES |
| FAM15 | FAM15 | 1Moe_5Pre_NX283_v_Moeder_201208 | | | female | FALSE | FALSE | | GRCh37 | _5Pre_NX283_v_Moeder_201208.g.vcf.gz | WES |
| FAM15 | FAM15 | 1Va_5Pre_NX283_m_Vader_201208 | | | male | FALSE | FALSE | | GRCh37 | Pre_NX283_m_Vader_201208.g.vcf.gz | WES |
| FAM16 | FAM16 | 2I_5Pre_NX283_v_Index_201208 | 2Va_5Pre_NX283_m_Vader_201208 | 2Moe_5Pre_NX283_v_Moeder_201208 | female | TRUE | TRUE | HP:0011461, HP:0000474 | GRCh37 | e_NX283_v_Index_201208.g.vcf.gz | WES |

| | | | | | | | | | | | |
|---|---|---|---|---|---|---|---|---|---|---|---|
| FAM16 | FAM16 | 2Moe_5Pre_NX283_v_Moeder_201208 | | | female | FALSE | FALSE | | GRCh37 | _5Pre_NX283_v_Moeder_201208.g.vcf.gz | WES |
| FAM16 | FAM16 | 2Va_5Pre_NX283_m_Vader_201208 | | | male | FALSE | FALSE | | GRCh37 | Pre_NX283_m_Vader_201208.g.vcf.gz | WES |
| FAM17 | FAM17 | 3I_5Pre_NX283_m_Index_201208 | 3Va_5Pre_NX283_m_Vader_201208 | 3Moe_5Pre_NX283_v_Moeder_201208 | male | TRUE | TRUE | HP:0002280, HP:0001320, HP:0006956 | GRCh37 | e_NX283_m_Index_201208.g.vcf.gz | WES |
| FAM17 | FAM17 | 3Moe_5Pre_NX283_v_Moeder_201208 | | | female | FALSE | FALSE | | GRCh37 | _5Pre_NX283_v_Moeder_201208.g.vcf.gz | WES |
| FAM17 | FAM17 | 3Va_5Pre_NX283_m_Vader_201208 | | | male | FALSE | FALSE | | GRCh37 | Pre_NX283_m_Vader_201208.g.vcf.gz | WES |
| FAM18 | FAM18 | 4I_5Pre_NX285_m_Index_201208 | 4Va_5Pre_NX285_m_Vader_201208 | 4Moe_5Pre_NX285_v_Moeder_201208 | male | TRUE | TRUE | HP:0002119, HP:0012443, HP:0001297 | GRCh37 | e_NX285_m_Index_201208.g.vcf.gz | WES |
| FAM18 | FAM18 | 4Moe_5Pre_NX285_v_Moeder_201208 | | | female | FALSE | FALSE | | GRCh37 | _5Pre_NX285_v_Moeder_201208.g.vcf.gz | WES |
| FAM18 | FAM18 | 4Va_5Pre_NX285_m_Vader_201208 | | | male | FALSE | FALSE | | GRCh37 | Pre_NX285_m_Vader_201208.g.vcf.gz | WES |
| FAM19 | FAM19 | 5I_5Pre_NX283_v_Index_201208 | 5Va_5Pre_NX283_m_Vader_201208 | 5Moe_5Pre_NX283_v_Moeder_201208 | female | TRUE | TRUE | HP:0011461, HP:0001539 | GRCh37 | e_NX283_v_Index_201208.g.vcf.gz | WES |

| | | | | | | | | | | | |
|---|---|---|---|---|---|---|---|---|---|---|---|
| FAM19 | FAM19 | 5Moe_5Pre_NX283_v_Moeder_201208 | | | female | FALSE | FALSE | | GRCh37 | _5Pre_NX283_v_Moeder_201208.g.vcf.gz | WES |
| FAM19 | FAM19 | 5Va_5Pre_NX283_m_Vader_201208 | | | male | FALSE | FALSE | | GRCh37 | Pre_NX283_m_Vader_201208.g.vcf.gz | WES |
| FAM2 | FAM2 | 3I_OA_NX307_m_Index_201204 | 3Va_OA_NX307_m_Vader_201204 | 3Moe_OA_NX307_v_Moeder_201204 | male | TRUE | TRUE | HP:0012759, HP:0000729, HP:0001513 | GRCh37 | _NX307_m_Index_201204.g.vcf.gz | WES |
| FAM2 | FAM2 | 3Moe_OA_NX307_v_Moeder_201204 | | | female | FALSE | FALSE | | GRCh37 | _OA_NX307_v_Moeder_201204.g.vcf.gz | WES |
| FAM2 | FAM2 | 3Va_OA_NX307_m_Vader_201204 | | | male | FALSE | FALSE | | GRCh37 | OA_NX307_m_Vader_201204.g.vcf.gz | WES |
| FAM20 | FAM20 | 6I_5Pre_NX285_m_Index_201208 | 6Va_5Pre_NX285_m_Vader_201208 | 6Moe_5Pre_NX285_v_Moeder_201208 | male | TRUE | TRUE | HP:0001561, HP:0000308, HP:0002190, HP:0010963, HP:0002650, | GRCh37 | /path_to/6I_5Pre_NX285_m_Index_201208.g.vcf.gz | WES |
| FAM20 | FAM20 | 6Moe_5Pre_NX285_v_Moeder_201208 | | | female | FALSE | FALSE | | GRCh37 | _5Pre_NX285_v_Moeder_201208.g.vcf.gz | WES |
| FAM20 | FAM20 | 6Va_5Pre_NX285_m_Vader_201208 | | | male | FALSE | FALSE | | GRCh37 | Pre_NX285_m_Vader_201208.g.vcf.gz | WES |

| | | | | | | | | | | | |
|---|---|---|---|---|---|---|---|---|---|---|---|
| FAM21 | FAM21 | 7I_5Pre_NX283_v_Index_201208 | 7Va_5Pre_NX283_m_Vader_201208 | 7Moe_5Pre_NX283_v_Moeder_201208 | female | TRUE | TRUE | HP:0011461 | GRCh37 | e_NX283_v_Index_201208.g.vcf.gz | WES |
| FAM21 | FAM21 | 7Moe_5Pre_NX283_v_Moeder_201208 | | | female | FALSE | FALSE | | GRCh37 | _5Pre_NX283_v_Moeder_201208.g.vcf.gz | WES |
| FAM21 | FAM21 | 7Va_5Pre_NX283_m_Vader_201208 | | | male | FALSE | FALSE | | GRCh37 | Pre_NX283_m_Vader_201208.g.vcf.gz | WES |
| FAM22 | FAM22 | 8I_5Pre_NX283_m_Index_201208 | 8Va_5Pre_NX283_m_Vader_201208 | 8Moe_5Pre_NX283_v_Moeder_201208 | male | TRUE | TRUE | HP:0011461 | GRCh37 | e_NX283_m_Index_201208.g.vcf.gz | WES |
| FAM22 | FAM22 | 8Moe_5Pre_NX283_v_Moeder_201208 | | | female | FALSE | FALSE | | GRCh37 | _5Pre_NX283_v_Moeder_201208.g.vcf.gz | WES |
| FAM22 | FAM22 | 8Va_5Pre_NX283_m_Vader_201208 | | | male | FALSE | FALSE | | GRCh37 | Pre_NX283_m_Vader_201208.g.vcf.gz | WES |
| FAM23 | FAM23 | 9I_5Pre_NX283_m_Index_201208 | 9Va_5Pre_NX283_m_Vader_201208 | 9Moe_5Pre_NX283_v_Moeder_201208 | male | TRUE | TRUE | HP:0011461, HP:0002250 | GRCh37 | e_NX283_m_Index_201208.g.vcf.gz | WES |
| FAM23 | FAM23 | 9Moe_5Pre_NX283_v_Moeder_201208 | | | female | FALSE | FALSE | | GRCh37 | _5Pre_NX283_v_Moeder_201208.g.vcf.gz | WES |
| FAM23 | FAM23 | 9Va_5Pre_NX283_m_Vader_201208 | | | male | FALSE | FALSE | | GRCh37 | Pre_NX283_m_Vader_201208.g.vcf.gz | WES |

| | | | | | | | | | | | |
|---|---|---|---|---|---|---|---|---|---|---|---|
| FAM24 | FAM24 | 10I_5Pre_NX283_m_Index_201208 | 10Va_5Pre_NX283_m_Vader_201208 | 10Moe_5Pre_NX283_v_Moeder_201208 | male | TRUE | TRUE | HP:0011461 | GRCh37 | Pre_NX283_m_Index_201208.g.vcf.gz | WES |
| FAM24 | FAM24 | 10Moe_5Pre_NX283_v_Moeder_201208 | | | female | FALSE | FALSE | | GRCh37 | e_5Pre_NX283_v_Moeder_201208.g.vcf.gz | WES |
| FAM24 | FAM24 | 10Va_5Pre_NX283_m_Vader_201208 | | | male | FALSE | FALSE | | GRCh37 | 5Pre_NX283_m_Vader_201208.g.vcf.gz | WES |
| FAM25 | FAM25 | 11I_5Pre_NX285_v_Index_201208 | 11Va_5Pre_NX285_m_Vader_201208 | 11Moe_5Pre_NX285_v_Moeder_201208 | female | TRUE | TRUE | HP:0011461 | GRCh37 | Pre_NX285_v_Index_201208.g.vcf.gz | WES |
| FAM25 | FAM25 | 11Moe_5Pre_NX285_v_Moeder_201208 | | | female | FALSE | FALSE | | GRCh37 | e_5Pre_NX285_v_Moeder_201208.g.vcf.gz | WES |
| FAM25 | FAM25 | 11Va_5Pre_NX285_m_Vader_201208 | | | male | FALSE | FALSE | | GRCh37 | 5Pre_NX285_m_Vader_201208.g.vcf.gz | WES |
| FAM26 | FAM26 | 12I_5Pre_NX285_m_Index_201208 | 12Va_5Pre_NX285_m_Vader_201208 | 12Moe_5Pre_NX285_v_Moeder_201208 | male | TRUE | TRUE | HP:0011461 | GRCh37 | Pre_NX285_m_Index_201208.g.vcf.gz | WES |
| FAM26 | FAM26 | 12Moe_5Pre_NX285_v_Moeder_201208 | | | female | FALSE | FALSE | | GRCh37 | e_5Pre_NX285_v_Moeder_201208.g.vcf.gz | WES |
| FAM26 | FAM26 | 12Va_5Pre_NX285_m_Vader_201208 | | | male | FALSE | FALSE | | GRCh37 | 5Pre_NX285_m_Vader_201208.g.vcf.gz | WES |

| | | | | | | | | | | | |
|---|---|---|---|---|---|---|---|---|---|---|---|
| FAM27 | FAM27 | 13I_5Pre_NX283_m_Index_201208 | 13Va_5Pre_NX283_m_Vader_201208 | 13Moe_5Pre_NX283_v_Moeder_201208 | male | TRUE | TRUE | HP:0011461 | GRCh37 | Pre_NX283_m_Index_201208.g.vcf.gz | WES |
| FAM27 | FAM27 | 13Moe_5Pre_NX283_v_Moeder_201208 | | | female | FALSE | FALSE | | GRCh37 | e_5Pre_NX283_v_Moeder_201208.g.vcf.gz | WES |
| FAM27 | FAM27 | 13Va_5Pre_NX283_m_Vader_201208 | | | male | FALSE | FALSE | | GRCh37 | 5Pre_NX283_m_Vader_201208.g.vcf.gz | WES |
| FAM28 | FAM28 | 14I_5Pre_NX285_m_Index_201208 | 14Va_5Pre_NX285_m_Vader_201208 | 14Moe_5Pre_NX285_v_Moeder_201208 | male | TRUE | TRUE | HP:0011461 | GRCh37 | Pre_NX285_m_Index_201208.g.vcf.gz | WES |
| FAM28 | FAM28 | 14Moe_5Pre_NX285_v_Moeder_201208 | | | female | FALSE | FALSE | | GRCh37 | e_5Pre_NX285_v_Moeder_201208.g.vcf.gz | WES |
| FAM28 | FAM28 | 14Va_5Pre_NX285_m_Vader_201208 | | | male | FALSE | FALSE | | GRCh37 | 5Pre_NX285_m_Vader_201208.g.vcf.gz | WES |
| FAM29 | FAM29 | 15I_5Pre_NX285_v_Index_201208 | 15Va_5Pre_NX285_m_Vader_201208 | 15Moe_5Pre_NX285_v_Moeder_201208 | female | TRUE | TRUE | HP:0011461 | GRCh37 | Pre_NX285_v_Index_201208.g.vcf.gz | WES |
| FAM29 | FAM29 | 15Moe_5Pre_NX285_v_Moeder_201208 | | | female | FALSE | FALSE | | GRCh37 | e_5Pre_NX285_v_Moeder_201208.g.vcf.gz | WES |
| FAM29 | FAM29 | 15Va_5Pre_NX285_m_Vader_201208 | | | male | FALSE | FALSE | | GRCh37 | 5Pre_NX285_m_Vader_201208.g.vcf.gz | WES |

| | | | | | | | | | | | |
|---|---|---|---|---|---|---|---|---|---|---|---|
| FAM3 | FAM3 | 4I_OA_NX307_v_Index_201204 | 4Va_OA_NX307_m_Vader_201204 | 4Moe_OA_NX307_v_Moeder_201204 | female | TRUE | TRUE | HP:0002267, HP:0000750, HP:0001250 | GRCh37 | _NX307_v_Index_201204.g.vcf.gz | WES |
| FAM3 | FAM3 | 4Moe_OA_NX307_v_Moeder_201204 | | | female | FALSE | FALSE | | GRCh37 | _OA_NX307_v_Moeder_201204.g.vcf.gz | WES |
| FAM3 | FAM3 | 4Va_OA_NX307_m_Vader_201204 | | | male | FALSE | FALSE | | GRCh37 | OA_NX307_m_Vader_201204.g.vcf.gz | WES |
| FAM30 | FAM30 | 16I_5Pre_NX285_m_Index_201208 | 16Va_5Pre_NX285_m_Vader_201208 | 16Moe_5Pre_NX285_v_Moeder_201208 | male | TRUE | TRUE | HP:0011461, HP:0001629, HP:0001680 | GRCh37 | Pre_NX285_m_Index_201208.g.vcf.gz | WES |
| FAM30 | FAM30 | 16Moe_5Pre_NX285_v_Moeder_201208 | | | female | FALSE | FALSE | | GRCh37 | e_5Pre_NX285_v_Moeder_201208.g.vcf.gz | WES |
| FAM30 | FAM30 | 16Va_5Pre_NX285_m_Vader_201208 | | | male | FALSE | FALSE | | GRCh37 | 5Pre_NX285_m_Vader_201208.g.vcf.gz | WES |
| FAM31 | FAM31 | 17I_5Pre_NX285_m_Index_201208 | 17Va_5Pre_NX285_m_Vader_201208 | 17Moe_5Pre_NX285_v_Moeder_201208 | male | TRUE | TRUE | HP:0011461, HP:0002744 | GRCh37 | Pre_NX285_m_Index_201208.g.vcf.gz | WES |
| FAM31 | FAM31 | 17Moe_5Pre_NX285_v_Moeder_201208 | | | female | FALSE | FALSE | | GRCh37 | e_5Pre_NX285_v_Moeder_201208.g.vcf.gz | WES |
| FAM31 | FAM31 | 17Va_5Pre_NX285_m_Vader_201208 | | | male | TRUE | FALSE | | GRCh37 | 5Pre_NX285_m_Vader_201208.g.vcf.gz | WES |

| | | | | | | | | | | | |
|---|---|---|---|---|---|---|---|---|---|---|---|
| FAM32 | FAM32 | 18I_5Pre_NX283_m_Index_201208 | 18Va_5Pre_NX283_m_Vader_201208 | 18Moe_5Pre_NX283_v_Moeder_201208 | male | TRUE | TRUE | [illegible] HP:0001274, HP:0010557, HP:0001558, HP:0001776, HP:0000278, | GRCh37 | /path_to/18I_5Pre_NX283_m_Index_201208.g.vcf.gz | WES |
| FAM32 | FAM32 | 18Moe_5Pre_NX283_v_Moeder_201208 | | | female | FALSE | FALSE | | GRCh37 | e_5Pre_NX283_v_Moeder_201208.g.vcf.gz | WES |
| FAM32 | FAM32 | 18Va_5Pre_NX283_m_Vader_201208 | | | male | FALSE | FALSE | | GRCh37 | 5Pre_NX283_m_Vader_201208.g.vcf.gz | WES |
| FAM33 | FAM33 | 19I_5Pre_NX285_m_Index_201208 | 19Va_5Pre_NX285_m_Vader_201208 | 19Moe_5Pre_NX285_v_Moeder_201208 | male | TRUE | TRUE | HP:0011461 | GRCh37 | Pre_NX285_m_Index_201208.g.vcf.gz | WES |
| FAM33 | FAM33 | 19Moe_5Pre_NX285_v_Moeder_201208 | | | female | FALSE | FALSE | | GRCh37 | e_5Pre_NX285_v_Moeder_201208.g.vcf.gz | WES |
| FAM33 | FAM33 | 19Va_5Pre_NX285_m_Vader_201208 | | | male | FALSE | FALSE | | GRCh37 | 5Pre_NX285_m_Vader_201208.g.vcf.gz | WES |
| FAM34 | FAM34 | 20I_5Pre_NX285_m_Index_201208 | 20Va_5Pre_NX285_m_Vader_201208 | 20Moe_5Pre_NX285_v_Moeder_201208 | male | TRUE | TRUE | [illegible] HP:0001195, HP:0010943, HP:0001669, HP:0000316, HP:0000047 | GRCh37 | /path_to/20I_5Pre_NX285_m_Index_201208.g.vcf.gz | WES |
| FAM34 | FAM34 | 20Moe_5Pre_NX285_v_Moeder_201208 | | | female | FALSE | FALSE | | GRCh37 | e_5Pre_NX285_v_Moeder_201208.g.vcf.gz | WES |

| | | | | | | | | | | | |
|---|---|---|---|---|---|---|---|---|---|---|---|
| FAM34 | FAM34 | 20Va_5Pre_NX285_m_Vader_201208 | | | male | FALSE | FALSE | | GRCh37 | 5Pre_NX285_m_Vader_201208.g.vcf.gz | WES |
| FAM35 | FAM35 | 21I_5Pre_NX283_m_Index_201208 | 21Va_5Pre_NX283_m_Vader_201208 | 21Moe_5Pre_NX283_v_Moeder_201208 | male | TRUE | TRUE | [illegible] HP:0002980, HP:0003026, HP:0000772, HP:0002682, HP:0002119, | GRCh37 | /path_to/21I_5Pre_NX283_m_Index_201208.g.vcf.gz | WES |
| FAM35 | FAM35 | 21Moe_5Pre_NX283_v_Moeder_201208 | | | female | FALSE | FALSE | | GRCh37 | e_5Pre_NX283_v_Moeder_201208.g.vcf.gz | WES |
| FAM35 | FAM35 | 21Va_5Pre_NX283_m_Vader_201208 | | | male | FALSE | FALSE | | GRCh37 | 5Pre_NX283_m_Vader_201208.g.vcf.gz | WES |
| FAM36 | FAM36 | 22I_5Pre_NX283_m_Index_201208 | 22Va_5Pre_NX283_m_Vader_201208 | 22Moe_5Pre_NX283_v_Moeder_201208 | male | TRUE | TRUE | HP:0011461, HP:0001776 | GRCh37 | Pre_NX283_m_Index_201208.g.vcf.gz | WES |
| FAM36 | FAM36 | 22Moe_5Pre_NX283_v_Moeder_201208 | | | female | FALSE | FALSE | | GRCh37 | e_5Pre_NX283_v_Moeder_201208.g.vcf.gz | WES |
| FAM36 | FAM36 | 22Va_5Pre_NX283_m_Vader_201208 | | | male | FALSE | FALSE | | GRCh37 | 5Pre_NX283_m_Vader_201208.g.vcf.gz | WES |
| FAM37 | FAM37 | 23I_5Pre_NX285_m_Index_201208 | 23Va_5Pre_NX285_m_Vader_201208 | 23Moe_5Pre_NX285_v_Moeder_201208 | male | TRUE | TRUE | HP:0011461, HP:0011546, HP:0012303 | GRCh37 | Pre_NX285_m_Index_201208.g.vcf.gz | WES |

| | | | | | | | | | | | |
|---|---|---|---|---|---|---|---|---|---|---|---|
| FAM37 | FAM37 | 23Moe_5Pre_NX285_v_Moeder_201208 | | | female | FALSE | FALSE | | GRCh37 | e_5Pre_NX285_v_Moeder_201208.g.vcf.gz | WES |
| FAM37 | FAM37 | 23Va_5Pre_NX285_m_Vader_201208 | | | male | FALSE | FALSE | | GRCh37 | 5Pre_NX285_m_Vader_201208.g.vcf.gz | WES |
| FAM38 | FAM38 | 24I_5Pre_NX285_v_Index_201208 | 24Va_5Pre_NX285_m_Vader_201208 | 24Moe_5Pre_NX285_v_Moeder_201208 | female | TRUE | TRUE | HP:0011461, HP:0010310, HP:0001631 | GRCh37 | Pre_NX285_v_Index_201208.g.vcf.gz | WES |
| FAM38 | FAM38 | 24Moe_5Pre_NX285_v_Moeder_201208 | | | female | FALSE | FALSE | | GRCh37 | e_5Pre_NX285_v_Moeder_201208.g.vcf.gz | WES |
| FAM38 | FAM38 | 24Va_5Pre_NX285_m_Vader_201208 | | | male | FALSE | FALSE | | GRCh37 | 5Pre_NX285_m_Vader_201208.g.vcf.gz | WES |
| FAM39 | FAM39 | 25I_5Pre_NX283_v_Index_201208 | 25Va_5Pre_NX283_m_Vader_201208 | 25Moe_5Pre_NX283_v_Moeder_201208 | female | TRUE | TRUE | HP:0011428, HP:0001539, HP:0000474 | GRCh37 | Pre_NX283_v_Index_201208.g.vcf.gz | WES |
| FAM39 | FAM39 | 25Moe_5Pre_NX283_v_Moeder_201208 | | | female | FALSE | FALSE | | GRCh37 | e_5Pre_NX283_v_Moeder_201208.g.vcf.gz | WES |
| FAM39 | FAM39 | 25Va_5Pre_NX283_m_Vader_201208 | | | male | FALSE | FALSE | | GRCh37 | 5Pre_NX283_m_Vader_201208.g.vcf.gz | WES |
| FAM4 | FAM4 | 5I_OA_NX307_m_Index_201204 | 5Va_OA_NX307_m_Vader_201204 | 5Moe_OA_NX307_v_Moeder_201204 | male | TRUE | TRUE | HP:0001249, HP:0000545, HP:0001007 | GRCh37 | _NX307_m_Index_201204.g.vcf.gz | WES |

| | | | | | | | | | | | |
|---|---|---|---|---|---|---|---|---|---|---|---|
| FAM4 | FAM4 | 5Moe_OA_NX307_v_Moeder_201204 | | | female | FALSE | FALSE | | GRCh37 | _OA_NX307_v_Moeder_201204.g.vcf.gz | WES |
| FAM4 | FAM4 | 5Va_OA_NX307_m_Vader_201204 | | | male | FALSE | FALSE | | GRCh37 | OA_NX307_m_Vader_201204.g.vcf.gz | WES |
| FAM40 | FAM40 | 1I_5Post_NX284_v_Index_201208 | 1Va_5Post_NX284_m_Vader_201208 | 1Moe_5Post_NX284_v_Moeder_201208 | female | TRUE | TRUE | HP:0010948, HP:0001642, HP:0005948 | GRCh37 | ost_NX284_v_Index_201208.g.vcf.gz | WES |
| FAM40 | FAM40 | 1Moe_5Post_NX284_v_Moeder_201208 | | | female | FALSE | FALSE | | GRCh37 | _5Post_NX284_v_Moeder_201208.g.vcf.gz | WES |
| FAM40 | FAM40 | 1Va_5Post_NX284_m_Vader_201208 | | | male | FALSE | FALSE | | GRCh37 | Post_NX284_m_Vader_201208.g.vcf.gz | WES |
| FAM41 | FAM41 | 2I_5Post_NX284_v_Index_201208 | 2Va_5Post_NX284_m_Vader_201208 | 2Moe_5Post_NX284_v_Moeder_201208 | female | TRUE | TRUE | HP:0005107, HP:0002023, HP:0001671 | GRCh37 | ost_NX284_v_Index_201208.g.vcf.gz | WES |
| FAM41 | FAM41 | 2Moe_5Post_NX284_v_Moeder_201208 | | | female | FALSE | FALSE | | GRCh37 | _5Post_NX284_v_Moeder_201208.g.vcf.gz | WES |
| FAM41 | FAM41 | 2Va_5Post_NX284_m_Vader_201208 | | | male | FALSE | FALSE | | GRCh37 | Post_NX284_m_Vader_201208.g.vcf.gz | WES |
| FAM42 | FAM42 | 3I_5Post_NX284_m_Index_201208 | 3Va_5Post_NX284_m_Vader_201208 | 3Moe_5Post_NX284_v_Moeder_201208 | male | TRUE | TRUE | HP:0002023 | GRCh37 | ost_NX284_m_Index_201208.g.vcf.gz | WES |

| | | | | | | | | | | | |
|---|---|---|---|---|---|---|---|---|---|---|---|
| FAM42 | FAM42 | 3Moe_5Post_NX284_v_Moeder_201208 | | | female | FALSE | FALSE | | GRCh37 | _5Post_NX284_v_Moeder_201208.g.vcf.gz | WES |
| FAM42 | FAM42 | 3Va_5Post_NX284_m_Vader_201208 | | | male | FALSE | FALSE | | GRCh37 | Post_NX284_m_Vader_201208.g.vcf.gz | WES |
| FAM43 | FAM43 | 4I_5Post_NX284_m_Index_201208 | 4Va_5Post_NX284_m_Vader_201208 | 4Moe_5Post_NX284_v_Moeder_201208 | male | TRUE | TRUE | HP:0000185, HP:0001321 | GRCh37 | ost_NX284_m_Index_201208.g.vcf.gz | WES |
| FAM43 | FAM43 | 4Moe_5Post_NX284_v_Moeder_201208 | | | female | FALSE | FALSE | | GRCh37 | _5Post_NX284_v_Moeder_201208.g.vcf.gz | WES |
| FAM43 | FAM43 | 4Va_5Post_NX284_m_Vader_201208 | | | male | FALSE | FALSE | | GRCh37 | Post_NX284_m_Vader_201208.g.vcf.gz | WES |
| FAM44 | FAM44 | 5I_5Post_NX284_m_Index_201208 | 5Va_5Post_NX284_m_Vader_201208 | 5Moe_5Post_NX284_v_Moeder_201208 | male | TRUE | TRUE | HP:0001396 | GRCh37 | ost_NX284_m_Index_201208.g.vcf.gz | WES |
| FAM44 | FAM44 | 5Moe_5Post_NX284_v_Moeder_201208 | | | female | FALSE | FALSE | | GRCh37 | _5Post_NX284_v_Moeder_201208.g.vcf.gz | WES |
| FAM44 | FAM44 | 5Va_5Post_NX284_m_Vader_201208 | | | male | FALSE | FALSE | | GRCh37 | Post_NX284_m_Vader_201208.g.vcf.gz | WES |
| FAM45 | FAM45 | 6I_5Post_NX284_m_Index_201208 | 6Va_5Post_NX284_m_Vader_201208 | 6Moe_5Post_NX284_v_Moeder_201208 | male | TRUE | TRUE | HP:0005160 | GRCh37 | ost_NX284_m_Index_201208.g.vcf.gz | WES |

| | | | | | | | | | | | |
|---|---|---|---|---|---|---|---|---|---|---|---|
| FAM45 | FAM45 | 6Moe_5Post_NX284_v_Moeder_201208 | | | female | FALSE | FALSE | | GRCh37 | _5Post_NX284_v_Moeder_201208.g.vcf.gz | WES |
| FAM45 | FAM45 | 6Va_5Post_NX284_m_Vader_201208 | | | male | FALSE | FALSE | | GRCh37 | Post_NX284_m_Vader_201208.g.vcf.gz | WES |
| FAM46 | FAM46 | 7I_5Post_NX284_v_Index_201208 | 7Va_5Post_NX284_m_Vader_201208 | 7Moe_5Post_NX284_v_Moeder_201208 | female | TRUE | TRUE | HP:0002180 | GRCh37 | ost_NX284_v_Index_201208.g.vcf.gz | WES |
| FAM46 | FAM46 | 7Moe_5Post_NX284_v_Moeder_201208 | | | female | FALSE | FALSE | | GRCh37 | _5Post_NX284_v_Moeder_201208.g.vcf.gz | WES |
| FAM46 | FAM46 | 7Va_5Post_NX284_m_Vader_201208 | | | male | FALSE | FALSE | | GRCh37 | Post_NX284_m_Vader_201208.g.vcf.gz | WES |
| FAM47 | FAM47 | 8I_5Post_NX284_m_Index_201208 | 8Va_5Post_NX284_m_Vader_201208 | 8Moe_5Post_NX284_v_Moeder_201208 | male | TRUE | TRUE | HP:0006554 | GRCh37 | ost_NX284_m_Index_201208.g.vcf.gz | WES |
| FAM47 | FAM47 | 8Moe_5Post_NX284_v_Moeder_201208 | | | female | FALSE | FALSE | | GRCh37 | _5Post_NX284_v_Moeder_201208.g.vcf.gz | WES |
| FAM47 | FAM47 | 8Va_5Post_NX284_m_Vader_201208 | | | male | FALSE | FALSE | | GRCh37 | Post_NX284_m_Vader_201208.g.vcf.gz | WES |
| FAM48 | FAM48 | 9I_5Post_NX284_v_Index_201208 | 9Va_5Post_NX284_m_Vader_201208 | 9Moe_5Post_NX284_v_Moeder_201208 | female | TRUE | TRUE | HP:0002093 | GRCh37 | ost_NX284_v_Index_201208.g.vcf.gz | WES |

| | | | | | | | | | | | |
|---|---|---|---|---|---|---|---|---|---|---|---|
| FAM48 | FAM48 | 9Moe_5Post_NX284_v_Moeder_201208 | | | female | FALSE | FALSE | | GRCh37 | _5Post_NX284_v_Moeder_201208.g.vcf.gz | WES |
| FAM48 | FAM48 | 9Va_5Post_NX284_m_Vader_201208 | | | male | FALSE | FALSE | | GRCh37 | Post_NX284_m_Vader_201208.g.vcf.gz | WES |
| FAM49 | FAM49 | 10I_5Post_NX284_m_Index_201208 | 10Va_5Post_NX284_m_Vader_201208 | 10Moe_5Post_NX284_v_Moeder_201208 | male | TRUE | TRUE | HP:0001342 | GRCh37 | Post_NX284_m_Index_201208.g.vcf.gz | WES |
| FAM49 | FAM49 | 10Moe_5Post_NX284_v_Moeder_201208 | | | female | FALSE | FALSE | | GRCh37 | e_5Post_NX284_v_Moeder_201208.g.vcf.gz | WES |
| FAM49 | FAM49 | 10Va_5Post_NX284_m_Vader_201208 | | | male | FALSE | FALSE | | GRCh37 | 5Post_NX284_m_Vader_201208.g.vcf.gz | WES |
| FAM5 | FAM5 | 6I_OA_NX307_m_Index_201204 | 6Va_OA_NX307_m_Vader_201204 | 6Moe_OA_NX307_v_Moeder_201204 | male | TRUE | TRUE | HP:0012759, HP:0000708, HP:0000384 | GRCh37 | _NX307_m_Index_201204.g.vcf.gz | WES |
| FAM5 | FAM5 | 6Moe_OA_NX307_v_Moeder_201204 | | | female | FALSE | FALSE | | GRCh37 | _OA_NX307_v_Moeder_201204.g.vcf.gz | WES |
| FAM5 | FAM5 | 6Va_OA_NX307_m_Vader_201204 | | | male | FALSE | FALSE | | GRCh37 | OA_NX307_m_Vader_201204.g.vcf.gz | WES |
| FAM50 | FAM50 | 11I_5Post_NX284_m_Index_201208 | 11Va_5Post_NX284_m_Vader_201208 | 11Moe_5Post_NX284_v_Moeder_201208 | male | TRUE | TRUE | HP:0005160 | GRCh37 | Post_NX284_m_Index_201208.g.vcf.gz | WES |

| | | | | | | | | | | | |
|---|---|---|---|---|---|---|---|---|---|---|---|
| FAM50 | FAM50 | 11Moe_5Post_NX284_v_Moeder_201208 | | | female | FALSE | FALSE | | GRCh37 | e_5Post_NX284_v_Moeder_201208.g.vcf.gz | WES |
| FAM50 | FAM50 | 11Va_5Post_NX284_m_Vader_201208 | | | male | FALSE | FALSE | | GRCh37 | 5Post_NX284_m_Vader_201208.g.vcf.gz | WES |
| FAM51 | FAM51 | 12I_5Post_NX284_v_Index_201208 | 12Va_5Post_NX284_m_Vader_201208 | 12Moe_5Post_NX284_v_Moeder_201208 | female | TRUE | TRUE | HP:0000202 | GRCh37 | Post_NX284_v_Index_201208.g.vcf.gz | WES |
| FAM51 | FAM51 | 12Moe_5Post_NX284_v_Moeder_201208 | | | female | FALSE | FALSE | | GRCh37 | e_5Post_NX284_v_Moeder_201208.g.vcf.gz | WES |
| FAM51 | FAM51 | 12Va_5Post_NX284_m_Vader_201208 | | | male | FALSE | FALSE | | GRCh37 | 5Post_NX284_m_Vader_201208.g.vcf.gz | WES |
| FAM52 | FAM52 | 13I_5Post_NX286_v_Index_201208 | 13Va_5Post_NX286_m_Vader_201208 | 13Moe_5Post_NX286_v_Moeder_201208 | female | TRUE | TRUE | HP:0001789 | GRCh37 | Post_NX286_v_Index_201208.g.vcf.gz | WES |
| FAM52 | FAM52 | 13Moe_5Post_NX286_v_Moeder_201208 | | | female | FALSE | FALSE | | GRCh37 | e_5Post_NX286_v_Moeder_201208.g.vcf.gz | WES |
| FAM52 | FAM52 | 13Va_5Post_NX286_m_Vader_201208 | | | male | FALSE | FALSE | | GRCh37 | 5Post_NX286_m_Vader_201208.g.vcf.gz | WES |
| FAM53 | FAM53 | 14I_5Post_NX284_v_Index_201208 | 14Va_5Post_NX284_m_Vader_201208 | 14Moe_5Post_NX284_v_Moeder_201208 | female | TRUE | TRUE | HP:0002205 | GRCh37 | Post_NX284_v_Index_201208.g.vcf.gz | WES |

| | | | | | | | | | | | |
|---|---|---|---|---|---|---|---|---|---|---|---|
| FAM53 | FAM53 | 14Moe_5Post_NX284_v_Moeder_201208 | | | female | FALSE | FALSE | | GRCh37 | e_5Post_NX284_v_Moeder_201208.g.vcf.gz | WES |
| FAM53 | FAM53 | 14Va_5Post_NX284_m_Vader_201208 | | | male | FALSE | FALSE | | GRCh37 | 5Post_NX284_m_Vader_201208.g.vcf.gz | WES |
| FAM54 | FAM54 | 15I_5Post_NX284_m_Index_201208 | 15Va_5Post_NX284_m_Vader_201208 | 15Moe_5Post_NX284_v_Moeder_201208 | male | TRUE | TRUE | HP:0001647, HP:0000239, HP:0004322 | GRCh37 | Post_NX284_m_Index_201208.g.vcf.gz | WES |
| FAM54 | FAM54 | 15Moe_5Post_NX284_v_Moeder_201208 | | | female | FALSE | FALSE | | GRCh37 | e_5Post_NX284_v_Moeder_201208.g.vcf.gz | WES |
| FAM54 | FAM54 | 15Va_5Post_NX284_m_Vader_201208 | | | male | FALSE | FALSE | | GRCh37 | 5Post_NX284_m_Vader_201208.g.vcf.gz | WES |
| FAM55 | FAM55 | 16I_5Post_NX286_v_Index_201208 | 16Va_5Post_NX286_m_Vader_201208 | 16Moe_5Post_NX286_v_Moeder_201208 | female | TRUE | TRUE | HP:0002093 | GRCh37 | Post_NX286_v_Index_201208.g.vcf.gz | WES |
| FAM55 | FAM55 | 16Moe_5Post_NX286_v_Moeder_201208 | | | female | FALSE | FALSE | | GRCh37 | e_5Post_NX286_v_Moeder_201208.g.vcf.gz | WES |
| FAM55 | FAM55 | 16Va_5Post_NX286_m_Vader_201208 | | | male | FALSE | FALSE | | GRCh37 | 5Post_NX286_m_Vader_201208.g.vcf.gz | WES |
| FAM56 | FAM56 | 17I_5Post_NX286_m_Index_201208 | 17Va_5Post_NX286_m_Vader_201208 | 17Moe_5Post_NX286_v_Moeder_201208 | male | TRUE | TRUE | HP:0001644 | GRCh37 | Post_NX286_m_Index_201208.g.vcf.gz | WES |

| | | | | | | | | | | | |
|---|---|---|---|---|---|---|---|---|---|---|---|
| FAM56 | FAM56 | 17Moe_5Post_NX286_v_Moeder_201208 | | | female | FALSE | FALSE | | GRCh37 | e_5Post_NX286_v_Moeder_201208.g.vcf.gz | WES |
| FAM56 | FAM56 | 17Va_5Post_NX286_m_Vader_201208 | | | male | FALSE | FALSE | | GRCh37 | 5Post_NX286_m_Vader_201208.g.vcf.gz | WES |
| FAM57 | FAM57 | 18I_5Post_NX284_m_Index_201208 | 18Va_5Post_NX284_m_Vader_201208 | 18Moe_5Post_NX284_v_Moeder_201208 | male | TRUE | TRUE | HP:0008207 | GRCh37 | Post_NX284_m_Index_201208.g.vcf.gz | WES |
| FAM57 | FAM57 | 18Moe_5Post_NX284_v_Moeder_201208 | | | female | FALSE | FALSE | | GRCh37 | e_5Post_NX284_v_Moeder_201208.g.vcf.gz | WES |
| FAM57 | FAM57 | 18Va_5Post_NX284_m_Vader_201208 | | | male | FALSE | FALSE | | GRCh37 | 5Post_NX284_m_Vader_201208.g.vcf.gz | WES |
| FAM58 | FAM58 | 19I_5Post_NX284_m_Index_201208 | 19Va_5Post_NX284_m_Vader_201208 | 19Moe_5Post_NX284_v_Moeder_201208 | male | TRUE | TRUE | HP:0005562 | GRCh37 | Post_NX284_m_Index_201208.g.vcf.gz | WES |
| FAM58 | FAM58 | 19Moe_5Post_NX284_v_Moeder_201208 | | | female | FALSE | FALSE | | GRCh37 | e_5Post_NX284_v_Moeder_201208.g.vcf.gz | WES |
| FAM58 | FAM58 | 19Va_5Post_NX284_m_Vader_201208 | | | male | FALSE | FALSE | | GRCh37 | 5Post_NX284_m_Vader_201208.g.vcf.gz | WES |
| FAM59 | FAM59 | 20I_5Post_NX284_m_Index_201208 | 20Va_5Post_NX284_m_Vader_201208 | 20Moe_5Post_NX284_v_Moeder_201208 | male | TRUE | TRUE | HP:0001263 | GRCh37 | Post_NX284_m_Index_201208.g.vcf.gz | WES |

| | | | | | | | | | | | |
|---|---|---|---|---|---|---|---|---|---|---|---|
| FAM59 | FAM59 | 20Moe_5Post_NX284_v_Moeder_201208 | | | female | FALSE | FALSE | | GRCh37 | e_5Post_NX284_v_Moeder_201208.g.vcf.gz | WES |
| FAM59 | FAM59 | 20Va_5Post_NX284_m_Vader_201208 | | | male | FALSE | FALSE | | GRCh37 | 5Post_NX284_m_Vader_201208.g.vcf.gz | WES |
| FAM6 | FAM6 | 7I_OA_NX307_m_Index_201204 | 7Va_OA_NX307_m_Vader_201204 | 7Moe_OA_NX307_v_Moeder_201204 | male | TRUE | TRUE | HP:0000750, HP:0000708, HP:0012759 | GRCh37 | _NX307_m_Index_201204.g.vcf.gz | WES |
| FAM6 | FAM6 | 7Moe_OA_NX307_v_Moeder_201204 | | | female | FALSE | FALSE | | GRCh37 | _OA_NX307_v_Moeder_201204.g.vcf.gz | WES |
| FAM6 | FAM6 | 7Va_OA_NX307_m_Vader_201204 | | | male | FALSE | FALSE | | GRCh37 | OA_NX307_m_Vader_201204.g.vcf.gz | WES |
| FAM60 | FAM60 | 21I_5Post_NX284_v_Index_201208 | 21Va_5Post_NX284_m_Vader_201208 | 21Moe_5Post_NX284_v_Moeder_201208 | female | TRUE | TRUE | HP:0001520, HP:0001319 | GRCh37 | Post_NX284_v_Index_201208.g.vcf.gz | WES |
| FAM60 | FAM60 | 21Moe_5Post_NX284_v_Moeder_201208 | | | female | FALSE | FALSE | | GRCh37 | e_5Post_NX284_v_Moeder_201208.g.vcf.gz | WES |
| FAM60 | FAM60 | 21Va_5Post_NX284_m_Vader_201208 | | | male | FALSE | FALSE | | GRCh37 | 5Post_NX284_m_Vader_201208.g.vcf.gz | WES |
| FAM61 | FAM61 | 22I_5Post_NX284_m_Index_201208 | 22Va_5Post_NX284_m_Vader_201208 | 22Moe_5Post_NX284_v_Moeder_201208 | male | TRUE | TRUE | HP:0001298 | GRCh37 | Post_NX284_m_Index_201208.g.vcf.gz | WES |

| | | | | | | | | | | | |
|---|---|---|---|---|---|---|---|---|---|---|---|
| FAM61 | FAM61 | 22Moe_5Post_NX284_v_Moeder_201208 | | | female | FALSE | FALSE | | GRCh37 | e_5Post_NX284_v_Moeder_201208.g.vcf.gz | WES |
| FAM61 | FAM61 | 22Va_5Post_NX284_m_Vader_201208 | | | male | FALSE | FALSE | | GRCh37 | 5Post_NX284_m_Vader_201208.g.vcf.gz | WES |
| FAM62 | FAM62 | 23I_5Post_NX284_m_Index_201208 | 23Va_5Post_NX284_m_Vader_201208 | 23Moe_5Post_NX284_v_Moeder_201208 | male | TRUE | TRUE | HP:0000239, HP:0001798 | GRCh37 | Post_NX284_m_Index_201208.g.vcf.gz | WES |
| FAM62 | FAM62 | 23Moe_5Post_NX284_v_Moeder_201208 | | | female | FALSE | FALSE | | GRCh37 | e_5Post_NX284_v_Moeder_201208.g.vcf.gz | WES |
| FAM62 | FAM62 | 23Va_5Post_NX284_m_Vader_201208 | | | male | FALSE | FALSE | | GRCh37 | 5Post_NX284_m_Vader_201208.g.vcf.gz | WES |
| FAM63 | FAM63 | 24I_5Post_NX284_v_Index_201208 | 24Va_5Post_NX284_m_Vader_201208 | 24Moe_5Post_NX284_v_Moeder_201208 | female | TRUE | TRUE | HP:0007359, HP:0001684, HP:0011623 | GRCh37 | Post_NX284_v_Index_201208.g.vcf.gz | WES |
| FAM63 | FAM63 | 24Moe_5Post_NX284_v_Moeder_201208 | | | female | FALSE | FALSE | | GRCh37 | e_5Post_NX284_v_Moeder_201208.g.vcf.gz | WES |
| FAM63 | FAM63 | 24Va_5Post_NX284_m_Vader_201208 | | | male | FALSE | FALSE | | GRCh37 | 5Post_NX284_m_Vader_201208.g.vcf.gz | WES |
| FAM64 | FAM64 | 25I_5Post_NX286_v_Index_201208 | 25Va_5Post_NX286_m_Vader_201208 | 25Moe_5Post_NX286_v_Moeder_201208 | female | TRUE | TRUE | HP:0003473, HP:0005235, HP:0011675 | GRCh37 | Post_NX286_v_Index_201208.g.vcf.gz | WES |

| | | | | | | | | | | | |
|---|---|---|---|---|---|---|---|---|---|---|---|
| FAM64 | FAM64 | 25Moe_5Post_NX286_v_Moeder_201208 | | | female | FALSE | FALSE | | GRCh37 | e_5Post_NX286_v_Moeder_201208.g.vcf.gz | WES |
| FAM64 | FAM64 | 25Va_5Post_NX286_m_Vader_201208 | | | male | FALSE | FALSE | | GRCh37 | 5Post_NX286_m_Vader_201208.g.vcf.gz | WES |
| FAM7 | FAM7 | 8I_OA_NX307_m_Index_201204 | 8Va_OA_NX307_m_Vader_201204 | 8Moe_OA_NX307_v_Moeder_201204 | male | TRUE | TRUE | HP:0000708, HP:0000750 | GRCh37 | _NX307_m_Index_201204.g.vcf.gz | WES |
| FAM7 | FAM7 | 8Moe_OA_NX307_v_Moeder_201204 | | | female | FALSE | FALSE | | GRCh37 | _OA_NX307_v_Moeder_201204.g.vcf.gz | WES |
| FAM7 | FAM7 | 8Va_OA_NX307_m_Vader_201204 | | | male | FALSE | FALSE | | GRCh37 | OA_NX307_m_Vader_201204.g.vcf.gz | WES |
| FAM8 | FAM8 | 9I_OA_NX307_m_Index_201204 | 9Va_OA_NX307_m_Vader_201204 | 9Moe_OA_NX307_v_Moeder_201204 | male | TRUE | TRUE | HP:0012759, HP:0007018, HP:0000729 | GRCh37 | _NX307_m_Index_201204.g.vcf.gz | WES |
| FAM8 | FAM8 | 9Moe_OA_NX307_v_Moeder_201204 | | | female | FALSE | FALSE | | GRCh37 | _OA_NX307_v_Moeder_201204.g.vcf.gz | WES |
| FAM8 | FAM8 | 9Va_OA_NX307_m_Vader_201204 | | | male | FALSE | FALSE | | GRCh37 | OA_NX307_m_Vader_201204.g.vcf.gz | WES |
| FAM9 | FAM9 | 10I_OA_NX307_v_Index_201204 | 10Va_OA_NX307_m_Vader_201204 | 10Moe_OA_NX307_v_Moeder_201204 | female | TRUE | TRUE | HP:0002463, HP:0000708, HP:0000098, HP:0001548 | GRCh37 | /path_to/10I_OA_NX307_v_Index_201204.g.vcf.gz | WES |

| | | | | | | | | | | | |
|---|---|---|---|---|---|---|---|---|---|---|---|
| FAM9 | FAM9 | 10Moe_OA_NX307_v_Moeder_201204 | | | female | FALSE | FALSE | | GRCh37 | e_OA_NX307_v_Moeder_201204.g.vcf.gz | WES |
| FAM9 | FAM9 | 10Va_OA_NX307_m_Vader_201204 | | | male | FALSE | FALSE | | GRCh37 | OA_NX307_m_Vader_201204.g.vcf.gz | WES |
| FAM90 | FAM90 | 1I_kDCM_NX288_m_Index_201203 | 1Va_kDCM_NX288_m_Vader_201203 | 1Moe_kDCM_NX288_v_Moeder_201203 | male | TRUE | TRUE | HP:0001319, HP:0004303, HP:0001397 | GRCh37 | CM_NX288_m_Index_201203.g.vcf.gz | WES |
| FAM90 | FAM90 | 1Moe_kDCM_NX288_v_Moeder_201203 | | | female | FALSE | FALSE | | GRCh37 | _kDCM_NX288_v_Moeder_201203.g.vcf.gz | WES |
| FAM90 | FAM90 | 1Va_kDCM_NX288_m_Vader_201203 | | | male | FALSE | FALSE | | GRCh37 | DCM_NX288_m_Vader_201203.g.vcf.gz | WES |
| FAM91 | FAM91 | 2I_kDCM_NX288_m_Index_201203 | 2Va_kDCM_NX288_m_Vader_201203 | 2Moe_kDCM_NX288_v_Moeder_201203 | male | TRUE | TRUE | HP:0001644, HP:0040196 | GRCh37 | CM_NX288_m_Index_201203.g.vcf.gz | WES |
| FAM91 | FAM91 | 2Moe_kDCM_NX288_v_Moeder_201203 | | | female | FALSE | FALSE | | GRCh37 | /path_to/2Moe_kDCM_NX288_v_Moeder_201203.g.vcf.gz | WES |
| FAM91 | FAM91 | 2Va_kDCM_NX288_m_Vader_201203 | | | male | FALSE | FALSE | | GRCh37 | DCM_NX288_m_Vader_201203.g.vcf.gz | WES |
| FAM92 | FAM92 | 3I_kDCM_NX288_v_Index_201203 | 3Va_kDCM_NX288_m_Vader_201203 | 3Moe_kDCM_NX288_v_Moeder_201203 | female | TRUE | TRUE | HP:0012817, HP:0001688, HP:0001698 | GRCh37 | CM_NX288_v_Index_201203.g.vcf.gz | WES |

| | | | | | | | | | | | |
|---|---|---|---|---|---|---|---|---|---|---|---|
| FAM92 | FAM92 | 3Moe_kDCM_NX288_v_Moeder_201203 | | | female | FALSE | FALSE | | GRCh37 | _kDCM_NX288_v_Moeder_201203.g.vcf.gz | WES |
| FAM92 | FAM92 | 3Va_kDCM_NX288_m_Vader_201203 | | | male | FALSE | FALSE | | GRCh37 | DCM_NX288_m_Vader_201203.g.vcf.gz | WES |
| FAM93 | FAM93 | 4I_kDCM_NX288_m_Index_201203 | 4Va_kDCM_NX288_m_Vader_201203 | 4Moe_kDCM_NX288_v_Moeder_201203 | male | TRUE | TRUE | HP:0012817 | GRCh37 | CM_NX288_m_Index_201203.g.vcf.gz | WES |
| FAM93 | FAM93 | 4Moe_kDCM_NX288_v_Moeder_201203 | | | female | FALSE | FALSE | | GRCh37 | _kDCM_NX288_v_Moeder_201203.g.vcf.gz | WES |
| FAM93 | FAM93 | 4Va_kDCM_NX288_m_Vader_201203 | | | male | FALSE | FALSE | | GRCh37 | DCM_NX288_m_Vader_201203.g.vcf.gz | WES |
| FAM94 | FAM94 | 5I_kDCM_NX288_m_Index_201203 | 5Va_kDCM_NX288_m_Vader_201203 | 5Moe_kDCM_NX288_v_Moeder_201203 | male | TRUE | TRUE | HP:0001644 | GRCh37 | CM_NX288_m_Index_201203.g.vcf.gz | WES |
| FAM94 | FAM94 | 5Moe_kDCM_NX288_v_Moeder_201203 | | | female | FALSE | FALSE | | GRCh37 | _kDCM_NX288_v_Moeder_201203.g.vcf.gz | WES |
| FAM94 | FAM94 | 5Va_kDCM_NX288_m_Vader_201203 | | | male | FALSE | FALSE | | GRCh37 | DCM_NX288_m_Vader_201203.g.vcf.gz | WES |

Supplementary table 5 **Example of sample sheet for routine diagnostics cohort**
