## supplementary table 6 for "MOLGENIS VIP: an open-source and modular pipeline for high-throughput and integrated DNA variant analysis"

| project_id | family_id | individual_id | paternal_id | maternal_id | sex | affected | proband | hpo_ids | assembly | gvcf | sequencing_method |
|---|---|---|---|---|---|---|---|---|---|---|---|
| FAM0000637 | FAM0000637 | E481613 | E558796 | E769834 | male | TRUE | TRUE | HP:0000508,HP:0002015,HP:0003323,HP:0003473,HP:0003691,HP:0008994,HP:0008997 | GRCh37 | /path_to/gvcftools_E481613.g.vcf | WES |
| FAM0000637 | FAM0000637 | E769834 | | | female | FALSE | FALSE | | GRCh37 | /path_to/gvcftools_E769834.g.vcf | WES |
| FAM0000637 | FAM0000637 | E558796 | | | male | FALSE | FALSE | HP:0000508 | GRCh37 | /path_to/gvcftools_E558796.g.vcf | WES |
| FAM0001617 | FAM0001617 | E431798 | E749507 | E163380 | | TRUE | TRUE | HP:0006879 | GRCh37 | /path_to/gvcftools_E431798.g.vcf | WES |
| FAM0001617 | FAM0001617 | E749507 | | | male | FALSE | FALSE | | GRCh37 | /path_to/gvcftools_E749507.g.vcf | WES |
| FAM0001617 | FAM0001617 | E163380 | | | female | FALSE | FALSE | | GRCh37 | /path_to/gvcftools_E163380.g.vcf | WES |

| | | | | | | | | | | | |
|---|---|---|---|---|---|---|---|---|---|---|---|
| FAM0001911 | FAM0001911 | E395076 | E528179 | E715649 | female | TRUE | TRUE | HP:0001252,HP:0001283,HP:0002015,HP:0002804,HP:0003473,HP:0030000,HP:0030192,HP:0030197 | GRCh37 | /path_to/gvcftools_E395076.g.vcf | WES |
| FAM0001911 | FAM0001911 | E715649 | | | female | FALSE | FALSE | | GRCh37 | /path_to/gvcftools_E715649.g.vcf | WES |
| FAM0001911 | FAM0001911 | E528179 | | | male | FALSE | FALSE | | GRCh37 | /path_to/gvcftools_E528179.g.vcf | WES |
| FAM0001919 | FAM0001919 | E989776 | E319195 | E226500 | | TRUE | TRUE | HP:0000483,HP:0000510,HP:0000518,HP:0000519,HP:0000545,HP:0001272,HP:0001288,HP:0002503,HP:0003198,HP:0006895,HP:0100543 | GRCh37 | /path_to/gvcftools_E989776.g.vcf | WES |

| | | | | | | | | | | | |
|---|---|---|---|---|---|---|---|---|---|---|---|
| FAM0001919 | FAM0001919 | E226500 | | | female | TRUE | FALSE | HP:0000518,HP:0001272,HP:0001288,HP:0001695,HP:0002878,HP:0100543 | GRCh37 | /path_to/gvcftools_E226500.g.vcf | WES |
| FAM0001919 | FAM0001919 | E319195 | | | male | FALSE | FALSE | | GRCh37 | /path_to/gvcftools_E319195.g.vcf | WES |
| FAM0006091 | FAM0006091 | E830835 | E952760 | E452470 | female | TRUE | TRUE | HP:0002987,HP:0003306,HP:0003327,HP:0003560,HP:0005750,HP:0006380,HP:0008959,HP:0008981,HP:0008994,HP:0008997,HP:0009053,HP:0100360 | GRCh37 | /path_to/gvcftools_E830835.g.vcf | WES |
| FAM0006091 | FAM0006091 | E952760 | | | male | FALSE | FALSE | | GRCh37 | /path_to/gvcftools_E952760.g.vcf | WES |
| FAM0006091 | FAM0006091 | E452470 | | | female | FALSE | FALSE | | GRCh37 | /path_to/gvcftools_E452470.g.vcf | WES |

| | | | | | | | | | | | |
|---|---|---|---|---|---|---|---|---|---|---|---|
| FAM0008013 | FAM0008013 | E603477 | E883874 | E638210 | male | TRUE | TRUE | HP:0001324,HP:0002151,HP:0003323,HP:0003403,HP:0003473,HP:0003701,HP:0003731,HP:0008994 | GRCh37 | /path_to/gvcftools_E603477.g.vcf | WES |
| FAM0008013 | FAM0008013 | E883874 | | | male | FALSE | FALSE | | GRCh37 | /path_to/gvcftools_E883874.g.vcf | WES |
| FAM0008013 | FAM0008013 | E638210 | | | female | FALSE | FALSE | | GRCh37 | /path_to/gvcftools_E638210.g.vcf | WES |
| FAM0008660 | FAM0008660 | E570269 | E442035 | E176978 | female | TRUE | TRUE | HP:0000252,HP:0000519,HP:0001263 | GRCh37 | /path_to/gvcftools_E570269.g.vcf | WES |
| FAM0008660 | FAM0008660 | E176978 | | | female | FALSE | FALSE | | GRCh37 | /path_to/gvcftools_E176978.g.vcf | WES |
| FAM0008660 | FAM0008660 | E442035 | | | male | FALSE | FALSE | | GRCh37 | /path_to/gvcftools_E442035.g.vcf | WES |

| | | | | | | | | | | | |
|---|---|---|---|---|---|---|---|---|---|---|---|
| FAM0008669 | FAM0008669 | E447588 | E863754 | E092350 | female | TRUE | TRUE | HP:0000020,HP:0000750,HP:0000924,HP:0001256,HP:0001263,HP:0001270,HP:0001310,HP:0001315,HP:0001337,HP:0002080,HP:0002119,HP:0002194,HP:0002345,HP:0002607,HP:0007083,HP:0010862,HP:0011968,HP:0012443,HP:0012759,HP:0025335,HP:0025336,HP:0040183,HP:0100851 | GRCh37 | /path_to/gvcftools_E447588.g.vcf | WES |
| FAM0008669 | FAM0008669 | E092350 | | | female | FALSE | FALSE | | GRCh37 | /path_to/gvcftools_E092350.g.vcf | WES |
| FAM0008669 | FAM0008669 | E863754 | | | male | FALSE | FALSE | | GRCh37 | /path_to/gvcftools_E863754.g.vcf | WES |

| | | | | | | | | | | | |
|---|---|---|---|---|---|---|---|---|---|---|---|
| FAM0009068 | FAM0009068 | E128884 | E849549 | E269678 | female | TRUE | TRUE | HP:0000252,HP:0000639,HP:0001250,HP:0001272,HP:0002131,HP:0002411,HP:0010553 | GRCh37 | /path_to/gvcftools_E128884.g.vcf | WES |
| FAM0009068 | FAM0009068 | E849549 | | | male | FALSE | FALSE | | GRCh37 | /path_to/gvcftools_E849549.g.vcf | WES |
| FAM0009068 | FAM0009068 | E269678 | | | female | FALSE | FALSE | | GRCh37 | /path_to/gvcftools_E269678.g.vcf | WES |
| FAM0009788 | FAM0009788 | E282663 | E822555 | E903395 | male | TRUE | TRUE | HP:0000508,HP:0002515,HP:0003325,HP:0003403,HP:0003473,HP:0008944 | GRCh37 | /path_to/gvcftools_E282663.g.vcf | WES |
| FAM0009788 | FAM0009788 | E822555 | | | male | FALSE | FALSE | | GRCh37 | /path_to/gvcftools_E822555.g.vcf | WES |
| FAM0009788 | FAM0009788 | E903395 | | | female | FALSE | FALSE | | GRCh37 | /path_to/gvcftools_E903395.g.vcf | WES |

| | | | | | | | | | | | |
|---|---|---|---|---|---|---|---|---|---|---|---|
| FAM0010156 | FAM0010156 | E476365 | E471938 | E078489 | male | TRUE | TRUE | HP:0000252,HP:0000399,HP:0000648,HP:0001290,HP:0002187,HP:0002376,HP:0002415,HP:0004322,HP:0004373,HP:0011097,HP:0200134 | GRCh37 | /path_to/gvcftools_E476365.g.vcf | WES |
| FAM0010156 | FAM0010156 | E471938 | | | male | FALSE | FALSE | | GRCh37 | /path_to/gvcftools_E471938.g.vcf | WES |
| FAM0010156 | FAM0010156 | E078489 | | | female | FALSE | FALSE | | GRCh37 | /path_to/gvcftools_E078489.g.vcf | WES |

| | | | | | | | | | | | |
|---|---|---|---|---|---|---|---|---|---|---|---|
| FAM0010164 | FAM0010164 | E062108 | E098556 | E444562 | female | TRUE | TRUE | HP:0000252,HP:0000253,HP:0000286,HP:0000341,HP:0000486,HP:0000545,HP:0000639,HP:0000733,HP:0001251,HP:0001263,HP:0001344,HP:0001511,HP:0001561,HP:0002342,HP:0008071,HP:0009183,HP:0009276 | GRCh37 | /path_to/gvcftools_E062108.g.vcf | WES |
| FAM0010164 | FAM0010164 | E098556 | | | male | FALSE | FALSE | | GRCh37 | /path_to/gvcftools_E098556.g.vcf | WES |
| FAM0010164 | FAM0010164 | E444562 | | | female | FALSE | FALSE | | GRCh37 | /path_to/gvcftools_E444562.g.vcf | WES |
| FAM0010183 | FAM0010183 | E221264 | E565934 | E479521 | male | TRUE | TRUE | HP:0000729,HP:0001249 | GRCh37 | /path_to/gvcftools_E221264.g.vcf | WES |

| | | | | | | | | | | | |
|---|---|---|---|---|---|---|---|---|---|---|---|
| FAM0010183 | FAM0010183 | E479521 | | | female | FALSE | FALSE | | GRCh37 | /path_to/gvcftools_E479521.g.vcf | WES |
| FAM0010183 | FAM0010183 | E565934 | | | male | FALSE | FALSE | | GRCh37 | /path_to/gvcftools_E565934.g.vcf | WES |
| FAM0010185 | FAM0010185 | E594094 | E205347 | E806188 | male | TRUE | TRUE | HP:0000252,HP:0001274,HP:0002269,HP:0007033 | GRCh37 | /path_to/gvcftools_E594094.g.vcf | WES |
| FAM0010185 | FAM0010185 | E205347 | | | male | FALSE | FALSE | | GRCh37 | /path_to/gvcftools_E205347.g.vcf | WES |
| FAM0010185 | FAM0010185 | E806188 | | | female | FALSE | FALSE | | GRCh37 | /path_to/gvcftools_E806188.g.vcf | WES |
| FAM0010234 | FAM0010234 | E339658 | E940878 | E202434 | male | TRUE | TRUE | HP:0000496,HP:0001249,HP:0002179,HP:0010819,HP:0011968 | GRCh37 | /path_to/gvcftools_E339658.g.vcf | WES |
| FAM0010234 | FAM0010234 | E940878 | | | male | FALSE | FALSE | | GRCh37 | /path_to/gvcftools_E940878.g.vcf | WES |
| FAM0010234 | FAM0010234 | E202434 | | | female | FALSE | FALSE | | GRCh37 | /path_to/gvcftools_E202434.g.vcf | WES |
| FAM0010252 | FAM0010252 | E733888 | E342683 | E266043 | male | TRUE | TRUE | HP:0001250,HP:0200134 | GRCh37 | /path_to/gvcftools_E733888.g.vcf | WES |
| FAM0010252 | FAM0010252 | E342683 | | | male | FALSE | FALSE | | GRCh37 | /path_to/gvcftools_E342683.g.vcf | WES |

| | | | | | | | | | | | |
|---|---|---|---|---|---|---|---|---|---|---|---|
| FAM0010252 | FAM0010252 | E266043 | | | female | FALSE | FALSE | | GRCh37 | /path_to/gvcftools_E266043.g.vcf | WES |
| FAM0010960 | FAM0010960 | E484318 | E664672 | E630769 | male | TRUE | TRUE | HP:0000218,HP:0001324,HP:0002058,HP:0002194,HP:0002460,HP:0003202,HP:0003323,HP:0003690,HP:0007149,HP:0008944,HP:0008948,HP:0008956,HP:0009046,HP:0009050,HP:0009063,HP:0009073,HP:0010628,HP:0011399,HP:0430025 | GRCh37 | /path_to/gvcftools_E484318.g.vcf | WES |
| FAM0010960 | FAM0010960 | E664672 | | | male | FALSE | FALSE | | GRCh37 | /path_to/gvcftools_E664672.g.vcf | WES |
| FAM0010960 | FAM0010960 | E630769 | | | female | FALSE | FALSE | | GRCh37 | /path_to/gvcftools_E630769.g.vcf | WES |

| | | | | | | | | | | | |
|---|---|---|---|---|---|---|---|---|---|---|---|
| FAM0011240 | FAM0011240 | E714923 | E254013 | E165708 | male | TRUE | TRUE | HP:0001270,HP:0002015,HP:0002355,HP:0002515,HP:0003388,HP:0003477,HP:0008994,HP:0008997 | GRCh37 | /path_to/gvcftools_E714923.g.vcf | WES |
| FAM0011240 | FAM0011240 | E254013 | | | male | FALSE | FALSE | | GRCh37 | /path_to/gvcftools_E254013.g.vcf | WES |
| FAM0011240 | FAM0011240 | E165708 | | | female | FALSE | FALSE | | GRCh37 | /path_to/gvcftools_E165708.g.vcf | WES |

| | | | | | | | | | | | |
|---|---|---|---|---|---|---|---|---|---|---|---|
| FAM0011274 | FAM0011274 | E023128 | E765800 | E906982 | female | TRUE | TRUE | HP:0000729,HP:0001263,HP:0001511,HP:0001518,HP:0001520,HP:0001545,HP:0001558,HP:0001561,HP:0001562,HP:0001622,HP:0001762,HP:0001787,HP:0004691,HP:0004692,HP:0007900,HP:0008071,HP:0009800,HP:0010519,HP:0011438,HP:0012188,HP:0030244,HP:0100603,HP:0100622 | GRCh37 | /path_to/gvcftools_E023128.g.vcf | WES |
| FAM0011274 | FAM0011274 | E765800 | | | male | FALSE | FALSE | | GRCh37 | /path_to/gvcftools_E765800.g.vcf | WES |
| FAM0011274 | FAM0011274 | E906982 | | | female | FALSE | FALSE | | GRCh37 | /path_to/gvcftools_E906982.g.vcf | WES |

| | | | | | | | | | | | |
|---|---|---|---|---|---|---|---|---|---|---|---|
| FAM0010233 | FAM0010233 | E685615 | E998039 | E335259 | male | TRUE | TRUE | HP:0000324,HP:0000709,HP:0001249,HP:0001250,HP:0001272,HP:0001276,HP:0001999,HP:0002304,HP:0002345,HP:0002493 | GRCh37 | /path_to/gvcftools_E685615.g.vcf | WES |
| FAM0010233 | FAM0010233 | E998039 | | | male | FALSE | FALSE | | GRCh37 | /path_to/gvcftools_E998039.g.vcf | WES |
| FAM0010233 | FAM0010233 | E335259 | | | female | FALSE | FALSE | | GRCh37 | /path_to/gvcftools_E335259.g.vcf | WES |

| | | | | | | | | | | | |
|---|---|---|---|---|---|---|---|---|---|---|---|
| FAM0004372 | FAM0004372 | E261621 | E707589 | E993720 | male | TRUE | TRUE | HP:0000007,HP:0000514,HP:0000571,HP:0000597,HP:0000657,HP:0001152,HP:0001249,HP:0001251,HP:0001513,HP:0002061,HP:0003438,HP:0006855,HP:0100275,HP:0100543 | GRCh37 | /path_to/gvcftools_E261621.g.vcf | WES |
| FAM0004372 | FAM0004372 | E993720 | | | female | TRUE | FALSE | HP:0001272,HP:0002136,HP:0003680,HP:0012104,HP:0100275 | GRCh37 | /path_to/gvcftools_E993720.g.vcf | WES |
| FAM0004372 | FAM0004372 | E707589 | | | male | FALSE | FALSE | | GRCh37 | /path_to/gvcftools_E707589.g.vcf | WES |

| | | | | | | | | | | | |
|---|---|---|---|---|---|---|---|---|---|---|---|
| FAM0004574 | FAM0004574 | E287287 | E258122 | E671780 | female | TRUE | TRUE | HP:0000407,HP:0000750,HP:0001197,HP:0001943,HP:0001992,HP:0002011,HP:0002015,HP:0002194,HP:0002384,HP:0002474,HP:0002490,HP:0002910,HP:0002921,HP:0002922,HP:0003074,HP:0003128,HP:0003643,HP:0004337,HP:0007359,HP:0010862,HP:0010863,HP:0011097,HP:0011346.HP:0011 | GRCh37 | /path_to/gvcftools_E287287.g.vcf | WES |
| FAM0004574 | FAM0004574 | E258122 | | | male | FALSE | FALSE | | GRCh37 | /path_to/gvcftools_E258122.g.vcf | WES |
| FAM0004574 | FAM0004574 | E671780 | | | female | FALSE | FALSE | | GRCh37 | /path_to/gvcftools_E671780.g.vcf | WES |

| | | | | | | | | | | | |
|---|---|---|---|---|---|---|---|---|---|---|---|
| FAM0004669 | FAM0004669 | E470458 | E754203 | E280783 | male | TRUE | TRUE | HP:0000486,HP:0000505,HP:0000733,HP:0000750,HP:0001141,HP:0001250,HP:0001257,HP:0001263,HP:0001266,HP:0001317,HP:0001332,HP:0001344,HP:0001347,HP:0002072,HP:0002151,HP:0002194,HP:0002376,HP:0002451,HP:0002453,HP:0003201,HP:0003236,HP:0003487,HP:0007256.HP:0008 | GRCh37 | /path_to/gvcftools_E470458.g.vcf | WES |
| FAM0004669 | FAM0004669 | E280783 | | | female | FALSE | FALSE | | GRCh37 | /path_to/gvcftools_E280783.g.vcf | WES |
| FAM0004669 | FAM0004669 | E754203 | | | male | FALSE | FALSE | | GRCh37 | /path_to/gvcftools_E754203.g.vcf | WES |

| | | | | | | | | | | | |
|---|---|---|---|---|---|---|---|---|---|---|---|
| FAM0004672 | FAM0004672 | E313710 | E959423 | E717224 | male | TRUE | TRUE | HP:0001285,HP:0001332,HP:0001511,HP:0001518,HP:0001520,HP:0001558,HP:0001561,HP:0001562,HP:0001622,HP:0001787,HP:0002514,HP:0003517,HP:0003561,HP:0004488,HP:0007256,HP:0008071,HP:0008936,HP:0009800,HP:0010519,HP:0011436,HP:0011438,HP:0011451,HP:0012188.HP:0030 | GRCh37 | /path_to/gvcftools_E313710.g.vcf | WES |
| FAM0004672 | FAM0004672 | E959423 | | | male | FALSE | FALSE | | GRCh37 | /path_to/gvcftools_E959423.g.vcf | WES |
| FAM0004672 | FAM0004672 | E717224 | | | female | FALSE | FALSE | | GRCh37 | /path_to/gvcftools_E717224.g.vcf | WES |

| | | | | | | | | | | | |
|---|---|---|---|---|---|---|---|---|---|---|---|
| FAM0005920 | FAM0005920 | E571022 | E907075 | E546332 | male | TRUE | TRUE | HP:0000365,HP:0000717,HP:0001251,HP:0001256,HP:0001260,HP:0001271,HP:0001272,HP:0001332,HP:0001618,HP:0001761, HP:0001765,HP:0002067,HP:0002136,HP:0002166,HP:0002406,HP:0002509,HP:0003438,HP:0004879,HP:0007338 | GRCh37 | /path_to/gvcftools_E571022.g.vcf | WES |
| FAM0005920 | FAM0005920 | E546332 | | | female | FALSE | FALSE | | GRCh37 | /path_to/gvcftools_E546332.g.vcf | WES |
| FAM0005920 | FAM0005920 | E907075 | | | male | FALSE | FALSE | | GRCh37 | /path_to/gvcftools_E907075.g.vcf | WES |

| | | | | | | | | | | | |
|---|---|---|---|---|---|---|---|---|---|---|---|
| FAM0006165 | FAM0006165 | E568630 | E104308 | E002657 | male | TRUE | TRUE | HP:0000639,HP:0000657,HP:0001251,HP:0001257,HP:0001263,HP:0002063,HP:0002080,HP:0100543 | GRCh37 | /path_to/gvcftools_E568630.g.vcf | WES |
| FAM0006165 | FAM0006165 | E002657 | | | female | FALSE | FALSE | | GRCh37 | /path_to/gvcftools_E002657.g.vcf | WES |
| FAM0006165 | FAM0006165 | E104308 | | | male | FALSE | FALSE | | GRCh37 | /path_to/gvcftools_E104308.g.vcf | WES |
| FAM0006891 | FAM0006891 | E597202 | E113690 | E042568 | male | TRUE | TRUE | HP:0000486,HP:0001252,HP:0002151,HP:0002275,HP:0010663 | GRCh37 | /path_to/gvcftools_E597202.g.vcf | WES |
| FAM0006891 | FAM0006891 | E113690 | | | male | FALSE | FALSE | | GRCh37 | /path_to/gvcftools_E113690.g.vcf | WES |
| FAM0006891 | FAM0006891 | E042568 | | | female | FALSE | FALSE | | GRCh37 | /path_to/gvcftools_E042568.g.vcf | WES |

| | | | | | | | | | | | |
|---|---|---|---|---|---|---|---|---|---|---|---|
| FAM0007156 | FAM0007156 | E473734 | E193041 | E734772 | female | TRUE | TRUE | HP:0000219,HP:0000268,HP:0000278,HP:0000369,HP:0000377,HP:0000582,HP:0000664,HP:0001263,HP:0001270,HP:0001273,HP:0001290,HP:0001320,HP:0001382,HP:0001852,HP:0001864,HP:0002007,HP:0002307,HP:0005274,HP:0005487,HP:0007429,HP:0011470,HP:0011918 | GRCh37 | /path_to/gvcftools_E473734.g.vcf | WES |
| FAM0007156 | FAM0007156 | E193041 | | | male | FALSE | FALSE | | GRCh37 | /path_to/gvcftools_E193041.g.vcf | WES |
| FAM0007156 | FAM0007156 | E734772 | | | female | FALSE | FALSE | | GRCh37 | /path_to/gvcftools_E734772.g.vcf | WES |

| | | | | | | | | | | | |
|---|---|---|---|---|---|---|---|---|---|---|---|
| FAM0007265 | FAM0007265 | E513765 | E597171 | E720071 | female | TRUE | TRUE | HP:0000154,HP:0000341,HP:0000350,HP:0000455,HP:0000718,HP:0001249,HP:0001250,HP:0001864,HP:0009748,HP:0011918,HP:0012810 | GRCh37 | /path_to/gvcftools_E513765.g.vcf | WES |
| FAM0007265 | FAM0007265 | E597171 | | | male | FALSE | FALSE | | GRCh37 | /path_to/gvcftools_E597171.g.vcf | WES |
| FAM0007265 | FAM0007265 | E720071 | | | female | FALSE | FALSE | | GRCh37 | /path_to/gvcftools_E720071.g.vcf | WES |

| | | | | | | | | | | | |
|---|---|---|---|---|---|---|---|---|---|---|---|
| FAM0007273 | FAM0007273 | E867414 | E993251 | E398289 | male | TRUE | TRUE | HP:0000098,HP:0000729,HP:0000736,HP:0000821,HP:0001270,HP:0001288,HP:0001762,HP:0001769,HP:0001773,HP:0001799,HP:0002342,HP:0002465,HP:0003763,HP:0004322,HP:0008071,HP:0008872,HP:0010862,HP:0012469 | GRCh37 | /path_to/gvcftools_E867414.g.vcf | WES |
| FAM0007273 | FAM0007273 | E398289 | | | female | FALSE | FALSE | | GRCh37 | /path_to/gvcftools_E398289.g.vcf | WES |
| FAM0007273 | FAM0007273 | E993251 | | | male | FALSE | FALSE | | GRCh37 | /path_to/gvcftools_E993251.g.vcf | WES |

| | | | | | | | | | | | |
|---|---|---|---|---|---|---|---|---|---|---|---|
| FAM0007279 | FAM0007279 | E599679 | E577417 | E255248 | male | TRUE | TRUE | HP:0000002,HP:0000218,HP:0000240,HP:0000324,HP:0000396,HP:0000411,HP:0000490,HP:0000527,HP:0000817,HP:0001156,HP:0001250,HP:0001263,HP:0001328,HP:0002121,HP:0002197,HP:0002342,HP:0002719,HP:0008872,HP:0009940 | GRCh37 | /path_to/gvcftools_E599679.g.vcf | WES |
| FAM0007279 | FAM0007279 | E577417 | | | male | FALSE | FALSE | | GRCh37 | /path_to/gvcftools_E577417.g.vcf | WES |
| FAM0007279 | FAM0007279 | E255248 | | | female | FALSE | FALSE | | GRCh37 | /path_to/gvcftools_E255248.g.vcf | WES |

| | | | | | | | | | | | |
|---|---|---|---|---|---|---|---|---|---|---|---|
| FAM0007311 | FAM0007311 | E499319 | E662416 | E028855 | male | TRUE | TRUE | HP:0001249,HP:0001679,HP:0009062 | GRCh37 | /path_to/gvcftools_E499319.g.vcf | WES |
| FAM0007311 | FAM0007311 | E028855 | | | female | FALSE | FALSE | | GRCh37 | /path_to/gvcftools_E028855.g.vcf | WES |
| FAM0007311 | FAM0007311 | E662416 | | | male | FALSE | FALSE | | GRCh37 | /path_to/gvcftools_E662416.g.vcf | WES |
| FAM0007318 | FAM0007318 | E822195 | E802554 | E416739 | female | TRUE | TRUE | HP:0000483,HP:0000540,HP:0000964,HP:0001216,HP:0001249 | GRCh37 | /path_to/gvcftools_E822195.g.vcf | WES |
| FAM0007318 | FAM0007318 | E802554 | | | male | FALSE | FALSE | | GRCh37 | /path_to/gvcftools_E802554.g.vcf | WES |
| FAM0007318 | FAM0007318 | E416739 | | | female | FALSE | FALSE | | GRCh37 | /path_to/gvcftools_E416739.g.vcf | WES |

| | | | | | | | | | | | |
|---|---|---|---|---|---|---|---|---|---|---|---|
| FAM0007319 | FAM0007319 | E547381 | E404660 | E283620 | male | TRUE | TRUE | HP:0000023,HP:0000341,HP:0000430,HP:0000540,HP:0000664,HP:0000750,HP:0000954,HP:0001249,HP:0001263,HP:0001290, HP:0001385,HP:0002020,HP:0002650,HP:0003693,HP:0003763 | GRCh37 | /path_to/gvcftools_E547381.g.vcf | WES |
| FAM0007319 | FAM0007319 | E404660 | | | male | FALSE | FALSE | | GRCh37 | /path_to/gvcftools_E404660.g.vcf | WES |
| FAM0007319 | FAM0007319 | E283620 | | | female | FALSE | FALSE | | GRCh37 | /path_to/gvcftools_E283620.g.vcf | WES |

| | | | | | | | | | | | |
|---|---|---|---|---|---|---|---|---|---|---|---|
| FAM0007339 | FAM0007339 | E038467 | E431719 | E936591 | female | TRUE | TRUE | HP:0000278,HP:0000494,HP:0000718,HP:0000733,HP:0000750,HP:0000958,HP:0001263,HP:0001290,HP:0001508,HP:0001511, HP:0002099,HP:0002194,HP:0002360,HP:0004209,HP:0005274,HP:0007687,HP:0008527,HP:0008872,HP:0010864 | GRCh37 | /path_to/gvcftools_E038467.g.vcf | WES |
| FAM0007339 | FAM0007339 | E936591 | | | female | FALSE | FALSE | | GRCh37 | /path_to/gvcftools_E936591.g.vcf | WES |
| FAM0007339 | FAM0007339 | E431719 | | | male | FALSE | FALSE | | GRCh37 | /path_to/gvcftools_E431719.g.vcf | WES |

| | | | | | | | | | | | |
|---|---|---|---|---|---|---|---|---|---|---|---|
| FAM0007343 | FAM0007343 | E440074 | E336032 | E717440 | male | TRUE | TRUE | HP:0000201,HP:0000308,HP:0000316,HP:0000343,HP:0000368,HP:0000463,HP:0001274,HP:0001671,HP:0005301,HP:0011430 | GRCh37 | /path_to/gvcftools_E440074.g.vcf | WES |
| FAM0007343 | FAM0007343 | E717440 | | | female | FALSE | FALSE | | GRCh37 | /path_to/gvcftools_E717440.g.vcf | WES |
| FAM0007343 | FAM0007343 | E336032 | | | male | FALSE | FALSE | | GRCh37 | /path_to/gvcftools_E336032.g.vcf | WES |

| | | | | | | | | | | | |
|---|---|---|---|---|---|---|---|---|---|---|---|
| FAM0007344 | FAM0007344 | E666478 | E152758 | E508534 | female | TRUE | TRUE | HP:0000256,HP:0000286,HP:0000337,HP:0000689,HP:0000718,HP:0000750,HP:0001233,HP:0001256,HP:0002194,HP:0002360,HP:0004692,HP:0011968,HP:0100400 | GRCh37 | /path_to/gvcftools_E666478.g.vcf | WES |
| FAM0007344 | FAM0007344 | E152758 | | | male | TRUE | FALSE | HP:0000256,HP:0000337,HP:0000750,HP:0001249,HP:0001274,HP:0001510 | GRCh37 | /path_to/gvcftools_E152758.g.vcf | WES |
| FAM0007344 | FAM0007344 | E508534 | | | female | FALSE | FALSE | | GRCh37 | /path_to/gvcftools_E508534.g.vcf | WES |

| | | | | | | | | | | | |
|---|---|---|---|---|---|---|---|---|---|---|---|
| FAM0008397 | FAM0008397 | E724379 | E932126 | E772691 | female | TRUE | TRUE | HP:0000505,HP:0000735,HP:0001249,HP:0001285,HP:0002353,HP:0006958,HP:0010837,HP:0011327,HP:0012000,HP:0012443,HP:0012705,HP:0200134 | GRCh37 | /path_to/gvcftools_E724379.g.vcf | WES |
| FAM0008397 | FAM0008397 | E932126 | | | male | FALSE | FALSE | | GRCh37 | /path_to/gvcftools_E932126.g.vcf | WES |
| FAM0008397 | FAM0008397 | E772691 | | | female | FALSE | FALSE | | GRCh37 | /path_to/gvcftools_E772691.g.vcf | WES |
| FAM0004547 | FAM0004547 | E100304 | E457060 | E419467 | male | TRUE | TRUE | HP:0000252,HP:0000639,HP:0001290,HP:0001344,HP:0002540,HP:0012758 | GRCh38 | /path_to/FAM0004547_FAM0004547_E100304_liftover_accepted.g.vcf | WES |

| | | | | | | | | | | | |
|---|---|---|---|---|---|---|---|---|---|---|---|
| FAM0004547 | FAM0004547 | E457060 | | | male | FALSE | FALSE | | GRCh38 | /path_to/FAM0004547_FAM0004547_E457060_liftover_accepted.g.vcf | WES |
| FAM0004547 | FAM0004547 | E419467 | | | female | FALSE | FALSE | | GRCh38 | /path_to/FAM0004547_FAM0004547_E419467_liftover_accepted.g.vcf | WES |
| FAM0004549 | FAM0004549 | E925075 | E833842 | E466771 | male | TRUE | TRUE | HP:0000977,HP:0001373,HP:0002650,HP:0003198 | GRCh38 | /path_to/FAM0004549_FAM0004549_E925075_liftover_accepted.g.vcf | WES |
| FAM0004549 | FAM0004549 | E833842 | | | male | FALSE | FALSE | | GRCh38 | /path_to/FAM0004549_FAM0004549_E833842_liftover_accepted.g.vcf | WES |
| FAM0004549 | FAM0004549 | E466771 | | | female | FALSE | FALSE | | GRCh38 | /path_to/FAM0004549_FAM0004549_E466771_liftover_accepted.g.vcf | WES |

| | | | | | | | | | | | |
|---|---|---|---|---|---|---|---|---|---|---|---|
| FAM0004916 | FAM0004916 | E186172 | E287418 | E830376 | female | TRUE | TRUE | HP:0000750,HP:0001276,HP:0002079,HP:0003394,HP:0012448,HP:0012758 | GRCh38 | /path_to/FAM0004916_FAM0004916_E186172_liftover_accepted.g.vcf | WES |
| FAM0004916 | FAM0004916 | E287418 | | | male | FALSE | FALSE | | GRCh38 | /path_to/FAM0004916_FAM0004916_E287418_liftover_accepted.g.vcf | WES |
| FAM0004916 | FAM0004916 | E830376 | | | female | FALSE | FALSE | | GRCh38 | /path_to/FAM0004916_FAM0004916_E830376_liftover_accepted.g.vcf | WES |

Supplementary table 6 **Example of sample sheet containing Solve-RD research cohort**
